# Assessing the potential population-level impacts of HIV self-testing distribution among key populations in Côte d’Ivoire, Mali, and Senegal: a mathematical modelling analysis

**DOI:** 10.1101/2023.08.23.23294498

**Authors:** Romain Silhol, Mathieu Maheu-Giroux, Nirali Soni, Arlette Simo Fotso, Nicolas Rouveau, Anthony Vautier, Clémence Doumenc-Aïdara, Olivier Geoffroy, Kouassi Noel N’Guessan, Younoussa Sidibé, Odé Kanku Kabemba, Papa Alioune Gueye, Christinah Mukandavire, Peter Vickerman, Abdelaye Keita, Cheikh Tidiane Ndour, Eboi Ehui, Joseph Larmarange, Marie-Claude Boily, ATLAS Team

## Abstract

**Background:** A third of people living with HIV (PLHIV) in Western Africa had an undiagnosed infection in 2020. In 2019-2021, the ATLAS programme has distributed a total of 380 000 HIV self-testing (HIVST) kits to key populations (KP) including female sex workers (FSW) and men who have sex with men (MSM), and their partners in Côte d’Ivoire, Mali and Senegal. We predicted the potential impact of ATLAS and of national HIVST scale-up strategies among KP.

**Methods:** A deterministic model of HIV transmission was calibrated to country-specific empirical HIV and intervention data over time. We simulated scenarios reflecting 1) the actual ATLAS HIVST distribution only over 2019-2021 (∼2% of all tests done in countries), and 2) ATLAS followed by a scale-up of HIVST distribution to KP (total of ∼570 000 kits distributed each year). Impacts on HIV diagnosis, new HIV infections and deaths were derived using counterfactual scenarios without HIVST.

**Findings:** ATLAS was predicted to substantially increase HIV diagnosis among KP by the end of 2021, especially among MSM in Mali (9·3 percentage point [pp] increase), and a 1·0pp increase overall. ATLAS might have averted a median of 706 new HIV infections among KP over 2019-2028 in the 3 countries combined, especially among MSM, and 1794 new HIV infections (0·4-3·3% of all new HIV infections across countries) and 591 HIV-related deaths overall. HIVST scale-up increased HIV diagnosis at the end of 2028 by around 8pp among FSW and 33pp among MSM in every country. Overall increases ranged from 1·0pp (Côte d’Ivoire) to 11·0pp (Senegal). HIVST scale-up may avert 3-5% of new HIV infections among FSW, 3-10% among FSW clients, and 20-28% among MSM across countries (and 2-16% overall), and avert 13-18% of HIV-related deaths among MSM over 2019-2028.

**Interpretation:** Scaling-up HIVST distribution among KP in Western Africa may substantially attenuate disparities in access to HIV testing and help reduce HIV infections and deaths among KP and their partners.

**Funding:** Unitaid MRC

## Introduction

HIV testing is the first step in the HIV prevention and care cascade. The Joint United Nations Programme on HIV/AIDS (UNAIDS) established a target where less than 5% of PLHIV should have an undiagnosed infection by 2025.^1^ A third of people living with HIV (PLHIV) in Western Africa remained undiagnosed in 2020,^2^ hindering progress towards HIV elimination. This large gap has been partly attributed to low levels of diagnosis among key populations (KP), including female sex workers (FSW) and men who have sex with men (MSM), among others.^2,3^ KP who are disproportionally burdened by HIV, regularly experience stigma and discrimination, which can result in less frequent use of HIV testing services compared to non-KP.^2,4–7^ HIV self-testing (HIVST) is a relatively new confidential testing modality which has been proposed as a possible solution to reduce diagnosis gaps.^8–10^ However, given its anonymous nature, it is usually challenging to track and characterise users of self-test kits, linkage to care following reactive self-tests, and to evaluate HIVST’s direct (to users) and indirect (to non-users by averting chains of transmission) impact on HIV transmission and morbidity.^11^

In Western Africa, HIV prevalence among adults (aged 15+ years) was relatively low in Mali (0·9%) and Senegal (0·4%), but higher in Côte d’Ivoire (2·2%) in 2022.^12^ However, HIV prevalence levels among KP are about 10 times higher than national averages in Senegal and Mali (e.g. currently around 28% among MSM in Senegal)^13^, and 3 times higher in Côte d’Ivoire,^14,15^ with clients and partners of KP estimated to have acquired 28% of all new HIV infections in Western and Central Africa in 2021.^16^ It is concerning that levels of HIV diagnosis are lower among most KP compared to non-KP in these countries, and especially among MSM.^4,17,18^ Addressing the unmet testing needs of KP could increase population-level viral suppression and reduce HIV infections and mortality among KP, their partners, and the overall population.

The global health initiative UNITAID^19^ implemented the ATLAS programme through a partnership with the NGO Solthis and the French institute for sustainable development. ATLAS aimed to address the diagnosis gap and evaluate the potential impact of HIVST in Côte d’Ivoire, Mali and Senegal,^20^ with an emphasis on secondary distribution of HIVST whereby KP directly reached by the programme could re-distribute some of their HIVST kits to their sexual partners, clients, and/or peers.^20–22^ In addition to the distribution of a total of 380 000 self-test kits to people at higher risk of HIV (of which 340 000 were distributed through activities targeting FSW and MSM) over 2019-2021, the programme included different work packages to understand the practicality, costs and population-level impact of HIVST, combining population studies,^11,23^ qualitative studies,^21,24,25^ economic analyses,^26^ and mathematical modelling.

Given the challenges in empirically measuring the impact of HIVST due to its anonymous nature, the potential population-level impact of HIVST on HIV infections in West Africa has not been quantified despite its potential.^27–31^ We aimed to estimate the impact of the ATLAS programme, and a national scale-up of HIVST distribution among FSW and MSM on HIV diagnosis and treatment coverage, HIV incidence and mortality among KP and the overall population in Côte d’Ivoire, Mali and Senegal using country-specific HIV dynamic transmission models incorporating detailed data from ATLAS and other local studies.

## Methods

### Mathematical model structure

We adapted a deterministic compartmental model of HIV transmission in Côte d’Ivoire,^32,33^ parameterised and fitted to country-specific demographic, behavioural, HIV epidemiological and intervention data in Côte d’Ivoire, Mali and Senegal separately over 1980-2020 (details in supplement).

The models simulate an open and growing population stratified in eight groups: FSW, MSM who report sex with female and male partners (MSMW) or only male partners (MSME), clients of FSW (“clients”), low risk (0-1 partner per year) and intermediate risk (>1 partner per year) non-KP females, and low risk (0-2 partners per year) and intermediate risk non-KP males (>2 partners per year; **Figure S1a**). Four age groups were represented (15-19, 20-24, 25-49, and 50-59 years old; **Figure S1b**). Individuals enter the sexually active population at age 15 and leave due to ageing at age 59, or via background- and HIV-related mortality (**Figure S1b**). Immigration in the 25-49-years age groups takes place as informed by demographic data.^34^ HIV transmission occur during heterosexual non-commercial and commercial partnerships, and non-commercial sex between men (**Table S1a,b**, **Figure S1a**).

People who acquire the virus progress through five infection stages, from acute to AIDS (**Figure S1c**). HIV diagnosis and linkage to treatment are explicitly modelled, including self- and conventional testing and confirmation tests following reactive self-tests (**Figure S1d**). Once diagnosed, PLHIV can initiate antiretroviral therapy (ART) and experience reduced HIV-related mortality; a fraction of treated PLHIV is virally suppressed and can’t transmit HIV.

Per-capita HIV acquisition rates depend on the annual number of new partners and sex acts with/without condoms, sexual mixing by risk/age group, HIV prevalence, coverage of viral suppression among sexual partners, and per-act HIV transmission probabilities (**Tables S1a-d**).

The model represents changes in the levels of intervention (condom use, HIV testing, and ART) over time by risk group and age using available data (**Tables S1d,e**) for 1991-2020 and future levels of condom use and HIV conventional testing remaining constant at the same levels as the last data estimates, and the fraction of PLHIV on ART with a suppressed viral load slowly increasing over time.

### Model parameterisation and fitting

The models were separately parameterised and fitted to country-specific sets of epidemiological and intervention outcomes stratified by sex, risk group, age, and HIV status when available (**Tables S2a-c**) over time within a Bayesian framework. The fitting outcomes included: age distribution, HIV prevalence, annual number of new HIV infections, HIV-related deaths and incidence rate, fraction of people ever tested for HIV and diagnosed with HIV, ART and viral suppression coverage, annual number of conventional tests and fraction that were positive (**Table S2a-c**, **Figures S3-5**). Most fitting outcomes were available for all countries and risk groups, except for clients of FSW, especially in Mali. Details on model fitting procedure are available in the supplement.

### Data sources

Data informing demography were sourced from the United Nations Population Division (UNPD) World Prospect (2019 revision),^35^ whereas most sexual behaviours (including sexual mixing), trends in condom use and HIV prevalence among non-KP were taken from countries-representative population-based surveys.^36–38^ The relative size of each KP and clients of FSW varied across simulations to reflect heterogeneities in available country-specific studies but was constant over time within simulations (**Table S1a, Figures S3c,S4c,S5c).** Data suggested similar proportions of FSW across countries but lower sizes of MSM populations in Mali^39,40^ and Senegal,^39,41^ compared to Côte d’Ivoire.^42–45^ The sizes of the client population were estimated by triangulating the estimated sizes of the FSW population, the number of sex acts paid by clients and paid for by FSW. As no client data was available for Mali, we used estimates of the number of sex acts paid by clients in Côte d’Ivoire. Sexual behaviours, trends in condom use and HIV prevalence of KP were based on local surveys in the three countries (**Tables S1,S2**).

Importantly, the annual HIV testing rates and fractions of PLHIV ever tested was mostly sourced from secondary analysis of local population-based and KP surveys by age, sex, and HIV status when available (**Tables S2a-c**). The fraction of all female and male PLHIV with a diagnosed infection over time were based on national UNAIDS Shiny90 model-based estimates,^46^ whereas self-reported knowledge of status among KP were taken from local surveys (**Tables S2a-c**). The coverage of ART by sex and fraction of treated PLHIV with a suppressed viral load by sex over time were obtained from UNAIDS^12^ and KP surveys (**Tables S2a-c**). Programmatic data on the annual number and positivity of conventional tests was from UNAIDS Shiny90.^12^ Biological parameters and condom/ART efficacy parameters were sourced from published literature (**Table S1c**).

We retained all simulations from original samples of 50 million parameter sets (sampled using Latin Hypercube) which agreed with pre-specified fitting targets (**Tables S2a-c**). Then, the 100 fitted simulations with the highest likelihood were identified for each country (details in supplement). The resulting posterior parameter sets were used to simulate the counterfactual scenario without HIVST (“no HIVST”) and the HIVST distribution scenarios.

### Modelling analysis

#### Main HIVST intervention scenarios among KPs

In addition to the counterfactual without any HIVST, we simulated 2 main intervention scenarios representing the most likely HIVST distribution and use over the modelled period (**Figure 1**, **Table S5**). The “ATLAS-only” scenario represented ATLAS’s HIVST distribution through activities targeting FSW an MSM over 2019-2021 (**Tables S3,4**, **Figures S2,3**), and no HIVST distribution from January 2022, to estimate the impact of the sole ATLAS program. The “HIVST scale-up” scenario represented the ATLAS program over 2019-2021 and further scale-up of HIVST distribution in FSW and MSM distribution channels increasing from January 2022 onwards and assuming constant numbers of kits distributed each year from 2025 onward (**Figure S3**).

**Figure 1:**
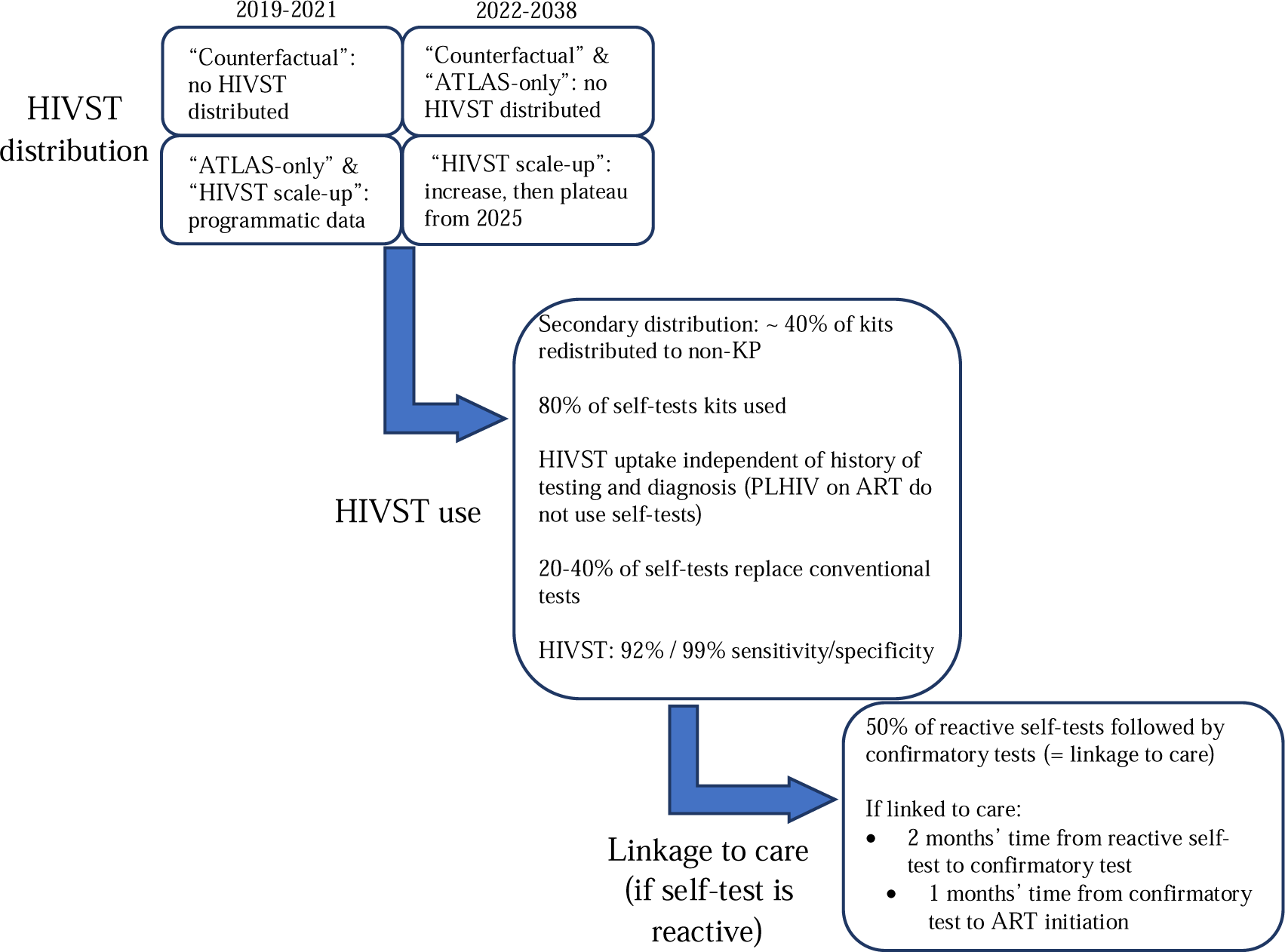
Model assumptions of the counterfactual and the two HIVST intervention scenarios using ATLAS programmatic data, ATLAS studies and the literature (fraction of tests used and tests sensitivity/specificity). Further description is available in Supplement **Table S5**.

We used ATLAS programmatic data on the number of distributed kits in each distribution channel over 2019-2021 and specific surveys designed to estimate the number of kits received by risk groups through secondary distribution (i.e., kits redistributed by those directly reached by the program to their partners, clients, and peers), and levels of linkage to care following a reactive self-test (**Tables S3-5**).^47^ Our HIVST scale-up scenarios assumed HIVST distribution could reach our pre-defined target of two self-tests distributed to 95% of “eligible” FSW or MSM primary contacts (i.e. without HIV or untreated PLHIV, regardless of their awareness of HIV status, based on the World Health Organisation (WHO) recommendations of testing KP twice a year^48^) from 2025 onwards. This plateau in number of tests distributed corresponded to medians of 365 000, 89 000, and 103 000 kits distributed annually through FSW and MSM in 2025 in Côte d’Ivoire, Mali, and Senegal, respectively (**Figure S3a-c**). After accounting for secondary distribution, this meant that 0·8 and 1·6 kits were received per “eligible” FSW and MSM in 2025, respectively (**Figure S3d-f**). HIV self-tests among KP would represent 48·7-86·9% of all tests done by KP (including conventional tests, and 6·9-15·2% of all HIV tests done) across countries in 2025.

In addition, based on East African data,^49^ we assumed that 80% of HIVST distributed are used (with uniform use across population groups). ATLAS survey data suggest that 50% of reactive self-tests were confirmed with conventional test and linked to care.^47^ From programmatic data from HIV testing services in Côte d’Ivoire and Senegal, we considered that 20% (Côte d’Ivoire), 30% (Mali) and 40% (Senegal) of ATLAS self-tests replaced conventional tests.^23^ Finally, HIVST was assumed to have 92% sensitivity and 99% specificity (false positive tests are disconfirmed after linkage to care or a negative self-test).^50^

#### Impact of HIVST intervention scenarios

We assessed the impact of ATLAS-only (scenario 2) and scale-up (scenario 3) on the absolute increase in the proportion of PLHIV diagnosed at the end of 2021 (scenario 2) and end of 2028 and 2038 (scenario 3), the cumulative numbers of additional HIV diagnosis and ART initiations and the fraction and absolute number of new HIV infections and HIV-related deaths averted over 2019-2021 and 2019-2028 (scenario 2) and over 2019-2028 and 2019-2038 (scenario 3), all compared to the counterfactual (scenario 1). A sensitivity analysis, presented in the supplement, assessed the influence of key assumptions (e.g. fraction of confirmed reactive self-tests) of our HIVST scale-up scenario on new HIV infections deaths over 2019-2028.

We predicted the change in the median time from HIV infection to diagnosis (or death before being diagnosed) between the HIVST scale-up and the counterfactual scenarios by calculating the time from January 2020 when half of the undiagnosed PLHIV would have become diagnosed, assuming no new infections over the period.

We report median estimates and 90% uncertainty interval (90%UI, 5^th^ and 95^th^ percentiles of the distribution) of model predictions based on the 100 posterior parameter sets.

## Results

### Model fitting and HIV epidemic contexts

Our model reproduced available empirical demographic, epidemiological and intervention data across risk groups in each country (**Figures S4-6**), with between 579 and 1550 simulations agreeing with all pre-specified fitting targets. National HIV prevalence was highest in Côte d’Ivoire, while prevalent HIV infections were more concentrated among KP in Mali and Senegal (**Table 1**). Côte d’Ivoire had smaller median fraction of PLHIV with an undiagnosed infection among KPs (18%), than Mali (28%), and Senegal (57%) (**Figure S7**). In line with several country-specific empirical estimates, our model reflected smaller MSM population size in Mali and Senegal (0·5%) compared to Côte d’Ivoire (1·2%) (**Table 1**, **Figures S4c,S5c,S6c**).

**Table 1.**
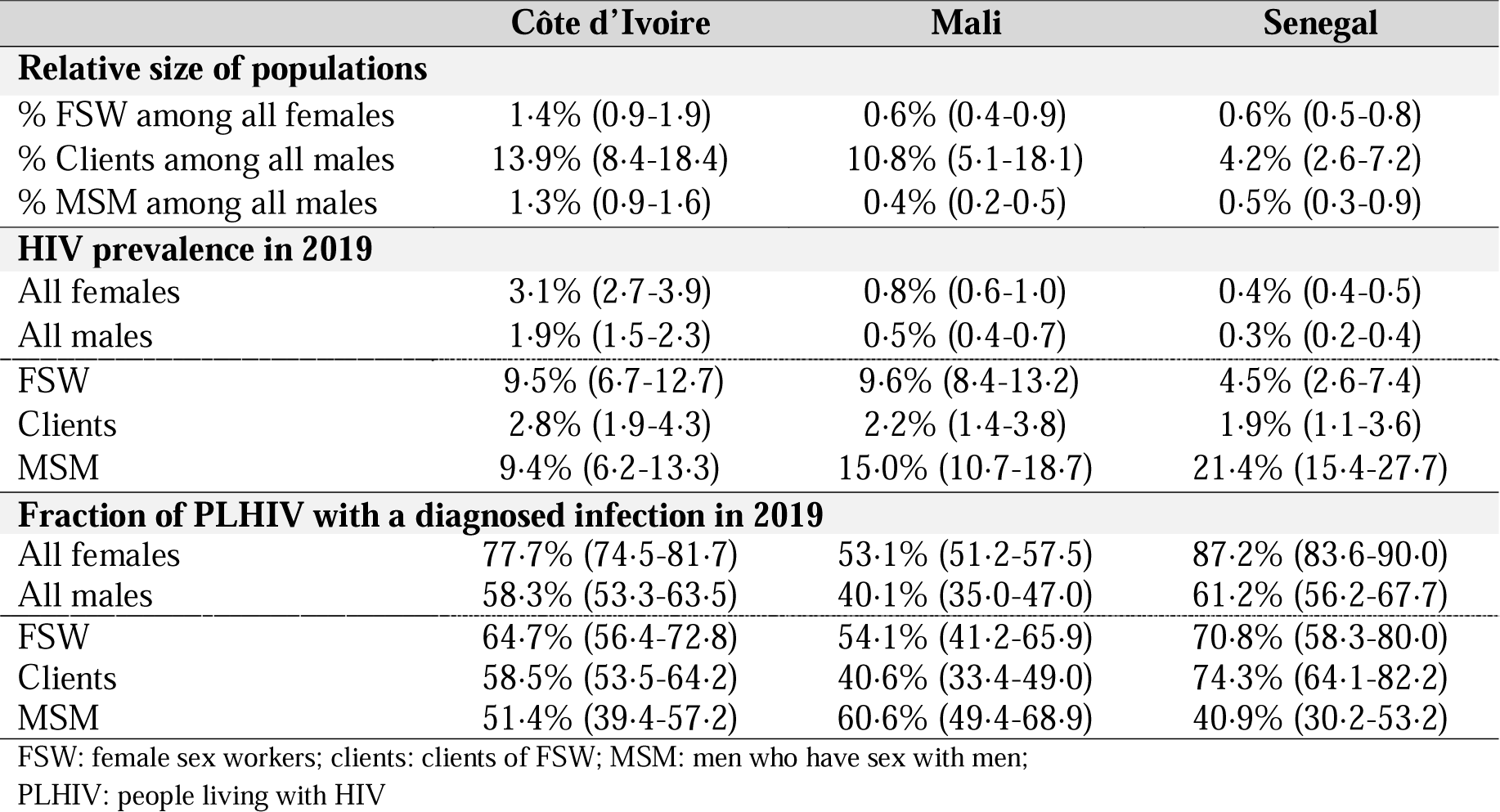
HIV epidemic characteristics of the three ATLAS countries: Model-based estimates of the KP and clients of FSW population size, HIV prevalence, and diagnosis at baseline. Median and 90%UI (5^th^ and 95^th^ percentiles) of posterior distributions in January 2019 are shown.

### Impact of the ATLAS program (HIVST distribution over 2019-2021)

Our model predicted that ATLAS may have led to 681 (211-930) additional HIV diagnoses in Côte d’Ivoire, 518 (327-863) in Mali, and 268 (39-673) in Senegal during the 3-year program compared to the counterfactual, 80% of which were among KPs across all three countries (Figure 2a and **S8a**).

**Figure 2:**
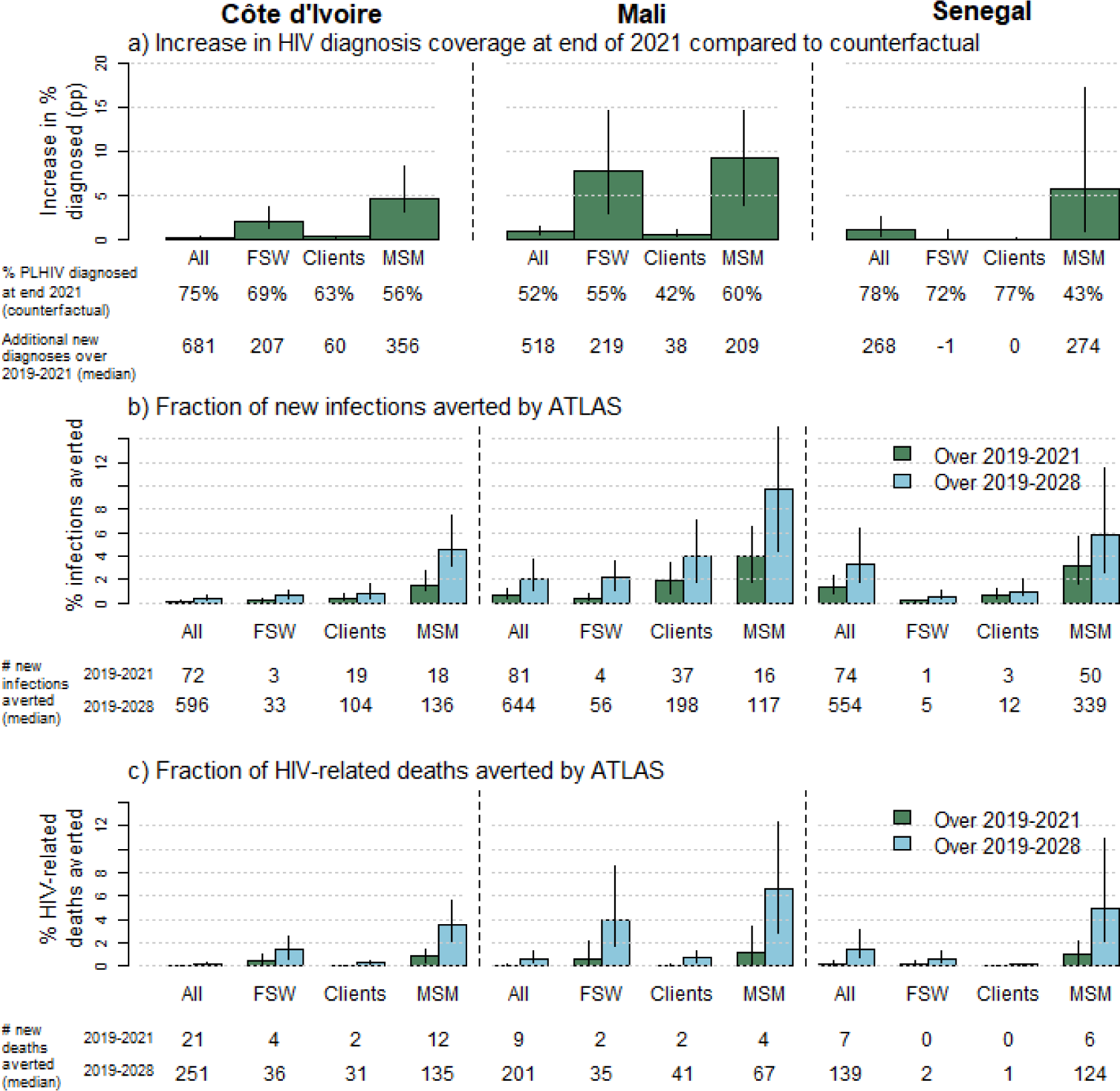
Estimated impact of the ATLAS only scenario compared to the “no-HIVST” counterfactual scenario: a) Percentage-points increases in HIV diagnosis coverage at the end of 2021, and fraction of cumulative b) HIV infections (c) and HIV-related deaths averted by ATLAS over 2019-2021 (green bars) and 2019-2028 (light blue bars). Bars height represent median of model estimates, whereas error bars represent 90%UI (5^th^ and 95^th^ percentiles of estimates across model estimates). Percentages on panel a) represent coverage of HIV diagnosis at the end of 2021 under the counterfactual (“no-HIVST”) scenario. Numbers under the panels represent the median estimates of the cumulative numbers of a) new HIV diagnoses over 2019-2021, b) new HIV infections averted over 2019-2021 (green) and 2019-2028 (light blue), and c) HIV-related deaths averted over 2019-2021 (green) and 2019-2028 (light blue), compared to the counterfactual. FSW: female sex workers; Clients: clients of FSW; MSM: men who have sex with men; PLHIV: people living with HIV

Largest increases in HIV diagnosis coverage due to ATLAS were predicted to have occurred among KP, and were highest among MSM with 4·6pp (3·1-8·3) increases compared to the counterfactual without HIVST in Côte d’Ivoire, 9·3pp (3·9-14·7) in Mali, and 5·7pp (1·0-17·2) in Senegal (Figure 2a, **Table S6a**). This corresponds to overall increases of ∼1pp in Mali and Senegal, and <0·5pp in Côte d’Ivoire at the end of 2021. The ATLAS impact on ART initiations over 2019-2021 were similar to the impact on new diagnoses (**Figure S9**).

Over 2019-2028, ATLAS may avert ∼1800 new HIV infections and ∼600 HIV-related deaths across the 3 countries, 50% of which would be among KPs and 15% among clients (Figure 2b**,c**, **S8b,c**). Most new HIV infections and deaths averted by ATLAS in Senegal over 2019-2028 would be among MSM. In relative terms, the fraction of infections averted was higher among MSM across all three countries (with a maximum of 9·7% (4·4-16·1) in Mali), and among FSW clients (e.g., 3·9% (1·8-7·1) in Mali), due to higher number of kits having been distributed to their sexual partners. Overall, the program is expected to avert 3·3% (1·7-6·3) of all new HIV infections occurring in Senegal over 2019-2028, 0·4% (0·3-0·6) in Côte d’Ivoire and 2·1% (1·1-3·7) in Mali, whilst it could avert 1·5% of all HIV-related deaths across countries over the period (Figure 2b).

### Impact of HIVST distribution to KP scale-up over 2019-2038

When scaling-up HIVST, the proportion of PLHIV diagnosed with HIV increased to 90% among MSM after 20 years in Côte d’Ivoire and Mali (vs ∼60% in the counterfactual), and 85% in Senegal (vs 48% in the counterfactual), and up to 83% among FSW in Côte d’Ivoire (vs 77% in the counterfactual), 74% in Mali (vs 60%), and 80% in Senegal (vs 76%). Compared to the counterfactual, the overall proportion of PLHIV diagnosed with HIV after 20 years increased more substantially in Senegal (13·6pp; 6·3-21·4), than Mali (4·8pp; 2·5-9·7) and Côte d’Ivoire (1·2pp; 0·7-2·0) (Figures 3, **S7**, **Table S6a**).

**Figure 3:**
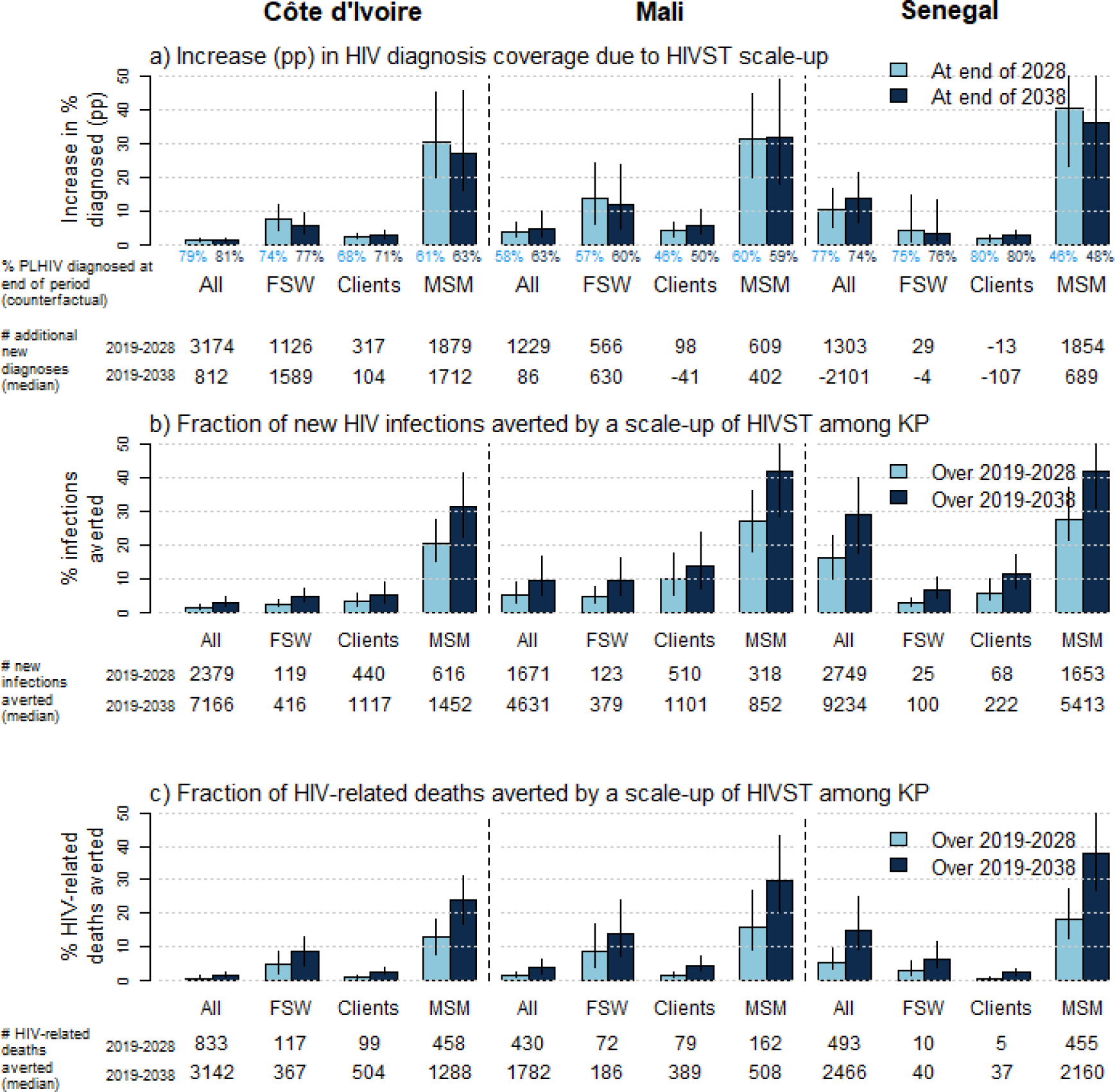
Estimated impact of the “HIVST scale-up” among KP scenario compared to the “no-HIVST” counterfactual scenario: a) Percentage points increases in HIV diagnosis coverage at end of 2029 (light blue bars) and at the end of 2038 (dark blue bars), and fractions of b) new HIV infections and c) HIV-related deaths averted over 2019-2028 (light blue bars) and 2019-2038 (dark blue bars). Bars height represent median of model estimates, whereas error bars represent 90% UI of estimates (5^th^ and 95^th^ percentiles of estimates across model estimates). Percentages on panel a) represent coverage of HIV diagnosis at the end of 2028 (light blue) and 2038 (dark blue) under the counterfactual “no-HIVST” scenario. Numbers under the panels represent the median estimates of the absolute numbers of a) additional HIV new diagnoses over 2019-2028 (light blue) and 2019-2038 (dark blue), b) averted new HIV infections over 2019-2028 (light blue) and 2019-2038 (dark blue), and c) averted HIV-related deaths over 2019-2028 (light blue) and 2019-2038 (dark blue), compared to the counterfactual. FSW: female sex workers; Clients: clients of FSW; MSM: men who have sex with men; KP: Key populations (FSW and MSM)

Overall median time to diagnostic from 2020 was longer for males (medians of 4·2-5·8 years across countries) than females (1·7-4·6 years) in all countries, and was especially long for MSM (4·6-5·7 years, Figure 4). Scaling-up HIVST is expected to reduce this duration by 2 years among MSM, and by 1.5 year among FSW in Mali and Côte d’Ivoire, respectively. There was no change for FSW in Senegal due to relatively high rate of conventional testing and the low number of HIVST kits distributed to them.

**Figure 4:**
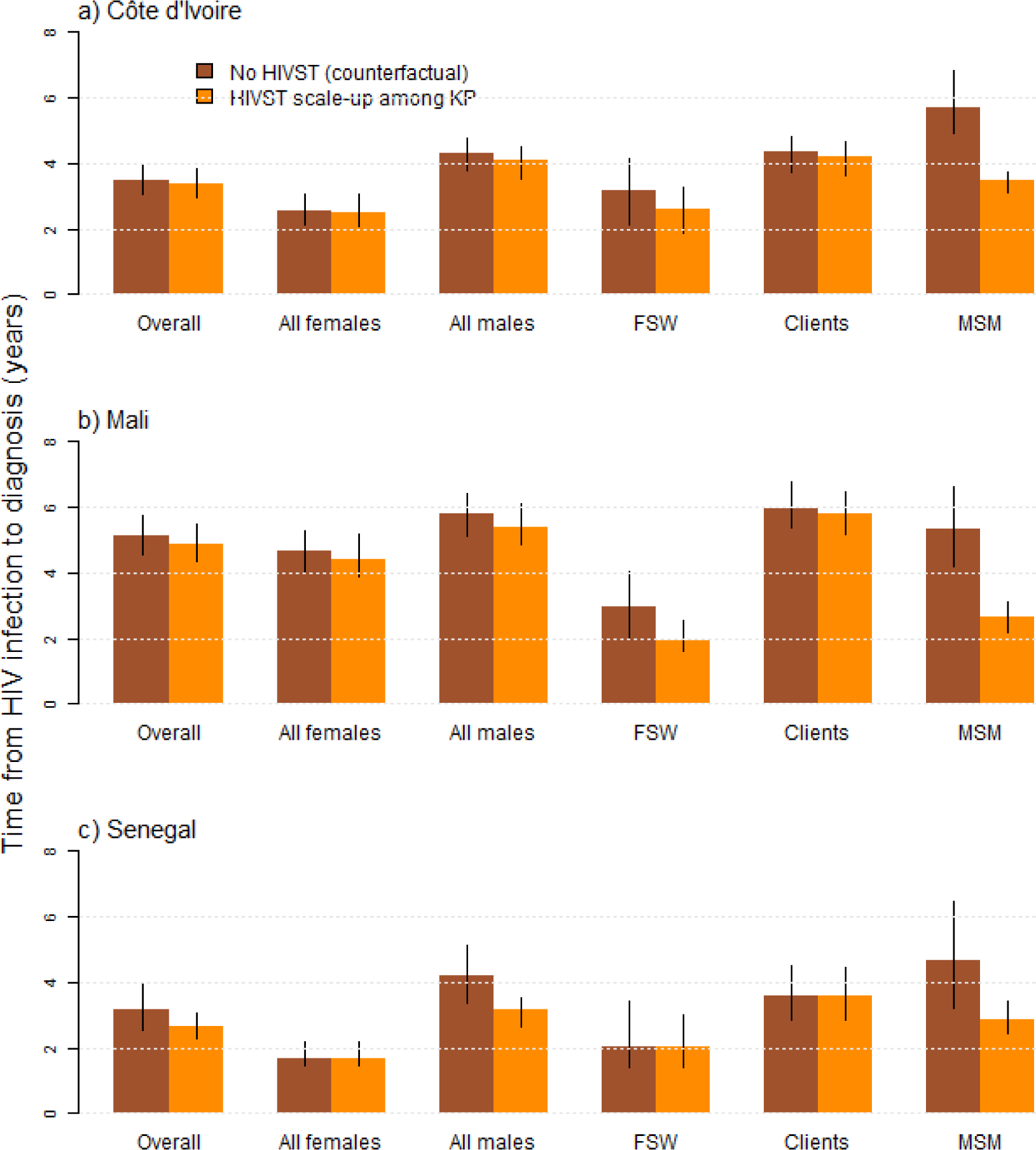
Estimated median time from infection to diagnosis (or to death if it occurs before diagnosis) from January 2020 among modelled risk groups in the three ATLAS countries under the counterfactual scenario assuming no HIVST in the country (brown bars) and a scenario of HIVST scale-up among KP (orange bars). Bars represent median estimates across model fits, whereas error bars represent 90%UI of estimates (5^th^ and 95^th^ percentiles of estimates across model simulations). FSW: female sex workers; Clients: clients of FSW; MSM: men who have sex with men.

Scaling up HIVST may avert 20·5% (15·3-27·6%) of new HIV infections among MSM over 2019-2028 in Côte d’Ivoire, 27·3% (18·0-36·2%) in Mali, and 28·0% (21·3-37·5%) in Senegal, 2-5% new infections among FSW across countries, and 2-fold more among their clients due to higher increases in viral suppression among their female partners (Figures 3b, **S10**, **Table S6b**). Overall, 16·2% (10·0-23·1%) of all new HIV infections may be averted over 2019-2028 in Senegal, 5·3% (3·0-8·9%) in Mali and 1·6% (1·0-2·4%) in Senegal, Mali, and Côte d’Ivoire, respectively. In total, 2 400 (1 500-3 400), 1 700 (900-3 100), and 2 700 (1 600-4 500) new HIV infections may be averted over 2019-2028 in Côte d’Ivoire, Mali and Senegal, and ∼3-times more infections averted over 2019-2038 (Figure 3b). Overall, HIVST scale-up may avert <6·0% HIV deaths in Senegal, and <1·5% in Côte d’Ivoire and Mali over 10 years (Figure 3c, **Table S6c**), but it could avert a median of 13-18% of HIV-related deaths among MSM, 3-9% among FSW, and <1·5% among clients over 10 years across countries, and 2- to 3-fold more over 20 years (Figure 3c and **Table S6c**).

A substantial fraction of all averted infections over 2019-2038 would be among non-KP, 73·0% (58·4-83·4%) in Côte d’Ivoire, 71·6% (51·8-82·1%) in Mali, and 40·4% (28·5-52·1%) in Senegal, while 12-59% of HIV-related deaths averted would be among non-KP across countries (**Figures S11a,b**). Our sensitivity analysis, presented in supplement, showed a limited influence to variations in our assumptions on the estimated fraction of new HIV infections and deaths averted by a scale-up of HIVST among KP.

## Discussion

Our analysis suggest that the ATLAS program has had a sizeable impact on HIV acquisitions and deaths overall considering the fairly small number of kits distributed. ATLAS may have already resulted in hundreds of additional HIV diagnoses among KP over its course (2019-2021) and could avert around 4-9% of new HIV infections and deaths among MSM and 1-3% among FSWs across countries over a 20-year period. A national scale-up of HIVST among FSW and MSM may not only improve access to HIV services, reduce inequity and improve the lives of KP and their partners, but also have some epidemiological impact among the overall population in Western Africa. Reduction in current inequities in HIV response between KP and non-KP was exemplified by the substantial reduction in time from HIV acquisition to diagnosis across KPs in all settings, especially where diagnosis rates were sub-optimal initially.

To our knowledge, this is the first study evaluating the plausible population-level impact of HIVST in Western Africa, whilst several other modelling analyses have evaluated the impact (and cost-effectiveness) of HIVST in Southern Africa, a region with 10-fold higher overall HIV prevalence where KP acquire lower fractions of infections.^27,29,51^ Our study reveals important heterogeneities in the potential impact of an HIVST scale-up among KP across Western African countries, due to differences in HIV epidemic contexts and existing levels of HIV diagnosis. The lower overall impact of HIVST scale-up among KP in Côte d’Ivoire was due to the higher overall HIV prevalence among adults (2·3%) compared to Mali and Senegal (<1%), with many HIV infections occurring among KP partners and non-KP, and to larger diagnosis gaps than in Mali. This contrasts with the highest estimated overall impact of ATLAS in Senegal, where an increasing fraction of all PLHIV are MSM (16% in 2020 in our study), which had suboptimal use of HIV testing and benefitted from substantial HIVST distribution. Large overall HIVST impact on new HIV infections in Senegal suggests an important role of this population on the epidemic dynamics, which was already highlighted by recent analyses.^52,53^ Around half of averted HIV infections were estimated to have occurred among non-KP groups, despite receiving only a third of the kits. Increases in HIV diagnosis among KP following HIVST distribution averted HIV-related deaths by increasing life-expectancy due to subsequent increase in coverage of treatment and by averting HIV infections due to higher population-level viral load suppression. As a result, the high impact of HIVST on acquisitions among MSM was because increases in their coverage of viral suppression immediately translated to reductions in their risk of acquiring HIV (and not only the risk of transmitting it). In contrast, the impact among FSW of HIVST distribution was around half that among their clients because increasing ART coverage among FSW first need to reduce HIV risk among clients, before subsequently preventing HIV among FSW not living with HIV.

Our estimated time from HIV acquisition to diagnosis of 4 years overall in the ATLAS countries in the absence of HIVST was similar to recent Shiny90 estimates for Western Africa (5 years).^2^ However, our analysis revealed its large variations across countries and risk groups. It was almost 2 years longer in Mali compared to Senegal and Côte d’Ivoire, and 1·1-2·3 time longer among MSM than non-KP across countries (and especially non-KP women). These gaps in HIV diagnosis justified the need for alternative anonymous testing modalities for KP, whereas scale-up of HIVST among KP attenuated disparities in HIV diagnosis by decreasing the time from infection to diagnosis among MSM by up to 2-fold. However, ATLAS only reached clients of FSW through secondary distribution from FSW (**Table S4a**), despite clients being a much larger population, stressing the need to specific HIVST direct distribution strategies them.

Our study has some limitations and highlights current data gaps for these three West African countries, especially regarding clients of FSW, for which no population size and HIV prevalence estimates were available in Mali, and scarce data in the other ATLAS countries. This was addressed by representing wider uncertainty in levels of HIV prevalence and diagnosis/treatment for this population, and by using client sexual behaviour data from Côte d’Ivoire. The number of HIVST kits distributed during its scale-up were proportional to the size of the KPs, thus our estimated impact of HIVST did not depend on uncertainties in KP size estimates. In particular, our model predicted relatively few MSM in Senegal in 2020 (0·5%; 0·3-0·9%) of all adult males, corresponding to 21 000 (13 000-41 000) MSM), however this modelled population size was higher than two national estimates from 2012 and 2016 (of approximatively 10 000 MSM in the country).^12^ Larger fraction of MSM population in Senegal could not be reproduced by the model due to incompatibilities with 1) concomitant low HIV prevalence among all males and high prevalence among MSM, and 2) concomitant high ART coverage among all males and low ART coverage among MSM. ATLAS distribution data was available by distribution channel, sex, and age, whereas secondary distribution and linkage to care following a reactive self-test was characterised during two anonymous phone surveys, which participants may not accurately reflect the population receiving the tests.^11,47^ However our sensitivity analysis found a modest influence of these possible biases on our projected impacts. Our model might have slightly underestimated the impact of ATLAS because we did not include specific distribution channels focussing on partners of PLHIV, patients of STI clinics, and people who use drugs, which represented only 12% of the kits distributed. These populations are not necessarily well characterised in terms of size, HIV prevalence and diagnosis, and often overlap with the populations included in the model (e.g. clients of FSW may visit STI clinics).

Our analysis relied on comprehensive reviews of published and unpublished data characterising past and current state of the HIV epidemic and interventions in each country, including recent IBBS surveys, while integrating a comprehensive knowledge of HIVST use in countries through programmatic and survey data from the ATLAS program, and expertise from our country partners and collaborators. The number of the kits distributed within our scale-up scenario was based on WHO recommendations and similar to country-anticipated purchases over the coming years,^48^ which allowed to predict the most likely impact of HIVST distribution in each modelled group. The model fitting to a large variety of outcomes such HIV testing history or viral load suppression among PLHIV using a Bayesian framework accounting for uncertainty and possible biases in current levels of HIV diagnosis (often underestimated by KP surveys relying on self-reporting of diagnosis history),^54^ increased the robustness of our estimates. Our study translated increases in diagnosis coverage into likely impacts on onward transmissions, which can only be done using mathematical models. Finally, our sensitivity analysis further explored the impact of uncertainties in use of HIVST and linkage to care, and suggested that our estimated impacts where fairly robust to them.

In conclusion, HIVST hold promise to increase diagnosis coverage among KP and their partners, and our study shows that its scale-up could substantially reduce disparities in HIV diagnosis in Western Africa by reaching and improving health outcomes of hard-to-reach populations, especially male KP.^55^ Although more informed recommendations could be obtained from further cost-effectiveness analyses, the modest overall impact of HIVST scale-up on new HIV infections (especially in Côte d’Ivoire and Mali), suggests that distributing self-tests kits to male populations at higher risk of transmitting HIV, such as clients of FSW or men engaged in concurrent partnerships could improve the impact of HIVST distribution and accelerate the decrease in HIV incidence.

## Data Availability

All data produced in the present study are available upon reasonable request to the authors

## Declarations

## Acknowledgements

The current work was supported by UNITAID as part of the ATLAS consortium. The authors thank all the participants and the operational field worker in Côte d’Ivoire, Mali and Senegal.

## Research in context

### Evidence before this study

Around a third of people living with HIV in Western Africa remained undiagnosed in 2020, yet the extent to which HIV self-testing (HIVST) might improve HIV diagnosis and reduce new HIV infections and deaths in the region has not been evaluated. We searched Pubmed on June 28, 2023, with the terms ((HIV) AND (self-test*)) AND (Africa*), with no language restriction, and identified no studies on the population-level epidemiological impact of HIV self-testing in Western Africa.

### Added value of this study

Programmatic and survey data from the ATLAS programme, which distributed a total of around 380 000 HIVST kits to key populations such as female sex workers (FSW) and men who have sex with men (MSM) in Côte d’Ivoire, Mali and Senegal over 2019-2021 were combined with carefully calibrated mathematical models representing the detailed dynamics of HIV transmission, testing behaviours and treatment among different population groups in the three countries over time. We found that the ATLAS programme and subsequent scale-up in HIVST in Western African countries (where 95% of FSW and MSM would receive 2 self-tests kits each year) could have a sizeable impact on HIV diagnosis, new infections and mortality, especially among key populations and their partners.

### Implications of all the available evidence

HIVST distribution among key populations and their partners should be scaled-up in Western Africa – where the three quarters of new HIV infections occur in these populations – as it could have a substantial impact on new infections and mortality and help reduce inequities across population subgroups.

