## Supplement Material for "Assessing the potential population-level impacts of HIV self-testing distribution among key populations in Côte d’Ivoire, Mali, and Senegal: a mathematical modelling analysis"

**Table of contents**

### Contents

***Methods: model description*** ... ***3***

**Overview** ... **3**

**Model equations** ... **4**

Model initiation (1970) ... 4

Seeding of HIV in the population ... 6

Population recruitment of sexually naïve populations ... 6

Recruitment of sexually active population ... 7

Turnover in sex-work ... 12

Non-HIV mortality ... 12

Population ageing ... 12

HIV infection ... 14

HIV disease progression and mortality ... 25

HIV conventional testing, and diagnosis ... 26

HIV treatment initiation and drop-out ... 32

HIV self-testing ... 33

**Model fitting overview** ... **35**

**Model fitting data** ... **35**

Fitting data (Côte d'Ivoire) ... 35

Fitting data (Mali) ... 40

Fitting data (Senegal) ... 44

***Methods: simulated HIVST scenarios*** ... ***48***

***Results: model fits (Côte d'Ivoire)*** ... ***57***

**Demography** ... **57**

**HIV epidemiology** ... **59**

**HIV treatment cascade** ... **63**

***Results: model fits (Mali)*** ... ***69***

**Demography** ... **69**

**HIV epidemiology** ... **71**

**HIV treatment cascade** ... **74**

***Results: model fits (Senegal)*** ... ***80***

**Demography** ... **80**

**HIV epidemiology** ... **82**

**HIV treatment cascade** ... **85**

### Methods: model description

This model description was adapted from Maheu-Giroux et al. [1]

#### Overview

The deterministic compartmental model used for this analysis was adapted from a previously published model of HIV transmission in Côte d'Ivoire[1-3] and coded in C++. Our model was fitted to country-specific demographic, behavioural, HIV epidemiological and intervention data in Côte d'Ivoire, Mali and Senegal over 1980-2020.

The sexually active modelled population is noted $X_{i,u,s}^{r,a}(t)$, where

- The subscript "*r*" corresponds to gender-risk groups combinations: low-risk females (*r*=0), intermediate-risk females (*r*=1), female sex worker (FSW, *r*=2), low-risk males (*r*=3), intermediate-risk males (*r*=4), clients of FSW (*r*=5), men who have sex with men and women (MSMW) (*r*=6), and men who have sex with men exclusively (MSME) (*r*=7). See **Figure S1a**.
- The subscript "*a*" corresponds to age group: 15-19 years olds (*a*=0), 20-24 years olds (*a*=1), 25-49 years olds (*a*=2), and 50-59 years olds (*a*=3). See **Figure S1b**.
- The subscript "*i*" corresponds to HIV infection status: susceptible (*i*=0), acute infection (*i*=1), chronic infection with CD4>500 cells/µL (*i*=2), with CD4 between 350 and 500 cells/µL (*i*=3), with CD4 between 200 and 350 cells/µL (*i*=4), and with CD4 <200 cells/µL (*i*=5). See **Figure S1c.**
- The subscript "*u*" corresponds to HIV testing/diagnosis/treatment status: never tested (*u*=0), ever tested and undiagnosed if PLHIV (*u*=1), had a reactive self-test but is not diagnosed (*u*=2), diagnosed by conventional test and without having had a reactive self-test) (*u*=3), diagnosed via a confirmatory test following a reactive self-test (*u*=4), treated (*u*=5), ever treated but dropped-out from treatment (*u*=6). See **Figure S1d**.
- The subscript "s" corresponds to the population history of HIV self-testing: have never used self-test (*s*=0), have ever used self-tests (*s*=1). See **Figure S1d**.
- Our equations also use the subscript "*g*", which corresponds to the population sex, with females (*g*=0), and males (*g*=1).

The sexually naïve population is noted $V_r(t)$, with everyone being assumed to be aged 15-19 year old, HIV uninfected, and never having tested for HIV.

#### Model equations

The model can be expressed as a set of ordinary differential equation reflecting changes in modelled number of sexually naïve ($V_r$) and sexually active individuals ($X_{i,u,s}^{r,a}$).

$$\frac{dV_r}{dt} = L_r(t) - G'_r(t) - Y'_r(t)$$

$$\frac{dX_{i,u,s}^{r,a}}{dt} = E_{i,u,s}^{r,a}(t) + J_{i,u,s}^{r,a}(t) + Y_{i,u,s}^{r,a}(t) + G_{i,u,s}^{r,a}(t) + D_{i,u,s}^{r,a}(t) + Q_{i,u,s}^{r,a}(t) + T_{i,u,s}^{r,a}(t) + U_{i,u,s}^{r,a}(t) + S_{i,u,s}^{r,a}(t)$$

Where

- $L_r$ represents recruitment of sexually naïve populations into the model
- $E_{i,u,s}^{r,a}$ represents the recruitment of sexually active population through sexual debut of sexually naïve populations or migration of 25-49 years old adults
- $J_{i,u,s}^{r,a}$ represents the turnover in sex-work between female sex workers and intermediate-risk females
- $Y_{i,u,s}^{r,a}$ represents non-HIV mortality among sexually active, and $Y'_r$ among sexually naïve
- $G_{i,u,s}^{r,a}$ represents ageing among sexually active, and $G'_r$ among sexually naïve
- $D_{i,u,s}^{r,a}$ represents HIV acquisition
- $Q_{i,u,s}^{r,a}$ represents HIV infection progression and mortality
- $T_{i,u,s}^{r,a}$ represents HIV conventional testing and diagnosis
- $U_{i,u,s}^{r,a}$ represents HIV treatment (ART) initiation and drop-out
- $S_{i,u,s}^{r,a}$ represents HIV self-testing

Each of the components of the two equations above is described in the section that follows the characterization of the population at model initiation

##### Model initiation (1970)

The model starts in 1970 with a population of size $N_0$ (estimated by the United Nations Population Division (UNPD)) [4], and using the age-distribution for the year 1970. The initial population is assumed to be HIV uninfected (*i*=0) and having never tested for HIV (*u*=0, *s*=0).

The relative sizes of the KPs are constant over time from simulation start, and reflect estimates from country-specific surveys spanning over 1995-2020. As in [1], the risk-distribution of the sexually naïve population mirrors the one of the sexually active populations to keep the size of KP constant over time.

$X_{i,u,s}^{r,a}(1970) = 0$ if *i*=0 or *u*=0 or *s*=0

In 1970, the female population is distributed as follows:

1) Lower-risk females (*r*=0) of age *a* (sexually active and naïve):

$$\begin{cases} X_{0,0,0}^{0,a}(1970) = N_0\ PR_{Fem}PR_{A_a}(1-PR_{FSW})(1-PR_{IRF})\big(1-Vir_0(1970)\big) \\ V_0(1970) = N_0\ PR_{Fem}PR_{A_a})(1-PR_{FSW})(1-PR_{IRF})Vir_0(1970) \end{cases} \text{ if } a=0$$

$$X_{0,0,0}^{0,a} = N_0\ PR_{Fem}PR_{A_a}(1-PR_{FSW})(1-PR_{IRF}) \text{ if } a>0$$

2) Intermediate-risk females ($r$=1) of age $a$ (sexually active and naïve):

$$\begin{cases} X_{0,0,0}^{1,a}(1970) = N_0\ PR_{Fem}PR_{A_a}(1-PR_{FSW})PR_{IRF}\big(1-Vir_0(1970)\big) \\ V_1(1970) = N_0\ PR_{Fem}PR_{A_a}(1-PR_{FSW})PR_{IRF}Vir_0(1970) \end{cases} \text{ if } a=0$$

$$X_{0,0,0}^{1,a} = N_0\ PR_{Fem}PR_{A_a}(1-PR_{FSW})PR_{IRF} \text{ if } a>0$$

3) Female sex workers ($r$=2) of age $a$ (sexually active and naïve):

$$\begin{cases} X_{0,0,0}^{2,a}(1970) = N_0\ PR_{Fem}PR_{A_a}PR_{FSW}\big(1-Vir_0(1970)\big) \\ V_2(1970) = N_0\ PR_{Fem}PR_{A_a}PR_{FSW}Vir_0(1970) \end{cases} \text{ if } a=0$$

$$X_{0,0,0}^{2,a}(1970) = N_0\ PR_{Fem}PR_{A_a}PR_{FSW} \text{ if } a>0$$

Where $PR_{Fem}$ is the proportion of females in the model, $PR_{A_a}$ is the relative size of the age group $a$ at model start, $Vir_0(1970)$ the fraction of females aged 15-19 years old that are sexually naïve in 1970 (this fraction being assumed to be equal to the fraction in the first data point), $PR_{FSW}$ the fraction of FSW among all females, $PR_{IRF}$ the fraction of intermediate-risk females among all non-KP females. Empirical studies suggested slightly higher proportions of FSW among adult females in Côte d'Ivoire, (e.g. > 1% in [5,6]) compared to Mali and Senegal.

In 1970, the male population is distributed as follows:

4) Lower-risk males ($r$=3) of age $a$ (sexually active and naïve):

$$\begin{cases} X_{0,0,0}^{3,a}(1970) = N_0\ (1-PR_{Fem})PR_{A_a}(1-PR_{Cli})(1-PR_{MSM})(1-PR_{IRM})\big(1-Vir_1(1970)\big) \\ V_3(1970) = N_0\ (1-PR_{Fem})PR_{A_a}(1-PR_{Cli})(1-PR_{MSM})(1-PR_{IRM})Vir_1(1970) \end{cases} \text{ if } a=0$$

$$X_{0,0,0}^{3,a}(1970) = N_0\ (1-PR_{Fem})PR_{A_a}(1-PR_{Cli})(1-PR_{MSM})(1-PR_{IRM}) \text{ if } a>0$$

5) Intermediate-risk males ($r$=4) of age $a$ (sexually active and naïve):

$$\begin{cases} X_{0,0,0}^{4,a}(1970) = N_0\ (1-PR_{Fem})PR_{A_a}(1-PR_{Cli})(1-PR_{MSM})PR_{IRM}\big(1-Vir_1(1970)\big) \\ V_4(1970) = N_0\ (1-PR_{Fem})PR_{A_a}(1-PR_{Cli})(1-PR_{MSM})\ PR_{IRM}Vir_1(1970) \end{cases} \text{ if } a=0$$

$$X_{0,0,0}^{4,a}(1970) = N_0\ (1-PR_{Fem})PR_{A_a}(1-PR_{Cli})(1-PR_{MSM})PR_{IRM} \text{ if } a>0$$

6) Clients of FSW ($r$=5) of age $a$ (sexually active and naïve):

$$\begin{cases} X_{0,0,0}^{5,a}(1970) = N_0\,(1 - PR_{Fem})PR_{A_a}PR_{Cli}\big(1 - Vir_1(1970)\big) \\ V_5(1970) = N_0\,(1 - PR_{Fem})PR_{A_a}\,PR_{Cli}Vir_1(1970) \end{cases} \text{ if } a=0$$

$X_{0,0,0}^{5,a}(1970) = N_0\,(1 - PR_{Fem})PR_{A_a}PR_{Cli}$ if $a$>0

7) Men who have sex with men and women (MSMW, *r*=6) of age *a* (sexually active and naïve):

$$\begin{cases} X_{0,0,0}^{6,a}(1970) = N_0\,(1 - PR_{Fem})PR_{A_a}PR_{MSM}PR_{Bi}\big(1 - Vir_1(1970)\big) \\ V_6(1970) = N_0\,(1 - PR_{Fem})PR_{A_a}PR_{MSM}PR_{Bi}Vir_1(1970) \end{cases} \text{ if } a=0$$

$X_{0,0,0}^{6,a}(1970) = N_0\,(1 - PR_{Fem})PR_{A_a}PR_{MSM}PR_{Bi}$ if $a$>0

7) Men who have sex with men exclusively (MSME, *r*=7) of age *a* (sexually active and naïve):

$$\begin{cases} X_{0,0,0}^{7,a}(1970) = N_0\,(1 - PR_{Fem})PR_{A_a}PR_{MSM}(1 - PR_{Bi})\big(1 - Vir_1(1970)\big) \\ V_7(1970) = N_0\,(1 - PR_{Fem})PR_{A_a}PR_{MSM}(1 - PR_{Bi})Vir_1(1970) \end{cases} \text{ if } a=0$$

$X_{0,0,0}^{7,a}(1970) = N_0\,(1 - PR_{Fem})PR_{A_a}PR_{MSM}(1 - PR_{Bi})$ if $a$>0

Where $(1 - PR_{Fem})$ is the proportion of males in the model, $PR_{A_a}$ is the relative size of the age group $a$ at model start, $Vir_1(1970)$ the fraction of males aged 15-19 years old that are sexually naïve in 1970 (again, this fraction being assumed to be equal to the estimate from each country first data point), $PR_{IRM}$ the fraction of intermediate-risk males among all non-KP and non-client males, $PR_{Cli}$ the fraction of FSW clients among all males, $PR_{MSM}$ the fraction of MSM among all males, $PR_{Bi}$ the fraction of MSM that ever had a female partner.

The fraction $PR_{Cli}$ is calculated using the multiplier method[7], accounting for partner change rates reported by FSW and their clients, as well as the size of the FSW population[1]. As in [1,3], simulations were discarded when the estimated fraction $PR_{Cli}$ was over 20%.

#### Seeding of HIV in the population

HIV is assumed to start spreading into a very small fraction of the modelled population between 1975 and 1979 (using a parameter $HIV_{Start}$). At that particular time point, fractions $Prev_{FSW}$, $Prev_{Cli}$, and $Prev_{MSM}$ of FSW, clients and MSM respectively are assumed to become infected by HIV (and all with a CD4>500 cells/µL) (see **Table S1a**).

#### Population recruitment of sexually naïve populations $L_r$

Recruitment of sexually naïve populations of each risk group ($L_r$) is determined at each time step using the following formula:

$$L_r(t) = \left(Y'_r(t) + \sum\nolimits_{a,i,u,s} \mu_a X^{r,a}_{i,u,s}(t) + \sum\nolimits_{a,i>0,u\neq 5,s} \gamma_4 X^{r,a}_{i,u,s}(t) + \sum\nolimits_{a,i>0,s} \frac{\gamma_4}{RR_\omega} X^{r,a}_{i,5,s}(t) + \varepsilon'\left(\sum\nolimits_{a,i,u,s} X^{r,a}_{i,u,s}(t) + V_r(t)\right) + \sum\nolimits_{i,u,s} G^{r,3}_{i,u,s}(t)\right) Vir_g(t)$$

Where $Y'_r$ $(= \mu_0 V_r)$ and $\mu_a X^{r,a}_{i,u,s}$ are the number of sexually naïve and sexually active individuals exiting the model due to non-HIV mortality at each time step, $\gamma_4$ the rate at which PLHIV in the last stage of infection (<200 CD4 cells/µL) die from AIDS. The parameter $RR_\omega$ reflects the increase in survival among PLHIV on ART compared to PLHIV not on ART[8]. The term $\varepsilon'$ is the population growth due to fertility and is calculated as $\varepsilon' = \varepsilon - (\chi PR\_A_2)$, where $\varepsilon$ is the total population growth rate, $\chi$ the migration rate and $PR\_A_2$ the fraction of people aged 25-49 years old in the model. As suggested by census data for Côte d'Ivoire, the large majority of immigrants are aged between 25 and 49 years[9]. The term $G^{r,3}_{i,u,s}$ is the number of people leaving the model having reached the age of 60 years old. Finally, the time-dependant parameter $Vir_g(t)$ is the fraction of female or male entering the 15-19 years old age group as sexually naïve and was informed using data from the countries successive DHS's.

#### Recruitment of sexually active population $E^{r,a}_{i,u,s}$

Sexually active individuals of each risk group are recruited at each time step through ageing of sexually naïve 15-19 years old or migration of sexually active 25-49 years old (which are assumed to have the same HIV prevalence as adults in Côte d'Ivoire but are assumed to have never tested for HIV).

$$E^{r,a=0}_{i=0,u=0,s=0} = \left(Y'_r(t) + \sum\nolimits_{a,i,u,s} \mu_a X^{r,a}_{i,u,s}(t) + \sum\nolimits_{a,i>0,u\neq 5,s} \gamma_4 X^{r,a}_{i,u,s}(t) + \sum\nolimits_{a,i>0,s} \frac{\gamma_4}{RR_\omega} X^{r,a}_{i,5,s}(t) + \varepsilon'\left(\sum\nolimits_{a,i,u,s} X^{r,a}_{i,u,s}(t) + V_r(t)\right) + \sum\nolimits_{i,u,s} G^{r,3}_{i,u,s}(t)\right)\left(1 - Vir_g(t)\right)$$

$$E^{r,a=1}_{i=0,u=0,s=0} = Age_0 V_r(t)$$

$$E^{r,a=2}_{i,u=0,s=0} = \chi PR\_A_2 \sum\nolimits_{a,u,s} X^{r,a}_{i,u,s}(t)$$

Where $\chi$ is the migration rate and $PR\_A_2$ the fraction of people aged 25-49 years old in the model

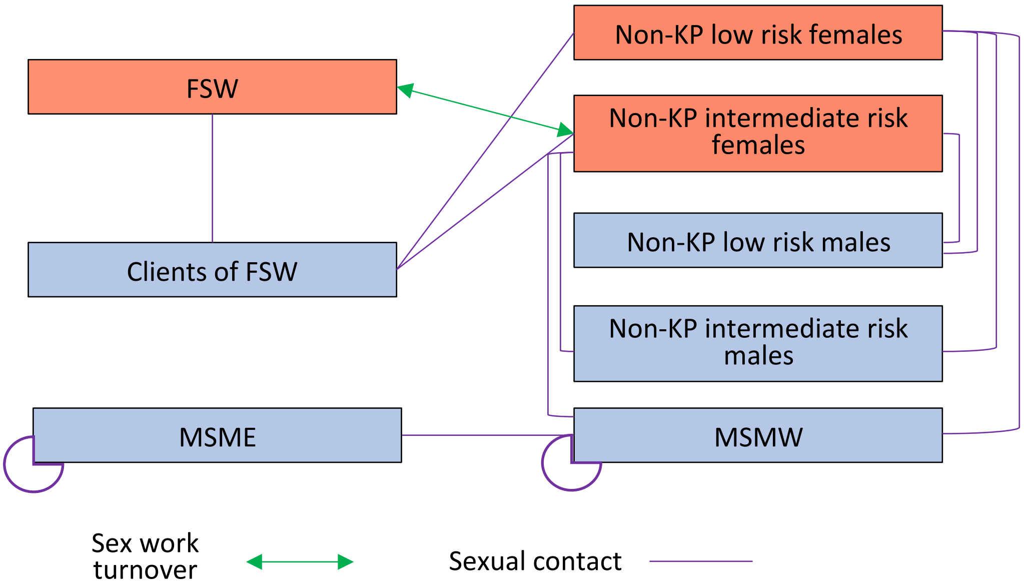

**Figure S1a:** modelled risk populations and their sexual contacts in the model, adapted from Maheu-Giroux et al. [1]. Female sex workers (FSW) only have sexual contacts with clients but can retire from sex work and move into the compartment of non-KP women with intermediate risk of infection. Clients of FSW have sexual contacts with all women risk groups. Non-KP women (low- and intermediate-risk women) have sexual contacts with all male risk groups, except MSME. The latter are assumed to form partnerships with other male exclusively, whereas MSMW form partnerships with both men and women.

| **Table S1a:** Model parameters related to demography and population structure | | | | | |
|---|---|---|---|---|---|
| **Population** | **Symbol** | **Value/ Prior distribution** | | | **References** |
| | | **Côte d'Ivoire** | **Mali** | **Senegal** | |
| **Demography** | | | | | |
| Total population aged 15-59 years in 1970 | $N_0$ | 2,583,135 | 3,152,660 | 2,176,151 | [4] |
| Population growth rate (year$^{-1}$) | $\varepsilon$ | 3.40% | 2.30% | 2.85% | [4] |
| Immigration rate of 25-49 years old (year$^{-1}$) | $\chi$ | U(0.0, 0.025) | U(0.0, 0.025) | U(0.0, 0.025) | [4] |
| Mortality rate (year$^{-1}$) | $\mu_{0:1}$<br>$\mu_2$<br>$\mu_3$ | U(0.0201, 0.0207)<br>U(0.0240, 0.0252)<br>U(0.0455, 0.0475) | U(0.0186, 0.0198)<br>U(0.0220, 0.0240)<br>U(0.0425, 0.0440) | U(0.0172, 0.0183)<br>U(0.0205, 0.0218)<br>U(0.0398, 0.0406) | (1/life expectancy at 15, 25, and 50 years) |
| Proportion of females in population | $PR_{Fem}$ | 47.3% | 51.1% | 51.8% | [4] (for the year 2000) |
| Age distribution (15-19, 20-24, 25-49, 50-59 years old) in 1970 | $PR_{A_0}$<br>$PR_{A_1}$<br>$PR_{A_2}$<br>$PR_{A_3}$ | 18.2%<br>14.7%<br>56.5%<br>13.2% | 19.7%<br>15.7%<br>51.4%<br>13.2% | 19.9%<br>15.8%<br>53.4%<br>10.9% | [4] (for the year 1970) |
| Rate of ageing in an older risk group or exiting the model at age 60 | $Age_0$<br>$Age_1$<br>$Age_2$<br>$Age_3$ | $Age_0 = \frac{1}{5};\ Age_1 = \frac{1}{5};\ Age_2 = \frac{1}{25};\ Age_3 = \frac{1}{10}$ | | | Based on the pre-defined modelled age groups (15-19, 20-24, 25-49, 50-59 years old) |
| **Population risk-structure** | | | | | |

| | | | | | |
|---|---|---|---|---|---|
| Proportion of sexually naïve among 15-19 years old females | $Vir_{g=0}(t)$ | Start = 27.4%<br>1994 = 27.4%<br>1999 = 35.9%<br>2004.5 = 34.2%<br>2012 = 35.3%<br>End = 35.3% | Start = 27.4%<br>1995.5 = 27.4%<br>2001 = 36.5%<br>2006 = 45.2%<br>2012.5 = 35.3%<br>2018 = 32.2%<br>End = 32.2% | Start = 64.8%<br>1992.5 = 64.8%<br>1997 = 66.0%<br>2005 = 71.2%<br>2011 = 72.1%<br>2013 = 74.8%<br>2014 = 72.9%<br>2015 = 74.8%<br>2016 = 74.4%<br>2017 = 74.1%<br>2018 = 75.8%<br>2019 = 77.2%<br>End = 77.2% | DHS surveys in Côte d'Ivoire[10-13], Mali[14-18], and Senegal[19-28]. Proportion is assumed to be constant from the last data point. |
| Proportion of sexually naïve among 15-19 years old males | $Vir_{g=1}(t)$ | Start = 44.3%<br>1994 = 44.3%<br>1999 = 44.3%<br>2004.5 = 48.9%<br>2012 = 57.3%<br>End = 57.3% | Start = 63.3%<br>1995.5 = 63.3%<br>2001 = 66.1%<br>2006 = 75.8%<br>2012.5 = 81.3%<br>2018 = 77.5%<br>End = 77.5% | Start = 69.0%<br>1992.5 = 69.0%<br>1997 = 69.0%<br>2005 = 69.0%<br>2011 = 80.9%<br>2013 = 89.0%<br>2014 = 89.0%<br>2015 = 86.5%<br>2016 = 87.8%<br>2017 = 84.6%<br>2018 = 89.3%<br>2019 = 92.4%<br>End = 92.4% | As above |
| Fraction of FSW among females | $PR_{FSW}$ | U(0.8, 2.1%) | U(0.4, 1.1%) | U(0.5, 0.9%) | Côte d'Ivoire: [5,6,29-31]<br>Mali: [6,32]<br>Senegal: [6,33,34] |
| Fraction of MSM among males | $PR_{MSM}$ | U(0.8, 1.7%) | U(0.2, 0.5%) | U(0.3, 1.2%) | Côte d'Ivoire: [35-38]<br>Mali: [6,32]<br>Senegal: [6,39] |
| Fraction of MSMW among all MSM | $PR_{Bi}$ | U(54.0, 75.5%) | U(53.5, 86.0%) | U(62.0, 85.0%) | Côte d'Ivoire: [38,40-46]<br>Mali: [45,47-49]<br>Senegal: [50-56] |
| Turnover of sex work (year$^{-1}$) | $tur$ | U(0.067, 0.2) | U(0.067, 0.2) | U(0.067, 0.2) | Assumed as in [1,3] |
| Fraction of intermediate-risk females (>1 partner/yr) among all non-KP females | $PR_{IRF}$ | U(5, 10%) | U(1, 5%) | U(1, 5%) | Côte d'Ivoire: [10-12,31,57]<br>Mali: [15-17,58]<br>Senegal: [21,22,24-27,59] |

| | | | | | |
|---|---|---|---|---|---|
| Fraction of intermediate-risk males (>2 partner/yr) among all non-KP males | $PR_{IRM}$ | U(5, 10%) | U(1, 5%) | U(1, 5%) | Côte d'Ivoire: [10-12,31,57]<br>Mali: [15-17,58]<br>Senegal: [21,22,24-27,59] |
| **HIV epidemic seeding** | | | | | |
| Year of epidemic start | $HIV_{Start}$ | U(1975, 1979) | U(1975, 1979) | U(1975, 1979) | Assumption |
| HIV prevalence among FSW at epidemic start | $Prev_{FSW}$ | U(0.1, 2%) | U(0.1, 2%) | U(0.1, 2%) | Assumption |
| HIV prevalence among FSW clients at epidemic start | $Prev_{Cli}$ | U(0.1, 1%) | U(0.1, 1%) | U(0.1, 1%) | Assumption |
| HIV prevalence among MSM clients at epidemic start | $Prev_{MSM}$ | U(0.1, 2%) | U(0.1, 2%) | U(0.1, 2%) | Assumption |

FSW: female sex workers; MSM: men who have sex with men; MSMW: men who have sex with men and women; U: uniform distribution (min, max)

#### Turnover in sex-work $J_{i,u,s}^{r,a}$

Female sex-workers ($r$=2) are assumed to start and cease sex work over their life course (see **Figure S1a** and **Table S1a**).

$J_{i,u,s}^{r,a}(t) = 0$ if r≠1 and r≠2

$J_{i,u,s}^{r=1,a}(t) = tur\, X_{i,u,s}^{r=2,a}(t) - tur'(t)\, X_{i,u,s}^{r=1,a}(t)$

$J_{i,u,s}^{r=2,a}(t) = tur'(t)\, X_{i,u,s}^{r=1,a}(t) - tur\, X_{i,u,s}^{r=2,a}(t)$

Where $tur$ is the rate at which FSW ($r$=2) cease forming commercial partnership and transition to the risk group of intermediate-risk females ($r$=1). This parameter varies across simulations but is fixed over time. Conversely, the parameter $tur'(t)$ represent the rate at which intermediate-risk females initiate sex work and transit into the compartment of FSW. This parameter is calculated at each time step so that the number of women initiating sex work is always equal to the number of women ceasing sex work.

$$tur'(t) = \frac{tur \sum_{r=2,a,i,u,s} X_{i,u,s}^{r,a}(t)}{\sum_{r=1,a,i,u,s} X_{i,u,s}^{r,a}(t)}$$

#### Non-HIV mortality $Y_{i,u,s}^{r,a}$

The rates of non-HIV mortality ($\mu_a$) vary by age group $a$ and are sourced from the United Nations Population Division (2019 revision of World Population Prospects) [4].

$Y_{i,u,s}^{r,a}(t) = -\mu_a X_{i,u,s}^{r,a}(t)$

Non-HIV mortality rates also apply to the sexually naïve population, with $Y'_r(t) = -\mu_0 V_r(t)$

#### Population ageing $G_{i,u,s}^{r,a}$

The rates of ageing of sexually active populations into older age groups (or to exit the model when reaching 60 years old) ($G_{i,u,s}^{r,a}$), are obtained as the inverse of the number of years covered by the age groups: $Age_0 = \frac{1}{5}$; $Age_1 = \frac{1}{5}$; $Age_2 = \frac{1}{25}$; $Age_3 = \frac{1}{10}$.

The term $G'_r$ correspond to the ageing of sexually naïve populations (all assumed to be 15-19 years old) age into a compartment of 20-24 years of sexually active.

$G'_r(t) = Age_0 V_r(t)$

Whereas,

$G_{i,u,s}^{r,0}(t) = -Age_0 X_{i,u,s}^{r,0}(t)$

$G_{i,u,s}^{r,1}(t) = Age_0 X_{i,u,s}^{r,0}(t) - Age_1 X_{i,u,s}^{r,0} + Age_0 V_r(t)$ if ($i$+$u$+s)=0 , and $G_{i,u,s}^{r,1}(t) = Age_0 X_{i,u,s}^{r,0}(t) - Age_1 X_{i,u,s}^{r,0}(t)$ otherwise

$G_{i,u,s}^{r,2}(t) = Age_1 X_{i,u,s}^{r,1}(t) - Age_2 X_{i,u,s}^{r,2}(t)$

$$G_{i,u,s}^{r,3}(t) = Age_2 X_{i,u,s}^{r,2}(t) - Age_3 X_{i,u,s}^{r,3}(t)$$

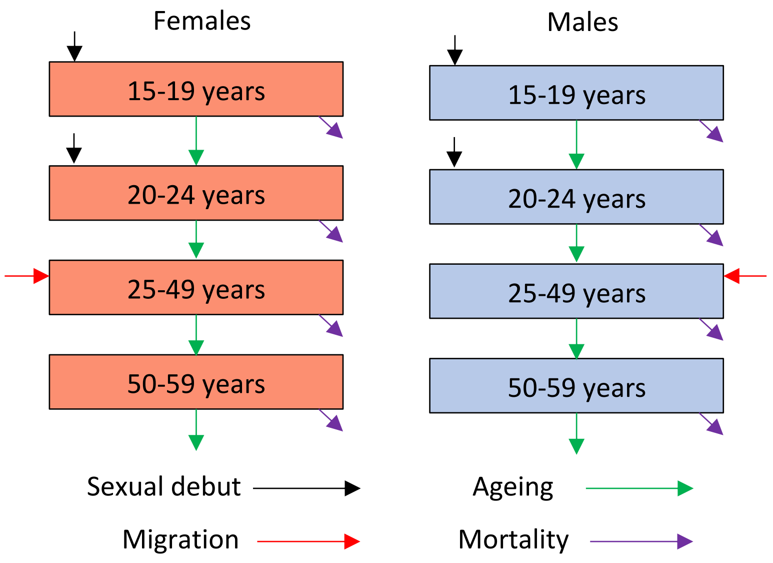

**Figure S1b:** modelled population age structure, migration, ageing and mortality, adapted from Maheu-Giroux et al. [1]

#### HIV infection $D_{i,u,s}^{r,a}$

The force of HIV infection, or rate at which people acquire HIV, vary over time and depend on individual and partner risk factors. We first describe how sexual mixing is represented in the model, then how the force of infection is derived.

##### Overall sexual mixing

As in [1], sexual mixing was modelled as a function of the gender, age, and risk group of individuals, and informed with data whenever possible. We define $p_{rar'a'}$ as the probability of a sexual contact between someone of risk group *r* and age *i* with someone of risk group *r'* and age *i'* , and estimated this quantity using the following equation:

$$p_{rir'i'} = WMW_{rr'}(M_{rr'})\Lambda_{raa'}$$

$$WMW_{rr'} = \begin{pmatrix} & F_{LR} & F_{IR} & F_{FSW} & M_{LR} & M_{IR} & M_{Cli} & M_{MSMW} & M_{MSME} \\ F_{LR} & 0 & 0 & 0 & 1 & 1 & 1 & 1 & 0 \\ F_{IR} & 0 & 0 & 0 & 1 & 1 & 1 & 1 & 0 \\ F_{FSW} & 0 & 0 & 0 & 0 & 0 & 1 & 0 & 0 \\ M_{LR} & 1 & 1 & 0 & 0 & 0 & 0 & 0 & 0 \\ M_{IR} & 1 & 1 & 0 & 0 & 0 & 0 & 0 & 0 \\ M_{Cli} & 1 & 1 & 1 & 0 & 0 & 0 & 0 & 0 \\ M_{MSMW} & 1 & 1 & 0 & 0 & 0 & 0 & 1 & 1 \\ M_{MSME} & 0 & 0 & 0 & 0 & 0 & 0 & 1 & 1 \end{pmatrix}$$

The binary matrix $WMW_{rr'}$ represent the types of partnerships allowed in the model for the risk groups of lower-risk females ($F_{LR}$), intermediate-risk female ($F_{IR}$), and female sex worker ($F_{FSW}$), as well as lower-risk males ($M_{LR}$), intermediate-risk males ($M_{IR}$), clients of female sex workers ($M_{Cli}$), men who have sex with men and women ($M_{MSMW}$), and men who have sex with men exclusively ($M_{MSME}$).

As in [1], the sizes of the low- and intermediate-risk groups were based on the number of sexual partners during the last 12 months in the countries DHS surveys ($F_{IR}$ defined as >1 partner per year; $M_{IR}$ defined as >2 partners per year; excluding those that reported selling or buying sex).

##### Risk-mixing

Sexual mixing by risk group was calculated using sexual behaviour data for low and intermediate-risk individuals. Due to data limitation, mixing by risk group was only available for couples living in the same household who both agreed to be interviewed and reported complete data on their sexual partners in Côte d'Ivoire, and was analysed from the female perspective. A DHS-reported matrix M was expanded to include the other risk groups and their associated parameters: $Bi_{Pref}$ being the fraction of partnerships that are with females for men having sex with men and women (MSMW), $Pr_{MSMW}$ the fraction of MSM also having sex with women (assuming proportional mixing between MSMW and MSME), and $Cli_{Mix}$ the fraction of partnerships that are with FSW for clients of FSW $Cli_{Mix} = \frac{c_5}{c_5+c_{5b}}$, where $c_5$ and $c_{5b}$ are the clients reported number of commercial partners and non-commercial partners, respectively.

$M_{rr'} =$

$$\begin{pmatrix} & F_{LR} & F_{IR} & F_{FSW} & M_{LR} & M_{IR} & M_{Cli} & M_{MSMW} & M_{MSME} \\ F_{LR} & 0 & 0 & 0 & 0.91 & \frac{0.09*M_{IR}}{M_{IR}+M_{Cli}+M_{MSMW}} & \frac{0.09*M_{Cli}}{M_{IR}+M_{Cli}+M_{MSMW}} & \frac{0.09*M_{MSMW}}{M_{IR}+M_{Cli}+M_{MSMW}} & 0 \\ F_{IR} & 0 & 0 & 0 & 0.88 & \frac{0.12*M_{IR}}{M_{IR}+M_{Cli}+M_{MSMW}} & \frac{0.12*M_{Cli}}{M_{IR}+M_{Cli}+M_{MSMW}} & \frac{0.12*M_{MSMW}}{M_{IR}+M_{Cli}+M_{MSMW}} & 0 \\ F_{FSW} & 0 & 0 & 0 & 0 & 0 & 1 & 0 & 0 \\ M_{LR} & 0.91 & 0.09 & 0 & 0 & 0 & 0 & 0 & 0 \\ M_{IR} & 0.88 & 0.12 & 1 & 0 & 0 & 0 & 0 & 0 \\ M_{Cli} & 0.88(1-Cli_{Mix}) & 0.12(1-Cli_{Mix}) & Cli_{Mix} & 0 & 0 & 0 & 0 & 0 \\ M_{MSMW} & 0.88(Bi_{Pref}) & 0.12(Bi_{Pref}) & 0 & 0 & 0 & 0 & Pr_{MSMW}(1-Bi_{Pref}) & (1-Pr_{MSMW})(1-Bi_{Pref}) \\ M_{MSME} & 0 & 0 & 0 & 0 & 0 & 0 & Pr_{MSMW} & 1-Pr_{MSMW} \end{pmatrix}$$

.

Mixing between risk groups was calculated using data from the 2011-2012 DHS in Côte d'Ivoire. See [1] for further details.

#### Age-mixing

Mixing patterns by age *(Λ)* were informed using data from the latest DHS surveys available for each country. This survey data reports the age of the most recent sexual partner among the population that was sexually active in the 12 months preceding the survey interview. The age-mixing matrices differ for males and females. As in [1], we assumed that age-mixing between FSW and their clients would correspond to that reported by males in the DHSs. For MSM, it was assumed that age mixing could range between completely assortative and proportional using the tuning parameter $MSM_{AgeMix}$, which was given a uniform distribution between 0 and 1.

For Côte d'Ivoire:

$$\Lambda_{raa'} = \begin{pmatrix} 0.130 & 0.130 & 0.719 & 0.021 \\ 0.130 & 0.130 & 0.719 & 0.021 \\ 0.003 & 0.003 & 0.753 & 0.241 \\ 0 & 0 & 0.187 & 0.813 \end{pmatrix} \text{ for } r=0,1$$

$$\Lambda_{raa'} = \begin{pmatrix} 0.483 & 0.483 & 0.035 & 0 \\ 0.483 & 0.483 & 0.035 & 0 \\ 0.194 & 0.194 & 0.609 & 0.004 \\ 0.125 & 0.125 & 0.187 & 0.167 \end{pmatrix} \text{ for } r=2:6$$

$$\Lambda_{raa'} = \left(1 - MSM_{AgeMix}\right) \begin{pmatrix} 1 & 0 & 0 & 0 \\ 0 & 1 & 0 & 0 \\ 0 & 0 & 1 & 0 \\ 0 & 0 & 0 & 1 \end{pmatrix} + MSM_{AgeMix} \begin{pmatrix} 0.182 & 0.147 & 0.565 & 0.106 \\ 0.182 & 0.147 & 0.565 & 0.106 \\ 0.182 & 0.147 & 0.565 & 0.106 \\ 0.182 & 0.147 & 0.565 & 0.106 \end{pmatrix} \text{ for } r=7:8$$

For Mali:

$$\Lambda_{raa'} = \begin{pmatrix} 0.076 & 0.076 & 0.820 & 0.028 \\ 0.076 & 0.076 & 0.820 & 0.028 \\ 0.001 & 0.001 & 0.674 & 0.324 \\ 0 & 0 & 0.112 & 0.888 \end{pmatrix} \text{ for } r=0,1$$

$$\Lambda_{raa'} = \begin{pmatrix} 0.496 & 0.496 & 0.008 & 0 \\ 0.496 & 0.496 & 0.008 & 0 \\ 0.175 & 0.175 & 0.645 & 0.004 \\ 0.024 & 0.024 & 0.876 & 0.077 \end{pmatrix} \text{ for } r\text{=2:6}$$

$$\Lambda_{raa'} = \left(1 - MSM_{AgeMix}\right)\begin{pmatrix} 1 & 0 & 0 & 0 \\ 0 & 1 & 0 & 0 \\ 0 & 0 & 1 & 0 \\ 0 & 0 & 0 & 1 \end{pmatrix} + MSM_{AgeMix}\begin{pmatrix} 0.197 & 0.157 & 0.514 & 0.132 \\ 0.197 & 0.157 & 0.514 & 0.132 \\ 0.197 & 0.157 & 0.514 & 0.132 \\ 0.197 & 0.157 & 0.514 & 0.132 \end{pmatrix} \text{ for } r\text{=7:8}$$

For Senegal:

$$\Lambda_{raa'} = \begin{pmatrix} 0.043 & 0.043 & 0.886 & 0.028 \\ 0.043 & 0.043 & 0.886 & 0.028 \\ 0.001 & 0.001 & 0.638 & 0.361 \\ 0 & 0 & 0.088 & 0.912 \end{pmatrix} \text{ for } r\text{=0,1}$$

$$\Lambda_{raa'} = \begin{pmatrix} 0.494 & 0.494 & 0.012 & 0 \\ 0.494 & 0.494 & 0.012 & 0 \\ 0.182 & 0.182 & 0.632 & 0.003 \\ 0.026 & 0.026 & 0.865 & 0.103 \end{pmatrix} \text{ for } r\text{=2:6}$$

$$\Lambda_{raa'} = \left(1 - MSM_{AgeMix}\right)\begin{pmatrix} 1 & 0 & 0 & 0 \\ 0 & 1 & 0 & 0 \\ 0 & 0 & 1 & 0 \\ 0 & 0 & 0 & 1 \end{pmatrix} + MSM_{AgeMix}\begin{pmatrix} 0.199 & 0.158 & 0.534 & 0.109 \\ 0.199 & 0.158 & 0.534 & 0.109 \\ 0.199 & 0.158 & 0.534 & 0.109 \\ 0.199 & 0.158 & 0.534 & 0.109 \end{pmatrix} \text{ for } r\text{=7:8}$$

As in [1], probabilities of sexual contacts are calculated separately for each partnership type *(rar'a')*. Imbalances between sexual partnerships demand and offer of the groups are likely (e.g. males typically reporting higher number of partners than females). The balance between supply of and demand for sexual partnership was obtained using a modified partner change rate *(*$c^*_{rar'a'}$*)* using the method described by Garnett and Anderson [60], and below:

$$\Delta_{rar'a'} = \frac{c_{r'a'} p_{r'a'ra} N_{r'a'}}{c_{ra} p_{rar'a'} N_{ra}}$$

$$c^*_{rar'a'} = c_{ra} \Delta^{\eta_k}_{rar'a'}$$

$$c^*_{r'a'ra} = c_{r'a'} \Delta^{-(1-\eta_k)}_{rar'a'}$$

Here, the parameter $\Delta_{rar'a'}$ measures the degree of imbalance between supply and demand for sexual partnerships of type *rar'a'*, while $\eta_k$ is the balance parameter which determines the degree to which partners alter their demand/offer of sexual partnerships. We assumed that clients of FSW would drive demand, whereas the balance parameter was assumed to be equal to 0.5 for MSM.

##### HIV force of infection

As in [1], we defined the force of infection (i.e. annual probability of HIV transmission) from an individual of risk group *r'* and age class *a'* to an individual of risk group *r* and age class *a* [61]. This probability partly depends on a "base" per-sex-act probability of transmission $\beta$ which has different values depending on the sex of both partners, with 1) female-to-male transmission probabilities ($= \beta_{fm}$) being lower than 2) male-to-female ($= \beta_{fm}RR_{\beta mf}$) and 3) male-to-male ($= \beta_{fm}RR_{\beta mm}$) transmission probabilities. Per-act transmission probabilities are further altered by a term of cofactors $Cof_{gg'a'}^{iu}$ (see next page and **Table S1c**). Since male circumcision is nearly ubiquitous in Western Africa, its protective effect was implicitly taken into account in the per-sex act transmission probabilities[62].

The matrix $\lambda_{rar'a'}(t)$ reflects the transmission probabilities for each partnership combination of category *r/r'* and age *a/a'*, where the susceptible partner is the one indexed by $ra$. The terms $g'$ and $g$ refers to the susceptible partner and infectious partner sex (=0 if female, =1 if male)

$$\lambda_{rar'a'}(t) = c^{*}_{rar'a'}p_{rar'a'}\left[\sum_{i,t}\left(\left(\frac{I^{iu}_{r'a'}(t)}{\sum_{i,t}I^{iu}_{r'a'}(t)+S_{r'a'}(t)}\right)\left(1-\left(\left(1-\beta Cof^{iu}_{gg'a'}(t)\right)^{\alpha_{rar'a'}(1-\nu_{ra}(t))}\left(1-\beta Cof^{iu}_{gg'a'}(t)(1-\varsigma)\right)^{\alpha_{rar'a'}(\nu_{ra}(t))}\right)\right)\right)\right]$$

Where $Cof_{gg'a'}^{iu}(t)$ represent a combination of cofactors for HIV acquisition (related to the susceptible individual gender $g'$ and age $a'$) and transmissions (related to the infectious individual disease stage $i$ and treatment status $u$). Here, the model accounts for the elevated risk of acquisition among young women ($RR_{\beta YF}$) (compared to older women) [63,64], the elevated risk of HIV transmission of individuals in the acute stage of HIV infection ($RR_{\beta Acute}$) [65]. We also assume that an increasing fraction of PLHIV on ART ($VLS_g(t)$) have a suppressed viral load and can't transmit HIV [66]. Estimates of this fraction are available over time and by sex from UNAIDS[67]. PLHIV on ART that don't have a suppressed viral load $\left(1-(VLS_g(t)\ VLSPtl)\right)$ are assumed to transmit HIV at the same rate as those not on ART.

The cofactor term $Cof_{gg'a'}^{iu}(t)$ is described below under three specific cases:

1) when both susceptible and infected partners don't have any specific risk factor for HIV acquisition/transmission (if $(g' = 1\ or\ a' > 1)\ and\ i > 1\ and\ u \neq 5$):

1.1. : $Cof_{gg'a'}^{iu}(t) = 1$

2) when the susceptible individual is not a young woman (i.e. if $g' = 1\ or\ a' > 1$) and the infected partner has a modified risk of HIV transmission because he/she is in the acute stage of infection or in on ART (i.e. if $i = 1\ or\ u = 5$):

2.1.: $Cof_{gg'a'}^{iu}(t) = RR_{\beta Acute}\ if\ i = 1\ and\ u \neq 5$

2.2.: $Cof_{gg'a'}^{iu}(t) = \left(1-\left(VLS_g(t)\ VLSPtl\right)\right)\ if\ i \neq 1\ and\ u = 5$

2.3.: $Cof_{gg'a'}^{iu}(t) = RR_{\beta Acute}\left(1-\left((VLS_g(t)\ VLSPtl\right)\right)\ if\ i = 1\ and\ u = 5$

3) when the susceptible individual is a young woman (i.e. if $g' = 0 \; and \; a' \leq 1$) and the infected partner has a modified risk of HIV transmission because it is in the acute stage of infection or in on ART (if $i = 1 \; or \; u = 5$):

2.1.: $Cof_{gg'a'}^{iu}(t) = RR_{\beta Acute} RR_{\beta YF} \; if \; i = 1 \; and \; u \neq 5$

2.2.: $Cof_{gg'a'}^{iu}(t) = \left(1 - \left(VLS_g(t) \; VLSPtl\right)\right) RR_{\beta YF} \; if \; i \neq 1 \; and \; u = 5$

2.3.: $Cof_{gg'a'}^{iu}(t) = RR_{\beta Acute} \left(1 - \left(VLS_g(t) \; VLSPtl\right)\right) RR_{\beta YF} \; if \; i = 1 \; and \; u = 5$

| **Table S1b:** Model parameters related to sexual behaviours | | | | | |
|---|---|---|---|---|---|
| **Parameter** | **Symbol** | **Prior distribution** | | | **References** |
| | | **Côte d'Ivoire** | **Mali** | **Senegal** | |
| **Number of sexual partners of sexually active risk groups** | | | | | |
| Lower-risk females | $c_0$ | U(0.8, 0.9) | U(0.79, 0.84) | U(0.63, 0.66) | Range selected as minimum and maximum values across DHS surveys in Côte d'Ivoire [10-13], Mali[14-18], and Senegal[19-28] |
| Intermediate-risk females | $c_1$ | U(2.4, 9.4) | U(2.0, 2.8) | U(2.0, 2.2) | As above |
| Lower-risk males | $c_3$ | U(1.0, 1.2) | U(0.86, 0.92) | U(0.61, 0.73) | As above |
| Intermediate-risk males | $c_4$ | U(4.7, 6.8) | U(4.2, 8.2) | U(3.2, 7.0) | As above |
| FSW | $c_2$ | U(216.0, 360.0) | U(200.0, 1007.0) | U(182.0, 273.0) | Studies among FSW in Côte d'Ivoire [68-71], Mali [72], and Senegal [73-76] |
| Clients of FSW with FSW | $c_5$ | U(23.0, 37.0) | U(23.0, 37.0) | U(23.0, 42.0) | Surveys in Côte d'Ivoire [77] and Senegal (personal communication of estimated from an unpublished IBBS client survey[78]). No data for Mali (used Côte d'Ivoire data) |
| Clients of FSW with non-KP females | $c_{5b}$ | U(1.0, 6.8) | U(1.0, 6.8) | U(2.5, 4.5) | Surveys in Côte d'Ivoire[68-71] and Senegal[78]. No data for Mali (used Côte d'Ivoire data) |
| MSMW | $c_6$ | U(1.0, 10.0) | U(1.0, 10.0) | U(1.0, 10.0) | Assumption |
| MSME | $c_7$ | U(1.0, 10.0) | U(1.0, 10.0) | U(1.0, 10.0) | Assumption |
| **Number of sex acts per partner-year** | | | | | |
| Lower-risk partners (r=0,3) | $\alpha_{rir'i'}$ | U(40.0, 48.0) | U(40.0, 48.0) | U(40.0, 48.0) | [10] |
| Intermediate-risk partners (r=1,4) | $\alpha_{rir'i'}$ | U(33.0, 66.0) | U(33.0, 66.0) | U(33.0, 66.0) | [10] |
| Clients-FSW partners (r=3,5) | $\alpha_{rir'i'}$ | U(1.0, 4.0) | U(1.0, 4.0) | U(1.0, 4.0) | [10] |
| MSM partners (r=6,7) | $\alpha_{rir'i'}$ | U(33.0, 66.0) | U(33.0, 66.0) | U(26.4, 39.6) | Data from MSM survey in Senegal [79]. Assumption for Côte d'Ivoire and Mali |
| Increase in numbers of sex acts of MSM from 2016 compared to before 2007 | $RR\alpha_{MSM2016}$ | No increase assumed | No increase assumed | U(1.5, 2.0) | Assuming a linear trend between 2007 [52] and 2016 [55] |
| Sexual balance parameter as per Garnett et al. 1994 [60] | $\eta$ | U(0, 1) | U(0, 1) | U(0, 1) | Assumption |

| | | | | | |
|---|---|---|---|---|---|
| Proportion of partnerships that are with females for MSMW | $Bi_{Pref}$ | U(32.0, 44.3%) | U(30.0, 45.0%) | U(34.7, 42.0%) | Surveys among MSM in Côte d'Ivoire [36] and Senegal [79]. Range for Mali expanded from Côte d'Ivoire and Senegal estimates (as no Mali data available) |
| Tuning parameter between assortative and proportional mixing by age among MSM | $MSM_{AgeMix}$ | U(0, 1) | U(0, 1) | U(0, 1) | Assumption |

FSW: female sex workers; MSM: men who have sex with men; MSMW: men who have sex with men and women; MSME: men who have sex with men exclusively; U: uniform distribution (min, max)

| **Table S1c:** Model parameters related to HIV infection and transmission (prior ranges assumed similar across countries) | | | |
|---|---|---|---|
| **Parameter** | **Symbol** | **Prior distribution** | **References** |
| **Natural history progression (PLHIV not on ART)** | | | |
| Average duration of acute infection (years) | $1/\gamma_0$ | U(0.11, 0.18) | [65] |
| Average time from seroconversion to 350 CD4 cells/μL | $1/\gamma_0 + 1/\gamma_1 + 1/\gamma_2$ | U(2.2, 4.6) | [80] |
| Average time from 350 CD4 to 200 CD4 cells/μL | $1/\gamma_3$ | U(3.9, 5.0) | [80] |
| Average time from 200 CD4 cells/μL to death | $1/\gamma_4$ | U(1.9, 3.9) | [80] |
| **HIV transmission** | | | |
| Female-to-male transmission probability per sex act | $\beta_{fm}$ | U(0.001, 0.017) | [81,82] |
| RR of HIV transmission from male to female compared to from female to male | $RR_{\beta mf}$ | U(1, 3) | [81] |
| RR of HIV transmission between males compared to from female to male | $RR_{\beta mm}$ | U(2, 6) | [83] |
| RR of HIV acquisition of females aged <25 years compared to females aged ≥25 years | $RR_{\beta YF}$ | U(1.25, 2.5) | [63,64] |
| Excess hazard-months of HIV transmission attributable to the acute stage | $EHM_{\beta Acute}$ | U(4.2, 16.8) | [65]. This parameter is used to calculate the RR of HIV transmission during acute HIV infection $RR_{\beta Acute}$ using the formula $RR_{\beta Acute} = \left(\frac{EHM_{\beta Acute}}{12/\gamma_0}\right) + 1$ |
| RR of HIV transmission among when a condom is used during a sex act (vs during a condomless sex act) | $\varsigma$ | U(0.75, 0.942) | [84] |
| **ART and viral suppression** | | | |
| ART initiation rate in the AIDS stage (<200 CD4 cells/μL) | $\rho_4$ | U(0.5, 4.0) | Assumption |
| Slope cofactor shaping linear relation between CD4 stages and ART initiation | $\varpi$ | U(0, 1.0) | Assumption |
| RR of ART initiation among diagnosed PLHIV in 2000 compared to 2020 | $RR_{\rho 2000}$ | U(0, 1.0) | Assumption |
| RR of ART initiation among KP compared to non-KP | $RR_{\rho KP}$ | U(0.2, 5.0) | Assumption |
| RR survival extension cofactor by HIV diagnosis/treatment status | $RR\omega_u$ | U(2.2, 6.3) if *u*=5, and 0 otherwise (only PLHIV on ART experience a reduced HIV mortality) | [8] |
| ART drop-out rate prior to 2015 | $\varphi$ | U(0.15, 0.27) | [85-91] |
| RR of ART drop-out for FSW and MSM (vs non-KP or clients) | $RR_{\varphi KP}$ | U(1.25, 1.75) | [92] (FSW data) |
| RR of ART drop-out after 2015 compared to before 2015 | $RR_{\varphi 2015p}$ | U(0.75, 1.00) | Assumption |

FSW: female sex workers; MSM: men who have sex with men; MSMW: men who have sex with men and women; MSME: men who have sex with men exclusively; RR: relative risk; U: uniform distribution (min, max)

| **Table S1d:** Model parameters related to condom use | | | | | |
|---|---|---|---|---|---|
| **Parameter** | **Symbol** | **Prior distribution** | | | **References** |
| | | **Côte d'Ivoire** | **Mali** | **Senegal** | |
| **During sex acts between non-KP groups** | | | | | |
| Among 15-24 years old | $Condom_{0:1}(t)$ | Start = (0-5%)<br>$YrCond_{NonKP}$= (3-5%)<br>1995 = (9.9-47.2%)<br>1999 = (11.5-55.7%)<br>2006 = (20.9-53.5%)<br>2012 = (20.7-59.5%)<br>End = (20.7-59.5%) | Start = (0-1%)<br>$YrCond_{NonKP}$= (1-2%)<br>2001 = (2.8-20.5%)<br>2006 = (20.6-30.5%)<br>2012 = (4.1-26.4%)<br>2018 = (2.5-20.7%)<br>End = (2.5-20.7%) | Start = (0-1%)<br>$YrCond_{NonKP}$= (1-2%)<br>2005 = (1.2-5.6%)<br>2011 = (0.0-1.8%)<br>2014 = (2.8-4.8%)<br>2016 = (1.7-3.8%)<br>2018 = (1.3-2.4%)<br>End = (1.3-2.4%) | [93,94] For estimates in the early 1980's. Range from DHS surveys in Côte d'Ivoire [10-13], Mali[14-18], and Senegal[19-28], using levels reported by females as minimum, and reported by males as maximum |
| Among 25-49 years old | $Condom_2(t)$ | Start = (0-2.5%)<br>$YrCond_{NonKP}$= (1-2.5%)<br>1995 = (2.5-21.7%)<br>1999 = (3.1-21.8%)<br>2006 = (4.7-23.8%)<br>2012 = (7.3-24.2%)<br>End = (7.3-24.2%) | Start = (0-1%)<br>$YrCond_{NonKP}$= (0.5-1%)<br>2001 = (0.1-4.1%)<br>2006 = (1.0-4.4%)<br>2012 = (1.4-4.5%)<br>2018 = (0.9-5.4%)<br>End = (0.9-5.4%) | Start = (0-1%)<br>$YrCond_{NonKP}$= (0.5-1%)<br>2005 = (1.2-4.6%)<br>2011 = (1.6-3.3%)<br>2014 = (1.5-2.0%)<br>2016 = (1.8-2.8%)<br>2018 = (0.5-1.5%)<br>End = (0.5-1.5%) | As above |
| Among 50-59 years old | $Condom_3(t)$ | Start = (0-2%)<br>$YrCond_{NonKP}$= (0.2-2%)<br>1995 = (0.2-9.7%)<br>1999 = (0.6-9.7%)<br>2006 = (0.6-10.0%)<br>2012 = (1.8-11.5%)<br>End = (1.8-11.5%) | Start = (0-1%)<br>$YrCond_{NonKP}$= (0.5-1%)<br>2001 = (0.1-4.1%)<br>2006 = (1.0-4.4%)<br>2012 = (1.4-4.5%)<br>2018 = (0.9-5.4%)<br>End = (0.9-5.4%) | Start = (0-1%)<br>$YrCond_{NonKP}$= (0.5-1%)<br>2005 = (1.2-4.6%)<br>2011 = (1.6-3.3%)<br>2014 = (1.5-2.0%)<br>2016 = (1.8-2.8%)<br>2018 = (0.5-1.5%)<br>End = (0.5-1.5%) | As above |
| **During sex acts of KP** | | | | | |

| | | | | | |
|---|---|---|---|---|---|
| All FSW with clients | $CondomSW(t)$ | Start = (0-5%)<br>$YrCond_{SW}$= (5-15%)<br>1991 = (57-68%)<br>1993 = (74-81%)<br>1995 = (73-88%)<br>1997 = (88-93%)<br>1998 = (88-98%)<br>2002 = (91-99%)<br>2007 = (90-99%)<br>2012 = (90-95%)<br>2014 = (85-93%)<br>End = (85-93%) | Start = (0-5%)<br>$YrCond_{SW}$= (5-15%)<br>1997 = (74-83%)<br>2000 = (90-98%)<br>2003 = (94-98%)<br>2009 = (94-98%)<br>2018 = (95-98%)<br>End = (95-98%) | Start = (0-5%)<br>$YrCond_{SW}$= (5-15%)<br>1990 = (79-89%)<br>2006 = (93-99%)<br>2010 = (90-98%)<br>2015 = (95-99%)<br>2019 = (88-96%)<br>End = (88-96%) | Surveys among FSW in Côte d'Ivoire[68,69,71,95], Mali[72,96-101] and Senegal[73-76,102] |
| All MSM | $CondomMSM(t)$ | Start = (0-0%)<br>$YrCond_{MSM}$= (5-15%)<br>2004 = (35-50%)<br>2012 = (57-69%)<br>2015 = (63-81%)<br>2017 = (68-82%)<br>End = (68-82%) | Start = (0-0%)<br>$YrCond_{MSM}$= (5-15%)<br>2014 = (70-82%)<br>2018 = (70-82%)<br>End = (70-82%) | Start = (0-0%)<br>$YrCond_{MSM}$= (5-15%)<br>2005 = (76-77%)<br>2014 = (73-76%)<br>2016 = (70-84%)<br>End = (70-84%) | Surveys among MSM in Côte d'Ivoire[36,40,50], Mali[32,103] and Senegal[52,55,78,104]. Estimates in Côte d'Ivoire and Senegal similar to levels of condom use reported by CohMSM participants (~75%) [45]. |
| **Year of increase in condom use in the 1980's** | | | | | |
| Non-KP groups (r=0,1,3,4) | $YrCond_{NonKP}$ | U(1981, 1990) | U(1981, 1990) | U(1981, 1990) | Assumption |
| FSW with clients (r=2,5) | $YrCond_{SW}$ | U(1981, 1990) | U(1981, 1990) | U(1981, 1990) | Assumption |
| MSM (r=6,7) | $YrCond_{MSM}$ | U(1981, 1990) | U(1981, 1990) | U(1981, 1990) | Assumption |
| **Scaling factors for the proportion of sex acts protected by condoms** | | | | | |
| Among non-KP aged 15-24 years | $CondPtl_{0:1}$ | U(0, 1) | U(0, 1) | U(0, 1) | Assumption |
| Among non-KP aged 25-49 years | $CondPtl_2$ | U(0, 1) | U(0, 1) | U(0, 1) | Assumption |
| Among non-KP aged 50-59 years | $CondPtl_3$ | U(0, 1) | U(0, 1) | U(0, 1) | Assumption |
| All FSW with clients | $CondSWPtl$ | U(0, 1) | U(0, 1) | U(0, 1) | Assumption |
| All MSM | $CondMSMPtl$ | U(0, 1) | U(0, 1) | U(0, 1) | Assumption |
| RR actual condom use during sex work (vs reported) | $RR_{CondSW}$ | U(0.7,1.0) | U(0.7,1.0) | U(0.7,1.0) | Conservative assumption based on studies among clients of FSW, or using biomarkers, pooling booth surveys or list randomisation[105-107] |

FSW: female sex workers; MSM: men who have sex with men; MSMW: men who have sex with men and women; MSME: men who have sex with men exclusively; RR: relative risk; U: uniform distribution (min, max)

#### HIV disease progression and mortality $Q_{i,u,s}^{r,a}$

At each time step, newly infected PLHIV progress through different stages of infections according to their CD4 cell counts, expressed by the superscript "*i*", (with *i*=0 corresponding to HIV-uninfected people, **Figure S1c**). The first stage of HIV infection (*i*=1) corresponds to acute infection, the second (*i*=2) to CD4>500 cells/µL, the third (*i*=3) for CD4 between 350 and 500 cells/µL, the fourth (*i*=4) for CD4 between 200 and 350 cells/µL, and the last one (*i*=5) for CD4 <200 cells/µL, the latter stage being associated with HIV-related mortality. Transitions rates between infection stages ($\gamma_i$) were sourced from the literature and shown in **Table S1c**. As in [1], the reduced HIV-mortality among PLHIV on ART was obtained by reducing the transition rates $\gamma_3$ and $\gamma_4$ by a factor $RR\omega_u$ (informed by mortality data) which is > 1 only when *u*=5.

For HIV-uninfected people: $Q_{i=0,u,s}^{r,a}(t) = 0$

For PLHIV:

$Q_{i=1,u,s}^{r,a}(t) = -\gamma_0 X_{i=1,u,s}^{r,a}(t)$, if in the acute infection stage

$Q_{i=2,u,s}^{r,a}(t) = \gamma_0 X_{i=1,u,s}^{r,a}(t) - \gamma_1 X_{i=2,u,s}^{r,a}(t)$, if CD4>500 cells/µL

$Q_{i=3,u,s}^{r,a}(t) = \gamma_1 X_{i=2,u,s}^{r,a}(t) - \gamma_2 X_{i=3,u,s}^{r,a}(t)$, if CD4 between 350 and 500 cells/µL

$Q_{i=4,u,s}^{r,a}(t) = \gamma_2 X_{i=3,u,s}^{r,a}(t) - (\frac{\gamma_3}{RR\omega_u}) X_{i=4,u,s}^{r,a}(t)$, if CD4 between 200 and 350 cells/µL

$Q_{i=5,u,s}^{r,a}(t) = (\frac{\gamma_3}{RR\omega_u}) X_{i=4,u,s}^{r,a}(t) - (\frac{\gamma_4}{RR\omega_u}) X_{i=5,u,s}^{r,a}(t)$, if CD4<200 cells/µL

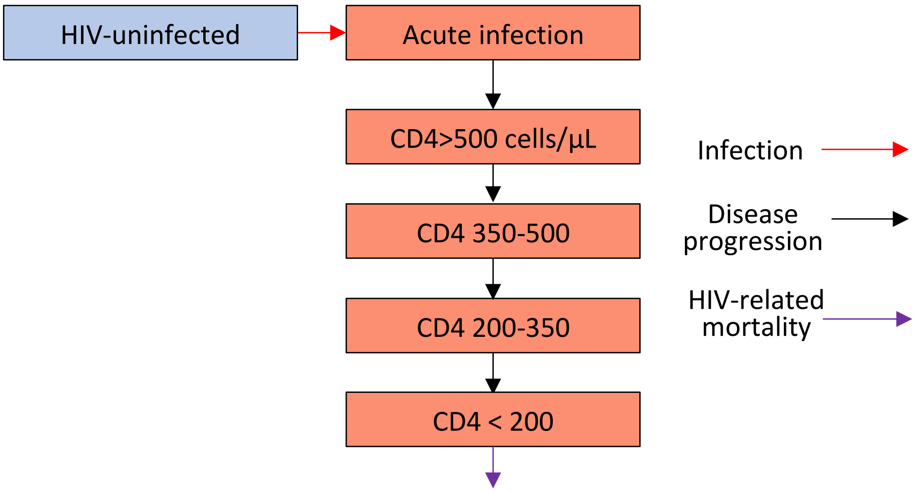

**Figure S1c:** modelled HIV infection stages among people not on ART.

##### HIV conventional testing, and diagnosis $T_{i,u,s}^{r,a}$

The rates of conventional HIV testing and diagnosis in the modelled populations vary over time and are informed by the trends in proportions of people having tested for HIV in the last year (parameter $\tau^{g,a}(t)$) (see **Table S2e**). This data was available by sex and age group from countries successive DHS surveys. As in[1], in order to replicate the trends in testing rates and maintain temporal consistency despite uncertainties in exact levels of testing, the parameter $TestPtl$ is used to represent the percentile of each data uncertainty range, and is sampled between 0 and 1 within each simulation. The variable $Year_\tau$ is the time where people start testing for HIV in each country, and is sampled in each simulation.

$T_{i,u,s}^{r,a}(t) = 0$ if time $\leq Year_\tau$

When time > $Year_\tau$, population flows will depend on population HIV and diagnosis/treatment status.

Among HIV-uninfected populations ($i$=0):

$T_{i,u=0,s}^{r,a}(t) = -X_{i,u=0,s}^{r,a}\,(t)\,\tau^{g,a}(t)\,RRTest_{i,u=0}^{r}(t)\,K_g$

$T_{i,u=1,s=0}^{r,a}(t) = 0$ (conventional HIV tests by this population are accounted for, but do not correspond to a flow into another population group)

$T_{i,u=1,s=1}^{r,a}(t) = X_{i,u=2,s=1}^{r,a}\,(t)\,\tau^{g,a}(t)\,RRTest_{i,u=2}^{r}(t)\,K_g$ (conventional HIV tests following false-reactive self-tests)

$T_{i,u=2,s=1}^{r,a}(t) = -X_{i,u=2,s=1}^{r,a}\,(t)\,\tau^{g,a}(t)\,RRTest_{i,u=2}^{r}(t)\,K_g$ (as above)

Among PLHIV ($i$ >0):

$T_{i,u=0,s}^{r,a}(t) = -X_{i,u=0,s}^{r,a}\,(t)\,\tau^{g,a}(t)\,RRTest_{i,u=0}^{r}(t)\,K_g$

$T_{i,u=1,s=0}^{r,a}(t) = -X_{i,u=1,s=0}^{r,a}\,(t)\,\tau^{g,a}(t)\,RRTest_{i,u=1}^{r}(t)\,K_g$

$T_{i,u=1,s=1}^{r,a}(t) = -X_{i,u=1,s=1}^{r,a}\,(t)\,\tau^{g,a}(t)\,RRTest_{i,u=1}^{r}(t)\,K_g$

$T_{i,u=2,s=1}^{r,a}(t) = -X_{i,u,s}^{r,a}\,(t)\,\tau_{ST}$ (confirmation of reactive HIV self-tests)

$$T_{i,u=3,s=0}^{r,a}(t) = \Big(\big(X_{i,u=0,s}^{r,a}\,(t)\,\tau^{g,a}(t)\,RRTest_{i,u=0}^{r}(t)\big) + \big(X_{i,u=1,s=0}^{r,a}\,(t)\,\tau^{g,a}(t)\,RRTest_{i,u=1}^{r}(t)\big)\Big)\,K_g$$

$T_{i,u=3,s=1}^{r,a}(t) = X_{i,u=1,s=1}^{r,a}\,(t)\,\tau^{g,a}(t)\,RRTest_{i,u=1}^{r}(t)\,K_g$

$T_{i,u=4,s}^{r,a}(t) = X_{i,u=2,s=1}^{r,a}\,(t)\,\tau^{g,a}(t)\,RRTest_{i,u=2}^{r}(t)\,K_g$

$T_{i,u\geq5,s}^{r,a}(t) = 0$ (conventional HIV tests by PLHIV on ART or that have dropped-out from ART are accounted for, but do not correspond to a flow into another population group)

Where the parameter $RRTest_{i,u}^{r}(t)$ is a product of cofactors (relative risks) defined using wide uncertainty ranges which aimed at reproducing empirical heterogeneities in HIV testing coverage by risk group, HIV status and HIV testing history status (see **Table S1e**):

- The parameter $RR_{\tau AIDS}$ reflects the increase in HIV conventional testing among PLHIV in the AIDS stage of infection (compared to PLHIV not in the AIDS stage).
- The parameters $RR_{\tau FSWStart}$ and $RR_{\tau FSW2020}$ represent the elevated HIV conventional testing rates among FSW compared to non-FSW females at the time points $Year_{\tau}$ and 2020, with the RR for a specific time being calculated assuming a linear trend between $RR_{\tau FSWStart}$ and $RR_{\tau FSW2020}$ over the period [$Year_{\tau}$ – 2020]. A similar assumption is made for the HIV testing rates of MSM (compared to non-MSM males), using the parameters $RR_{\tau MSMStart}$ and $RR_{\tau MSM2020}$.
- Heterogeneities in HIV conventional testing rates among PLHIV vs HIV-uninfected populations are represented using one parameter for KPs ($RR_{\tau PLHIV\_KP}$) and one for non-KPs ($RR_{\tau PLHIV\_NKP}$), and similarly for populations never having tested for HIV (vs ever testing, including HIV self-tests) (using the parameters $RR_{\tau NTest\_KP}$ and $RR_{\tau NTest\_NKP}$).
- Possible heterogeneities in HIV conventional testing among diagnosed PLHIV (compared to undiagnosed PLHIV) are captured using the parameter $RR_{\tau Diagn}$.

An overall $K_g$ parameter is used as an overall fudge factor to fit the model to history of HIV testing (by risk group, and HIV status when available) as well as number of conventional HIV tests done in the countries.

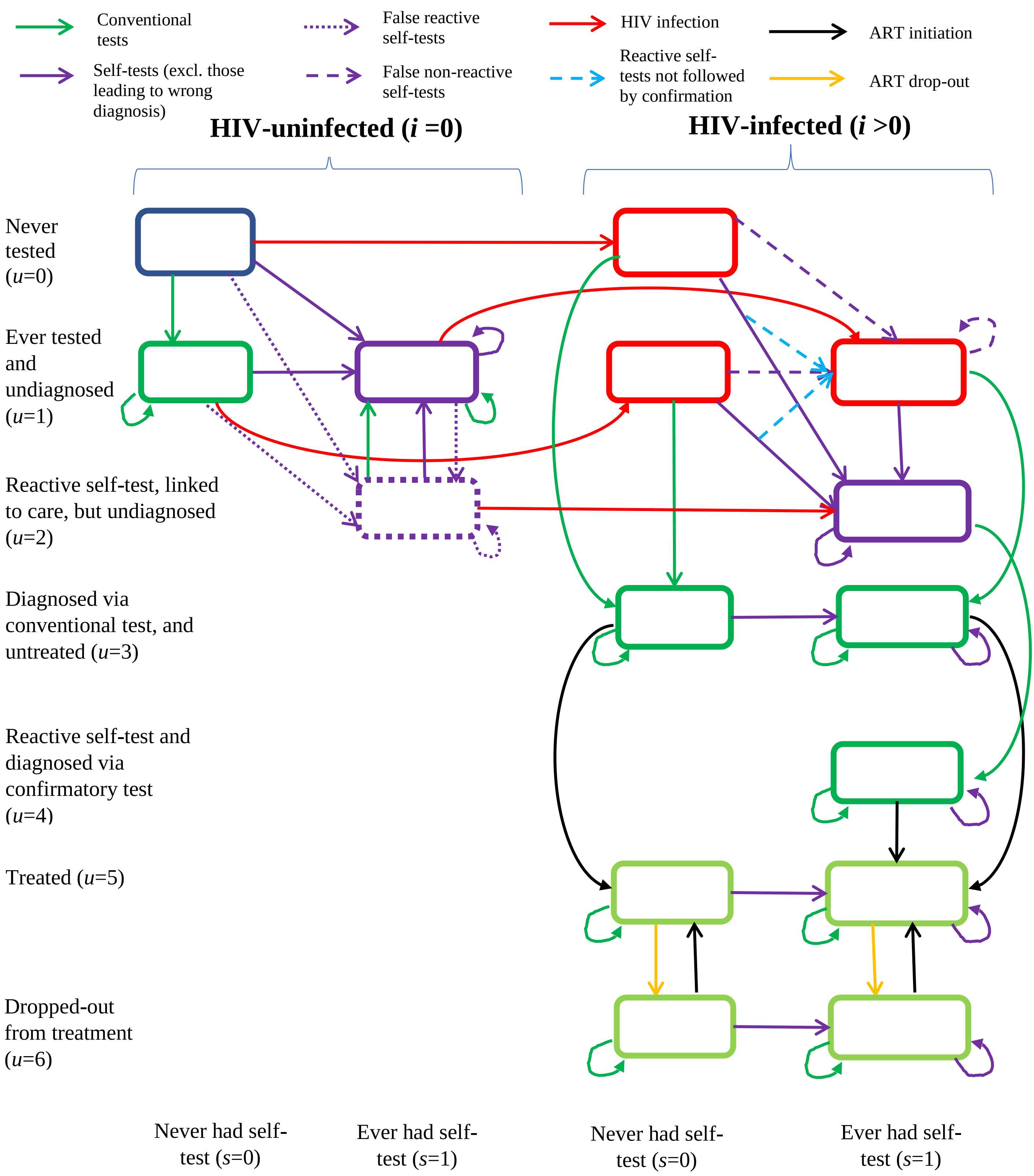

**Figure S1d:** modelled stages of HIV testing, diagnosis and treatment.

| **Table S1e:** Model parameters related to HIV testing and treatment | | | | | |
|---|---|---|---|---|---|
| **Parameter** | **Symbol** | **Prior distribution** | | | **References** |
| | | **Côte d'Ivoire** | **Mali** | **Senegal** | |
| **HIV conventional testing probabilities (last 12 months) $\tau^{g,a}(t)$** | | | | | |
| Start year of HIV testing | $Year_\tau$ | U(1996, 1999) | U(1996, 1999) | U(1996, 1999) | Assumed |
| Non-FSW females aged 15-24 years | $\tau^{0,0:1}(t)$ | Start = (0, 0%)<br>$Year_\tau$ = (0, 0%)<br>2005 = (1, 6%)<br>2011 = (7, 26%)<br>2017 = (8, 33%)<br>End = (8, 33%) | Start = (0, 0%)<br>$Year_\tau$ = (0, 0%)<br>2006 = (2, 7%)<br>2013 = (3, 12%)<br>2018 = (4, 15%)<br>End = (4, 15%) | Start = (, 0%)<br>$Year_\tau$ = (0, 0%)<br>2005 = (0, 2%)<br>2011 = (5, 25%)<br>2014 = (7, 28%)<br>End = (7, 28%) | DHS surveys in Côte d'Ivoire [10-13], Mali[14-18], and Senegal[19-22,28] |
| Non-FSW females aged 25-49 years | $\tau^{0,2}(t)$ | Start = (0, 0%)<br>$Year_\tau$ = (0, 0%)<br>2005 = (2, 9%)<br>2011 = (7, 26%)<br>2017 = (11, 45%)<br>End = (11, 45%) | Start = (0, 0%)<br>$Year_\tau$ = (0, 0%)<br>2006 = (1, 5%)<br>2013 = (3, 12%)<br>2018 = (5, 18%)<br>End = (5, 18%) | Start = (0, 0%)<br>$Year_\tau$ = (0, 0%)<br>2005 = (0, 1%)<br>2011 = (7, 28%)<br>2014 = (8, 33%)<br>End = (8, 33%) | As above |
| Non-FSW females aged 50-59 years | $\tau^{0,3}(t)$ | Start = (0, 0%)<br>$Year_\tau$ = (0, 0%)<br>2005 = (2, 9%)<br>2011 = (7, 26%)<br>2017 = (11, 45%)<br>End = (11, 45%) | Start = (0, 0%)<br>$Year_\tau$ = (0, 0%)<br>2006 = (1, 3%)<br>2013 = (2, 4%)<br>2018 = (3, 12%)<br>End = (3, 12%) | Start = (0, 0%)<br>$Year_\tau$ = (0, 0%)<br>2005 = (0, 2%)<br>2011 = (4, 17%)<br>2014 = (6, 25%)<br>End = (6, 25%) | As above |
| Non-MSM males aged 15-24 years | $\tau^{1,0:1}(t)$ | Start = (0, 0%)<br>$Year_\tau$ = (0, 0%)<br>2005 = (1, 4%)<br>2011 = (4, 15%)<br>2017 = (3, 11%)<br>End = (3, 11%) | Start = (0, 0%)<br>$Year_\tau$ = (0, 0%)<br>2006 = (1, 4%)<br>2013 = (2, 7%)<br>2018 = (1, 4%)<br>End = (1, 4%) | Start = (0, 0%)<br>$Year_\tau$ = (0, 0%)<br>2005 = (0, 2%)<br>2011 = (3, 15%)<br>2014 = (3, 15%)<br>End = (3, 15%) | As above |
| Non-MSM males aged 25-49 years | $\tau^{1,2}(t)$ | Start = (0, 0%)<br>$Year_\tau$ = (0, 0%)<br>2005 = (2, 8%)<br>2011 = (5, 22%)<br>2017 = (7, 28%)<br>End = (7, 28%) | Start = (0, 0%)<br>$Year_\tau$ = (0, 0%)<br>2006 = (2, 10%)<br>2013 = (4, 15%)<br>2018 = (3, 12%)<br>End = (3, 12%) | Start = (0, 0%)<br>$Year_\tau$ = (0, 0%)<br>2005 = (1, 4%)<br>2011 = (5, 20%)<br>2014 = (5, 20%)<br>End = (5, 20%) | As above |

| | | | | | |
|---|---|---|---|---|---|
| Non-MSM males aged 50-59 years | $\tau^{1,3}(t)$ | Start = (0, 0%)<br>$Year_\tau$ = (0, 0%)<br>2005 = (1, 3%)<br>2011 = (5, 22%)<br>2017 = (4, 16%)<br>End = (4, 16%) | Start = (0, 0%)<br>$Year_\tau$ = (0, 0%)<br>2006 = (2, 10%)<br>2013 = (4, 15%)<br>2018 = (3, 12%)<br>End = (3, 12%) | Start = (0, 0%)<br>$Year_\tau$ = (0, 0%)<br>2005 = (1, 4%)<br>2011 = (4, 17%)<br>2014 = (3, 15%)<br>End = (3, 15%) | As above |
| Scaling factor for HIV testing among non-FSW and non-MSM | $TestPtl$ | U(0, 1) | U(0, 1) | U(0, 1) | Assumed |
| Overall HIV testing scaling factor (by gender) | $K_g$ | U(0.2, 5) | U(0.2, 5) | U(0.2, 5) | Assumed |
| **Relative change in conventional testing rate among populations** | | | | | |
| RR HIV testing among PLHIV in AIDS stage (vs PLHIV not in the AIDS stage) | $RR_{\tau AIDS}$ | U(1, 8) | U(1, 8) | U(1, 8) | Assumption similar to [1] |
| RR HIV testing among FSW (vs non-KP females) at $Year_\tau$ | $RR_{\tau FSWStart}$ | U(1, 10) | U(1, 10) | U(1, 10) | Assumption based on higher reported history of testing among FSW compared to non-FSW women |
| RR HIV testing among FSW (vs non-KP females) from 2020 | $RR_{\tau FSW2020}$ | U(1, 10) | U(1, 10) | U(1, 10) | As above |
| RR HIV testing among MSM (vs non-MSM males) at $Year_\tau$ | $RR_{\tau MSMStart}$ | U(1, 10) | U(1, 10) | U(1, 10) | Assumption based on higher reported history of testing among MSM compared to non-MSM men |
| RR HIV testing among MSM (vs non-MSM males) from 2020 | $RR_{\tau MSM2020}$ | U(1, 10) | U(1, 10) | U(1, 10) | As above |
| RR HIV testing among KP PLHIV (vs HIV-uninfected KP) | $RR_{\tau PLHIV\_KP}$ | U(0.2, 5) | U(0.2, 5) | U(0.2, 5) | Assumed |
| RR HIV testing among non-KP PLHIV (vs HIV-uninfected non-KP) | $RR_{\tau PLHIV\_NKP}$ | U(0.2, 5) | U(0.2, 5) | U(0.2, 5) | Assumed |
| RR HIV testing among KP never having tested (vs KP ever tested) | $RR_{\tau NTest\_KP}$ | U(0.2, 5) | U(0.2, 5) | U(0.2, 5) | Assumed |
| RR HIV testing among non-KP never having tested (vs non-KP ever tested) | $RR_{\tau NTest\_NKP}$ | U(0.2, 5) | U(0.2, 5) | U(0.2, 5) | Assumed |
| RR HIV testing among diagnosed PLHIV (vs undiagnosed) | $RR_{\tau Diagn}$ | U(0.2, 3) | U(0.2, 3) | U(0.2, 3) | Assumed |
| **HIV self-testing** | | | | | |
| Rate of HIV self-testing | $\upsilon_{i,u,s}^{r,a}(t)$ | | | | Calculated from the number of kits distributed and used, see specific section |

| | | | | | |
|---|---|---|---|---|---|
| Fraction of HIV self-tests replacing conventional tests | $Repl$ | 20% | 30% | 40% | From an analysis of ATLAS and subnational programmatic data in Côte d'Ivoire and Senegal[108]. Mali assumed as the average between the estimates for Côte d'Ivoire and Senegal |
| HIV self-test sensitivity and specificity | $STSe_i$ and $STSp$ | | $STSe_i = 0.92$ if i>1<br>$STSe_i = 0$ if i=1<br><br>$STSp = 0.99$ | | Assumptions of the base-case scenario, based on manufacturer data[109]. We assumed that tests cannot detect HIV when the individual is in the acute stage of infection |
| Fraction of HIV self-tests which are followed by a confirmation test and linkage to care | $STLnk$ | | $STLnk = 0.5$ | | ATLAS phone survey data (2nd stage)[110] |
| Rate of HIV confirmatory testing among having had a reactive test and linked to care | $\tau_{ST}$ | | $\tau_{ST} = 6$ | | As above |
| Rate of ART initiation among those diagnosed via a confirmed reactive test | $\rho_{ST}$ | | $\rho_{ST} = 12$ | | As above |
| **Viral suppression among PLHIV** | | | | | |
| Fraction of female PLHIV on ART that have a suppressed viral load | $VLS_{g=0}(t)$ | Start = (40-60%)<br>2000 = (40-60%)<br>2018 = (70-89%)<br>2020 = (73-93%)<br>2030 = (85-95%)<br>End = (85-95%) | Start = (40-60%)<br>2000 = (40-60%)<br>2020 = (63-83%)<br>2030 = (85-95%)<br>End = (85-95%) | Start = (40-60%)<br>2000 = (40-60%)<br>2016 = (73-91%)<br>2020 = (77-97%)<br>2030 = (85-97%)<br>End = (85-97%) | From[67] for 2016 and 2020, assumptions otherwise |
| Fraction of male PLHIV on ART that have a suppressed viral load | $VLS_{g=1}(t)$ | Start = (40-60%)<br>2000 = (40-60%)<br>2018 = (69-91%)<br>2020 = (75-97%)<br>2030 = (85-97%)<br>End = (85-97%) | Start = (40-60%)<br>2000 = (40-60%)<br>2020 = (65-87%)<br>2030 = (85-95%)<br>End = (85-97%) | Start = (40-60%)<br>2000 = (40-60%)<br>2016 = (66-83%)<br>2020 = (76-96%)<br>2030 = (85-96%)<br>End = (85-96%) | As above |
| Scaling factor for viral suppression among PLHIV on ART | $VLSPtl$ | U(0, 1) | U(0, 1) | U(0, 1) | Assumption |

FSW: female sex workers; MSM: men who have sex with men; MSMW: men who have sex with men and women; MSME: men who have sex with men exclusively; RR: Relative risk; U: uniform distribution (min, max)

#### HIV treatment initiation and drop-out $U_{i,u,s}^{r,a}$

Our model reused the approach from[1,3] to represent ART initiation, which accounts for changes in ART eligibility, while reflecting the fact that CD4 cell counts may have been widely available in the region[90]. Our model assumes that individuals could only initiate ART after being diagnosed, hence no treatment was given until the period 1980-$Year_\tau$ , with $Year_\tau$ being sampled over 1996-1999. PLHIV diagnosed through a confirmed reactive self-test initiate ART at a rate $\rho_{ST}$, informed by ATLAS survey data (see **Figure S1d**).

$U_{i,u,s}^{r,a}(t) = 0$ if u ≤ 2

$U_{i,u=3,s}^{r,a}(t) = -UStart_i\ RRUStart_r(t)\ X_{i,u=3,s}^{r,a}\ (t)$

$U_{i,u=4,s}^{r,a}(t) = -\rho_{ST}\ X_{i,u=4,s}^{r,a}\ (t)$

$$U_{i,u=5,s=0}^{r,a}(t) = UStart_i\ RRUStart_r(t)\ X_{i,u=3,s=0}^{r,a}\ (t) + \Big(\big(UStart_i\ RRUStart_r(t)\big) - \big(UStop\ RRUStop(t)\big)\Big)\ X_{i,u=6,s=0}^{r,a}\ (t)$$

$$U_{i,u=5,s=1}^{r,a}(t) = UStart_i\ RRUStart_r(t)\ X_{i,u=3,s=1}^{r,a}\ (t) + \rho_{ST}\ X_{i,u=4,s}^{r,a}\ (t) + \Big(\big(UStart_i\ RRUStart_r(t)\big) - \big(UStop\ RRUStop(t)\big)\Big)\ X_{i,u=6,s=1}^{r,a}\ (t)$$

$U_{i,u=6,s}^{r,a}(t) = UStop\ RRUStop(t)\ X_{i,u=6,s}^{r,a}\ (t)$

The parameter $UStart_i$ is the "base" rate of ART initiation and depend on PLHIV HIV stage of infection, whereas $UStop$ is the "base" rate of ceasing ART. As in [1,3], we assumed that PLHIV with a diagnosed infection and a CD4 count <200 cells/µL were more likely to show clinical symptoms and initiate ART. A linear relation was assumed between CD4 count stage and initiation rate using parameter $\varpi$. This parameter was given a uniform prior over [0-1] so that PLHIV with lower CD4 cell counts always had higher initiation rates. ART initiation rates were first sampled among PLHIV with a CD4 count <200 cells/µL ($UStart_4 = \rho_4$), and the rates among the other infection stages were calculated using the formula described in [1], where $m = ((0 - \rho_4)/9.25)\ \varpi$ , $UStart_3 = \rho_3 = 3m - \rho_4$, $UStart_2 = \rho_2 = 6m - \rho_4$, and $UStart_1 = \rho_1 = 9m - \rho_4$.

The parameter $RRUStart_r(t)$ is a product of cofactors for initiating ART (see **Table S1c**): the parameter $RR_{\rho 2000}$ is the relative risk of ART initiation among PLHIV with a diagnosed infection at the time $Year_\tau$ compared to 2020, and we assumed that ART initiation rates would linearly increase between $Year_\tau$ and 2020, then stay constant at the 2020 levels after this period. Our model allowed the ART initiation rates to differ between KP and non-KP, using a wide prior range for a parameter $RR_{\rho KP}$.

Similarly, the parameter $RRUStop_r(t)$ is a product of cofactors for ceasing ART: our model assumed that ART drop-out rates could slightly decrease over time and be different between FSW and MSM compared to non-KP and FSW clients.

Our model used a specific ART initiation rate among people that had confirmed a reactive HIV reactive self-test ($\rho_{ST}$), which could be directly informed by ATLAS data (see specific section).

A fraction of PLHIV on ART have a suppressed viral load and are assumed not to be able to transmit HIV. This fraction increases over time and vary by sex ($VLS_g(t)$), using estimates from UNAIDS[67]. The scaling factor $VLSPtl$ sampled between 0 and 1 was used to reflect uncertainties in estimates as well as overall time trends.

#### HIV self-testing $S_{i,u,s}^{r,a}$

We calculated the number of HIV self-test kits used by each modelled population at each timestep ($\upsilon_{i,u}^{r,a}$). For the period 2019-2021, this number was based on the ATLAS programmatic data (available for each quarter over 2019-2021) and survey data described in the "HIVST scenarios" of this supplement. From January 2022, the number of kits used was calculated by assuming that 95% of KP HIV-uninfected or living with HIV but not on ART should receive two HIV tests each year (see specific section on HIV self-testing). The parameters $STSe_i$ and $STSp$ represent the sensitivity and specificity of the self-tests, whereas $STLnk$ is the fraction of reactive self-tests which are followed by conventional tests (informed by ATLAS survey data). False-reactive self-tests among people not living with HIV were contradicted by subsequent non-reactive tests or negative conventional tests (see Figure S1d), whereas false non-reactive self-tests among PLHIV would prevent them to undertake confirmatory tests in facilities.

Transitions between model states $S_{i,u,s}^{r,a}$ are shown in **Figure S1d** and only apply to:

1) people who have never tested for HIV (*u*=0), or ever had a conventional test but never a self-test (*u*=1, *s*=0):

$$S_{i,u=0-1,s=0}^{r,a} = -\upsilon_{i,u=0-1,s=0}^{r,a}$$

2) people not living with HIV (*i*=0) who have ever had a conventional HIV test but never had a self-test (*u*=1, *s*=0), and that can have a first self-test that is a false reactive:

$$S_{i=0,u=1,s=0}^{r,a} = -\upsilon_{i=0,u=1,s=0}^{r,a}(1 - STSp)$$

3) people not living with HIV (*i*=0) who have ever had a conventional HIV test, ever had a self-test, and their last self-test was not reactive (*u*=1, *s*=1):

$$S^{r,a}_{i=0,u=1,s=1} = \left[\left(v^{r,a}_{i=0,u=0,s=0} + v^{r,a}_{i=0,u=1,s=0} + v^{r,a}_{i=0,u=2,s=1}\right)STSp\right] - v^{r,a}_{i=0,u=1,s=1}(1 - STSp)$$

4) people not living with HIV (*i*=0) who have ever had a conventional HIV test and a self-test, but their last self-test was reactive and they haven't had a conventional test since (*u*=2, *s*=1):

$$S^{r,a}_{i=0,u=2,s=1} = \left[\left(v^{r,a}_{i=0,u=0,s=0} + v^{r,a}_{i=0,u=1,s=0} + v^{r,a}_{i=0,u=1,s=1}\right)(1 - STSp)\right] - v^{r,a}_{i=0,u=2,s=1}STSp$$

5) PLHIV (*i* >0) who are currently undiagnosed (*u*=1) and have never tested for HIV (*s*=0):

$$S^{r,a}_{i>0,u=1,s=0} = -v^{r,a}_{i>0,u=1,s=0}$$

6) PLHIV (*i* >0) who are currently undiagnosed (*u*=1) and have ever tested for HIV (*s*=1), some of which may have had a reactive self-test, but not followed by a confirmation test (fraction $(1 - STLnk)$).

$$S^{r,a}_{i>0,u=1,s=1} = \sum_{u=0-1} v^{r,a}_{i>0,u,s=0}\left((1 - STSe_i) + (STSe_i \times (1 - STLnk))\right) - v^{r,a}_{i>0,u=1,s=1}\, STSe_i$$

7) PLHIV who have had a reactive self-test, are undiagnosed but can do a confirmation test (*u*=2).

$$S^{r,a}_{i>0,u=2,s=1} = \left[v^{r,a}_{i>0,u=0,s=0} + v^{r,a}_{i>0,u=1,s=0} + v^{r,a}_{i>0,u=1,s=1}\right] STSe_i$$

8) PLHIV who have never used a self-test (*s*=0) but that have a diagnosed infection (*u*=3,5,6). Our base case scenario assumes that PLHIV in ART (*u*=5) do not receive self-tests.

$$S^{r,a}_{i>0,u>2,s=0} = -v^{r,a}_{i>0,u>2,s=0}$$

7) PLHIV who have ever used a self-test (*s*=1) but that have a diagnosed infection (*u*=3,5,6).

$$S^{r,a}_{i>0,u>2,s=1} = v^{r,a}_{i>0,u>2,s=0}$$

#### Model fitting overview

Each model was fitted under a Bayesian framework in three steps. In the first step, a Latin hypercube of model parameters was used to simulate 50M simulations from prior distributions of the parameters describe above. In the second step, we only retained the simulations which agreed with all the widen confidence intervals of the fitting outcomes described in **Tables S2a-c** (between 579 and 1550 simulations were retained across models). In the third step, the 100 fitted simulations with the highest overall likelihood (calculated on all outcomes except on HIV incidence rate, number of conventional tests and fraction of positive tests, for which there were no sample size) were identified for each country. The resulting posterior parameter sets were used to simulate all our model scenarios.

#### Model fitting data

##### Fitting data (Côte d'Ivoire)

**Table S2a:** List of demographic, epidemiological, and intervention outcomes used for model fitting in Côte d'Ivoire

| Population or age group | Year | Point estimate | Original 95%CI | Prior constraint | Reference |
|---|---|---|---|---|---|
| **Population size** | | | | | |
| Total number of 15-59 years-old | 1970<br>1980<br>1990<br>2000<br>2010<br>2020 | 2.58 million<br>4.05 million<br>6.02 million<br>8.51 million<br>10.64 million<br>14.19 million | N.A. | Initial value for 1970 and direct calibration using growth rate between 1970 and 2020 estimates | From [4] |
| **Age distribution among 15–59-year-old females** | | | | | |
| | 1970 | 15-24 years: 33.5%<br>25-49 years: 55.5%<br>50-59 years: 10.9% | N.A. | Used for comparison | From [4] |
| | 1980 | 15-24 years: 37.1%<br>25-49 years: 52.7%<br>50-59 years: 10.4% | N.A. | Used for comparison | As above |
| | 1990 | 15-24 years: 38.1%<br>25-49 years: 51.7%<br>50-59 years: 10.2% | N.A. | Used for comparison | As above |
| | 2000 | 15-24 years: 40.0%<br>25-49 years: 51.1%<br>50-59 years: 9.0% | N.A. | Used for comparison | As above |

| | | | | | |
|---|---|---|---|---|---|
| | 2010 | 15-24 years: 39.6%<br>25-49 years: 51.0%<br>50-59 years: 9.3% | N.A. | Used for comparison | As above |
| | 2020 | 15-24 years: 38.7%<br>25-49 years: 52.3%<br>50-59 years: 9.0% | N.A. | 35.2-42.6%<br>47.5-57.5%<br>6.9-11.7% | Fitted from [4] |
| **Age distribution among 15–59-year-old males** | | | | | |
| | 1970 | 15-24 years: 32,2%<br>25-49 years: 57.5%<br>50-59 years: 10.3% | N.A. | Used for comparison | From [4] |
| | 1980 | 15-24 years: 33.4%<br>25-49 years: 56.1%<br>50-59 years: 10.5% | N.A. | Used for comparison | As above |
| | 1990 | 15-24 years: 33.9%<br>25-49 years: 54.9%<br>50-59 years: 11.3% | N.A. | Used for comparison | As above |
| | 2000 | 15-24 years: 36.6%<br>25-49 years: 52.7%<br>50-59 years: 10.7% | N.A. | Used for comparison | As above |
| | 2010 | 15-24 years: 37.6%<br>25-49 years: 51.7%<br>50-59 years: 10.7% | N.A. | Used for comparison | As above |
| | 2020 | 15-24 years: 38.0%<br>25-49 years: 52.1%<br>50-59 years: 9.9% | N.A. | 34.5-41.8%<br>47.4-57.3%<br>7.6-12.9% | Fitted from [4] |
| **HIV prevalence among all adult females (except female sex workers)** | | | | | |
| 15-24 | 1989 | 2.5% | (0.9-4.4%) | 0.1-18.0% | [111] |
| 15-24 | 2005 | 2.4% | (1.6-3.0%) | 1.0-10.0% | [11] |
| 15-24 | 2012 | 2.2% | (1.5-3.0%) | 1.0-10.0% | [10] |
| 15-24 | 2017 | 0.9% | (0.5-1.4%) | 0.2-6.0% | [31] |
| 15-24 | 2018 | 0.4% | (0.0-0.8%) | Only used for comparison | [112] |
| 25-49 | 1989 | 2.5% | (1.0-4.3%) | 0.8-20.0% | [111] |
| 25-49 | 2005 | 9.9% | (8.4-12.0%) | 4.0-18.0% | [11] |
| 25-49 | 2012 | 6.3% | (5.2-8.0%) | 3.0-15.0% | [10] |
| 25-49 | 2017 | 5.5% | (4.5-6.8%) | 3.0-8.0% | [31] |
| 50-59 | 1989 | 1.9% | (0.4-4.4%) | 0.1-10.0% | [111] |
| 50-59 | 2005 | 10.2% | (6.2-16.0%) | 4.0-20.0% | [11] |
| 50-59 | 2012 | 9.5% | (5.7-15.0%) | 4.0-20.0% | [10] |
| 50-59 | 2017 | 8.2% | (5.0-13.1%) | 4.0-20.0% | [31] |
| **HIV prevalence among all adult males** | | | | | |
| 15-24 | 1989 | 2.4% | (0.8-4.3%) | 0.5-15.0% | [111] |
| 15-24 | 2005 | 0.3% | (0.1-1.0%) | 0.1-4.0% | [11] |
| 15-24 | 2012 | 0.3% | (0.1-1.0%) | 0.1-4.0% | [10] |
| 15-24 | 2017 | 0.3% | (0.1-0.9%) | 0.1-4.0% | [31] |
| 15-24 | 2018 | 0.3% | (0.0-0.6%) | Only used for comparison | [112] |
| 25-49 | 1989 | 7.7% | (5.5-10.0%) | 2.0-40.0% | [111] |
| 25-49 | 2005 | 4.8% | (3.6-6.0%) | 2.0-15.0% | [11] |
| 25-49 | 2012 | 4.3% | (3.3-5.0%) | 2.0-15.0% | [10] |
| 25-49 | 2017 | 2.1% | (1.4-2.9%) | 1.0-4.0% | [31] |
| 50-59 | 1989 | 1.7% | (0.4-3.9%) | 1.0-25.0% | [111] |
| 50-59 | 2005 | 4.9% | (2.4-10.0%) | 2.0-20.0% | [11] |
| 50-59 | 2012 | 8.7% | (5.4-14.0%) | 4.0-30.0% | [10] |
| 50-59 | 2017 | 3.6% | (1.7-7.4%) | 2.0-15.0% | [31] |
| **HIV prevalence among all female sex workers** | | | | | |
| 15-59 | 1986 | 36.9% | (27.6-46.7%) | 10.0-85.0% | [113] |

| | | | | | |
|---|---|---|---|---|---|
| 15-59 | 1987 | 36.9% | (30.1-48.2%) | 10.0-85.0% | [113] |
| 15-59 | 1989 | 47.6% | (38.4-56.7%) | 10.0-85.0% | [113] |
| 15-59 | 1990 | 68.4% | (57.0-78.6%) | 10.0-85.0% | [113] |
| 15-59 | 1994 | 67.0% | (63.2-70.8%) | 10.0-85.0% | [68] |
| 15-59 | 1995 | 54.0% | (50.5-57.4%) | 10.0-85.0% | [68] |
| 15-59 | 1996 | 52.0% | (48.7-55.2%) | 10.0-85.0% | [68] |
| 15-59 | 1997 | 52.0% | (48.7-55.4%) | 10.0-85.0% | [68] |
| 15-59 | 1998 | 32.0% | (28.9-35.3%) | 10.0-65.0% | [114] |
| 15-59 | 1999 | 32.0% | (28.7-35.5%) | 10.0-65.0% | [114] |
| 15-59 | 2000 | 28.0% | (24.8-31.4%) | 10.0-55.0% | [114] |
| 15-59 | 2001 | 31.0% | (27.7-34.4%) | 10.0-55.0% | [114] |
| 15-59 | 2002 | 27.0% | (23.8-30.3%) | 10.0-55.0% | [114] |
| 15-59 | 2003 | 33.0% | (28.0-38.0%) | 10.0-55.0% | [115] |
| 15-59 | 2004 | 27.0% | (22.4-31.9%) | 10.0-55.0% | [115] |
| 15-59 | 2005 | 18.0% | (14.1-22.4%) | 8.0-40.0% | [115] |
| 15-59 | 2006 | 19.0% | (15.1-23.6%) | 8.0-40.0% | [115] |
| 15-59 | 2007 | 21.0% | (16.9-25.7%) | 8.0-40.0% | [115] |
| 15-59 | 2007 | 22.9% | (13.7-35.6%) | 8.0-40.0% | [116] |
| 15-59 | 2008 | 19.0% | (15.1-23.6%) | 8.0-40.0% | [115] |
| 15-59 | 2009 | 20.0% | (15.9-22.5%) | 8.0-40.0% | [115] |
| 15-59 | 2009 | 11.2% | (6.4-18.9%) | 8.0-40.0% | [116] |
| 15-59 | 2010 | 21.0% | (10.0-35.0%) | 8.0-40.0% | [115] |
| 15-59 | 2014 | 11.0% | (8.3-14.3%) | 5.0-25.0% | [95] |
| 15-59 | 2016 | 11.4% | (8.8-14.8%) | 5.0-25.0% | [117] |
| 15-59 | 2020 | 4.9% | (3.8-6.3%) | 2.0-15.0% | [118] |
| **HIV prevalence among clients of sex workers** | | | | | |
| 15-59 | 1999 | 13.4% | (10.5-17.0%) | 5.0-30.0% | [77] |
| **HIV Prevalence among MSM** | | | | | |
| All 15-59 | 2015 | 11.2% | (9.6-13.1%) | 5.0-25.0% | [42] |
| All 15-59 | 2016 | 19.6% | N.A. | 10.0-40.0% | [119] |
| All 15-59 | 2017 | 12.3% | (9.3-16.1%) | 5.0-25.0% | [117] |
| All 15-59 | 2020 | 6.4% | (5.2-7.5%) | 4.0-20.0% | [46] |
| All 15-24 | 2012 | 12.5% | (6.8-18.2%) | 3.0-35.0% | [40] |
| All 15-24 | 2015 | 11.4% | (6.6-19.0%) | 3.0-25.0% | [42] |
| All 15-24 | 2020 | 5.1% | (3.4-6.8%) | 2.0-15.0% | [46] |
| All 25-49 | 2012 | 24.8% | (16.7-34.9%) | 10.0-60.0% | [40] |
| All 25-49 | 2015 | 16.3% | (7.5-32.0%) | 5.0-40.0% | [42] |
| All 25-49 | 2020 | 11.2% | (7.3-14.9%) | 3.0-22.0% | [46] |
| All MSMW | 2012 | 12.3% | (9.2-16.3%) | 5.0-25.0% | [40] |
| All MSMW` | 2020 | 6.2% | (4.1-8.3%) | 3.0-15.0% | [46] |
| All MSME | 2012 | 25.7% | (20.8-31.3%) | 10.0-45.0% | [40] |
| All MSME | 2020 | 6.2% | (4.0-8.2%) | 3.0-15.0% | [46] |
| **HIV incidence rate (per 100 susceptible-year)** | | | | | |
| 15-59 | 2005 | 0.161 | (0.069-0.304) | 0.03-0.75 | [120] |
| 15-59 | 2010 | 0.152 | (0.065-0.286) | 0.03-0.75 | As above |
| 15-59 | 2017 | 0.129 | (0.055-0.243) | 0.05-0.24 | As above |
| **Number of new HIV infections** | | | | | |
| 15-59 | 2005 | 18100 | (7900-34000) | 3000-65000 | [120] |
| 15-59 | 2010 | 22000 | (9700-41000) | 4000-80000 | As above |
| 15-59 | 2017 | 26000 | (11400-49000) | 11400-49000 | As above |
| **Number of HIV-related deaths** | | | | | |
| 15-59 | 2005 | 48000 | (29000-72000) | 1000-100000 | [120] |
| 15-59 | 2010 | 29000 | (17500-45000) | 5000-70000 | As above |
| 15-59 | 2017 | 21700 | (12700-32000) | 12700-32000 | As above |
| **Fraction of all females ever tested for HIV** | | | | | |
| 15-49 | 2000 | 7.2% | (6.7-7.7%) | 1.0-25.0% | [121] |

| | | | | | |
|---|---|---|---|---|---|
| 15-49 | 2005 | 10.9% | (8.2-14.5%) | 5.0-35.0% | [11] |
| 15-49 | 2011 | 35.4% | (33.1-37.8%) | 20.0-50.0% | [10] |
| 15-49 | 2016 | 56.0% | (53.9-58.0%) | 35.0-75.0% | [122] |
| 15-24 | 2009 | 25.3% | (23.6-27.1) | 10.0-50.0% | [123] |
| **Fraction of all females not living with HIV ever tested for HIV** | | | | | |
| 15-49 | 2005 | 10.0% | (7.2-13.7%) | 4.0-25.0% | [11] |
| 15-49 | 2011 | 34.5% | (31.8-37.2%) | 15.0-55.0% | [10] |
| 15-49 | 2017 | 56.3% | (54.3-58.2%) | 35.0-75.0% | [31] |
| **Fraction of all females living with HIV ever tested for HIV** | | | | | |
| 15-49 | 2005 | 13.6% | (7.9-22.5%) | 5.0-40.0% | [11] |
| 15-49 | 2011 | 42.0% | (34.2-50.2%) | 20.0-70.0% | [10] |
| 15-49 | 2017 | 74.7% | (67.5-82.0%) | 50.0-95.0% | [31] |
| **Fraction of all males ever tested for HIV** | | | | | |
| 15-49 | 2005 | 7.9% | (6.2-9.9%) | 3.0-25.0% | [11] |
| 15-49 | 2011 | 23.1% | (20.7-25.7%) | 10.0-40.0% | [10] |
| 15-49 | 2016 | 34.6% | (32.1-37.1%) | 20.0-55.0% | [122] |
| 15-24 males | 2009 | 18.1% | (16.7-19.6%) | 5.0-40.0% | [123] |
| **Fraction of all males not living with HIV ever tested for HIV** | | | | | |
| 15-49 | 2005 | 7.4% | (6.2-9.9%) | 3.0-20.0% | [11] |
| 15-49 | 2011 | 23.0% | (20.7-25.7%) | 10.0-40.0% | [10] |
| 15-49 | 2017 | 32.2% | (30.1-34.4%) | 20.0-45.0% | [31] |
| **Fraction of all males living with HIV ever tested for HIV** | | | | | |
| 15-49 | 2005 | 23.7% | (10.9-44.1%) | 8.0-50.0% | [11] |
| 15-49 | 2011 | 39.0% | (28.4-50.7%) | 25.0-65.0% | [10] |
| 15-49 | 2017 | 53.4% | (39.1-67.6%) | 30.0-75.0% | [31] |
| **Fraction of FSW ever tested for HIV** | | | | | |
| 15-59 | 2007 | 54.0% | (52.0-56.0%) | 30.0-85.0% | [124] |
| 15-59 | 2020 | 82.0% | (79.0-83.0%) | 65.0-92.0% | [118] |
| 15-24 | 2014 | 75.3% | (68.5-81.0%) | 50.0-95.0% | [95] |
| 25-49 | 2014 | 85.7% | (81.1-89.3%) | 65.0-99.0% | [95] |
| **Fraction of MSM ever tested for HIV** | | | | | |
| 15-59 | 2011 | 62.6% | (56.5-68.2%) | 40.0-95.0% | [40] |
| 15-59 | 2015 | 92.4% | (85.7-96.1%) | 70.0-99.0% | [125] |
| 15-59 | 2020 | 70.0% | (67.0-72.0%) | 60.0-90.0% | [46] |
| **Fraction of all females living with HIV which are diagnosed** | | | | | |
| 15-59 | 2015 | 68.0% | (61.0-77.0%) | 40.0-85.0% | [126] |
| 15-59 | 2016 | 71.0% | (64.0-81.0%) | 40.0-90.0% | [126] |
| 15-59 | 2017 | 75.0% | (67.0-85.0%) | 45.0-95.0% | [126] |
| 15-59 | 2018 | 78.0% | (70.0-89.0%) | 50.0-99.0% | [126] |
| 15-59 | 2019 | 81.0% | (72.0-92.0%) | 55.0-99.0% | [126] |
| 15-59 | 2020 | 84.0% | (75.0-95.0%) | 70.0-99.0% | [126] |
| **Fraction of all males living with HIV which are diagnosed** | | | | | |
| 15-59 | 2015 | 49.0% | (43.0-58.0%) | 30.0-75.0% | [126] |
| 15-59 | 2016 | 52.0% | (46.0-62.0%) | 30.0-75.0% | [126] |
| 15-59 | 2017 | 56.0% | (50.0-66.0%) | 30.0-80.0% | [126] |
| 15-59 | 2018 | 60.0% | (54.0-71.0%) | 40.0-80.0% | [126] |
| 15-59 | 2019 | 64.0% | (58.0-75.0%) | 45.0-85.0% | [126] |
| 15-59 | 2020 | 68.0% | (61.0-79.0%) | 54.0-85.0% | [126] |
| **Fraction of all FSW living with HIV with a diagnosed infection** | | | | | |
| 15-59 | 2014 | 26.7% | (15.9-41.0%) | 5.0-70.0% | [95] |
| 15-59 | 2020 | 81.0% | (69.0-89.0%) | 50.0-95.0% | [118] |
| **Fraction of all MSM living with HIV with a diagnosed infection** | | | | | |
| 15-59 | 2011 | 15.9% | (10.3-23.8%) | 30.0-50.0% | [40] |
| 15-59 | 2015 | 37.0% | (29.6-45.1%) | 15.0-65.0% | [36] |
| 15-59 | 2017 | 26.7% | (16.0-41.0%) | 10.0-60.0% | [44] |
| 15-59 | 2020 | 32.7% | (34.1-42.4%) | 15.0-60.0% | [46] |
| **Fraction of all females living with HIV with a treated infection** | | | | | |
| 15-59 | 2015 | 44.0% | (40.0-50.0%) | 25.0-60.0% | [67] |
| 15-59 | 2020 | 83.0% | (74.0-94.0%) | 69.0-99.0% | [67] |

| | | | | | |
|---|---|---|---|---|---|
| **Fraction of all males living with HIV with a treated infection** | | | | | |
| 15-59 | 2015 | 29.0% | (25.0-34.0%) | 15.0-50.0% | [67] |
| 15-59 | 2020 | 61.0% | (55.0-71.0%) | 49.0-78.0% | [67] |
| **Fraction of all FSW living with HIV with a treated infection** | | | | | |
| 15-59 | 2012 | 45.6% | (42.2-49.1%) | 20.0-70.0% | [115] |
| 15-59 | 2020 | 65.0% | (52.0-76.0%) | 52.0-76.0% | [118] |
| **Fraction of PLHIV with a suppressed viral load** | | | | | |
| 15-49 females | 2017 | 38.4% | (29.2-47.7%) | 20.0-55.0% | [31] |
| 15-49 males | 2017 | 20.1% | (11.9-28.3%) | 5.0-40.0% | [31] |
| 15-49 MSM | 2017 | 19.4% | (9.8-35.2%) | 10.0-45.0% | [44] |
| **Number of conventional HIV tests done by females each year** | | | | | |
| 15-59 | 2015 | 1,601,691 | N.A. | (800,846-3,203,382) | Programmatic data reported by countries to UNAIDS's Shyny90[126] |
| 15-59 | 2016 | 1,826,826 | N.A. | (913,413-3,653,652) | As above |
| 15-59 | 2017 | 1,631,236 | N.A. | (815,718-3,262,672) | As above |
| 15-59 | 2018 | 1,809,731 | N.A. | (904,866-3,619,462) | As above |
| **Number of conventional HIV tests done by males each year** | | | | | |
| 15-59 | 2015 | 492,691 | N.A. | (246,346-985,382) | [126] |
| 15-59 | 2016 | 553,680 | N.A. | (276,840-1,107,360) | As above |
| 15-59 | 2017 | 437,692 | N.A. | (218,846-975,384) | As above |
| 15-59 | 2018 | 902,838 | N.A. | (451,419-1,805,676) | As above |
| **Fraction of conventional HIV tests done by females which are positive** | | | | | |
| 15-59 | 2015 | 3.2% | N.A. | (1.6-6.5%) | Programmatic data reported by countries to UNAIDS's Shyny90[126] |
| 15-59 | 2016 | 2.6% | N.A. | (1.3-5.2%) | As above |
| 15-59 | 2017 | 2.2% | N.A. | (1.1-4.4%) | As above |
| 15-59 | 2018 | 1.8% | N.A. | (0.9-3.5%) | As above |
| **Fraction of conventional HIV tests done by males which are positive** | | | | | |
| 15-59 | 2015 | 3.6% | N.A. | (1.8-7.1%) | Programmatic data reported by countries to UNAIDS's Shyny90[126] |
| 15-59 | 2016 | 3.0% | N.A. | (1.5-6.0%) | As above |
| 15-59 | 2017 | 3.0% | N.A. | (1.5-6.0%) | As above |
| 15-59 | 2018 | 3.5% | N.A. | (1.8-7.0%) | As above |

MSMW: men who have sex with men as well as female partners; MSME: men who have sex with men exclusively.
N.A.: Not available

#### Fitting data (Mali)

| **Table S2b:** List of demographic, epidemiological, and intervention outcomes used for model fitting in Mali | | | | | |
|---|---|---|---|---|---|
| **Population or age group** | **Year** | **Point estimate** | **Original 95%CI** | **Prior constraint** | **Reference** |
| **Population size** | | | | | |
| Total number of 15-59 years-old | 1970<br>1980<br>1990<br>2000<br>2010<br>2020 | 3.15 million<br>3.57 million<br>4.00 million<br>5.30 million<br>7.27 million<br>9.95 million | N.A. | Initial value for 1970 and direct calibration using growth rate between 1970 and 2020 estimates | From [4] |
| **Age distribution among 15–59-year-old females** | | | | | |
| | 1970 | 15-24 years: 35.0%<br>25-49 years: 51.6%<br>50-59 years: 13.4% | N.A. | Used for comparison | From [4] |
| | 1980 | 15-24 years: 36.3%<br>25-49 years: 51.5%<br>50-59 years: 12.2% | N.A. | Used for comparison | As above |
| | 1990 | 15-24 years: 39.0%<br>25-49 years: 49.8%<br>50-59 years: 11.2% | N.A. | Used for comparison | As above |
| | 2000 | 15-24 years: 40.5%<br>25-49 years: 49.3%<br>50-59 years: 10.2% | N.A. | Used for comparison | As above |
| | 2010 | 15-24 years: 39.0%<br>25-49 years: 51.7%<br>50-59 years: 9.3% | N.A. | Used for comparison | As above |
| | 2020 | 15-24 years: 39.9%<br>25-49 years: 51.1%<br>50-59 years: 9.0% | N.A. | 30.7-51.9%<br>39.3-66.4%<br>6.0-13.5% | Fitted from [4] |
| **Age distribution among 15–59-year-old males** | | | | | |
| | 1970 | 15-24 years: 35.8%<br>25-49 years: 51.2%<br>50-59 years: 13.0% | N.A. | Initial value from data | From [4] |
| | 1980 | 15-24 years: 38.0%<br>25-49 years: 50.2%<br>50-59 years: 11.8% | N.A. | Used for comparison | As above |
| | 1990 | 15-24 years: 42.0%<br>25-49 years: 47.9%<br>50-59 years: 10.1% | N.A. | Used for comparison | As above |
| | 2000 | 15-24 years: 43.3%<br>25-49 years: 48.1%<br>50-59 years: 8.6% | N.A. | Used for comparison | As above |
| | 2010 | 15-24 years: 40.5%<br>25-49 years: 51.5%<br>50-59 years: 8.0% | N.A. | Used for comparison | As above |
| | 2020 | 15-24 years: 41.1%<br>25-49 years: 50.9%<br>50-59 years: 8.0% | N.A. | 31.6-53.4%<br>39.2-66.2%<br>5.3-12.0% | Fitted from [4] |
| **HIV prevalence among all adult females (except female sex workers)** | | | | | |
| 15-24 | 2001 | 1.3% | (0.7-2.0%) | 0.1-5.0% | [17] |
| 15-24 | 2006 | 0.9% | (0.4-1.5%) | 0.1-4.0% | [16] |

| | | | | | |
|---|---|---|---|---|---|
| 15-24 | 2013 | 1.1% | (0.6-1.7%) | 0.3-2.3% | [15] |
| 25-49 | 2001 | 2.6% | (1.0-4.3%) | 0.5-10.0% | [17] |
| 25-49 | 2006 | 1.7% | (1.0-2.4%) | 0.2-7.0% | [16] |
| 25-49 | 2013 | 1.5% | (1.0-1.9%) | 0.4-2.9% | [15] |
| **HIV prevalence among all adult males** | | | | | |
| 15-24 | 2001 | 0.3% | (0.0-0.7%) | 0.0-3.0% | [17] |
| 15-24 | 2006 | 0.5% | (0.0-0.9%) | 0.0-4.0% | [16] |
| 15-24 | 2013 | 0.3% | (0.0-0.7%) | 0.0-1.0% | [15] |
| 25-49 | 2001 | 2.0% | (1.0-4.3%) | 0.5-8.0% | [17] |
| 25-49 | 2006 | 1.1% | (0.5-1.8%) | 0.2-5.0% | [16] |
| 25-49 | 2013 | 1.1% | (0.6-1.6%) | 0.2-2.5% | [15] |
| 50-59 | 2006 | 1.2% | (0.0-2.4%) | 0.0-7.0% | [16] |
| 50-59 | 2013 | 1.2% | (0.0-2.3%) | 0.0-3.5% | [15] |
| **HIV prevalence among all female sex workers** | | | | | |
| 15-59 | 1987 | 36.0% | N.A. | 5.0-85.0% | [96] |
| 15-59 | 1995 | 46.0% | (38.8-53.4%) | 20.0-85.0% | [127] |
| 15-59 | 1997 | 30.4% | (24.3-37.2%) | 15.0-80.0% | [97] |
| 15-59 | 2000 | 28.9% | N.A. | 13.0-70.0% | [72] |
| 15-59 | 2003 | 31.9% | N.A. | 15.0-60.0% | [72] |
| 15-59 | 2006 | 35.3% | N.A. | 15.0-70.0% | [72] |
| 15-59 | 2009 | 24.2% | (21.4-27.2%) | 12.0-60.0% | [72] |
| 15-59 | 2013 | 18.3% | (14.8-22.5%) | 10.0-50.0% | [72] |
| 15-59 | 2017 | 20.8% | (16.6-25.7%) | 10.0-40.0% | [128] |
| 15-59 | 2018 | 20.4% | (16.3-25.0%) | 10.0-40.0% | [101] |
| 15-59 | 2019 | 8.7% | (7.3-10.4%) | 4.0-30.0% | [14] |
| **HIV prevalence among clients of sex workers** | | | | | |
| 15-59 | 2009 | 2.7% | (1.8-4.2%) | N.A. | [72] Only used for comparison (truck drivers) |
| 15-59 | 2019 | 1.9% | (1.2-2.9%) | N.A. | [14]Only used for comparison (truck drivers) |
| **HIV Prevalence among MSM** | | | | | |
| All 15-59 | 2011 | 20.1% | N.A. | 5.0-70.0% | [6] (original source not found) |
| All 15-59 | 2015 | 18.1% | (15.1-21.5%) | 3.0-32.0% | [129] |
| All 15-59 | 2015 | 9.5% | (5.6-13.5%) | 3.0-32.0% | [47] |
| All 15-24 | 2020 | 8.5% | (6.5-11.0%) | 4.0-15.0% | [32] |
| All 25-49 | 2020 | 19.1% | (15.6-23.1%) | 10.0-28.0% | [32] |
| **HIV incidence rate (per 100 susceptible-year)** | | | | | |
| 15-59 | 1990 | 0.255 | (0.128-0.398) | 0.03-1.40 | [67] |
| 15-59 | 1995 | 0.232 | (0.179-0.310) | 0.06-0.36 | As above |
| 15-59 | 2000 | 0.133 | (0.105-0.167) | 0.05-0.30 | As above |
| 15-59 | 2005 | 0.092 | (0.071-0.115) | 0.03-0.21 | As above |
| 15-59 | 2010 | 0.066 | (0.049-0.086) | 0.03-0.19 | As above |
| 15-59 | 2015 | 0.048 | (0.032-0.067) | 0.01-0.17 | As above |
| 15-59 | 2020 | 0.030 | (0.018-0.052) | 0.01-0.06 | As above |
| **Number of new HIV infections** | | | | | |
| 15-59 | 1990 | 11000 | (5600-17000) | 2600-37000 | [67] |
| 15-59 | 1995 | 11000 | (8700-15000) | 2700-35000 | As above |
| 15-59 | 2000 | 7400 | (5800-9300) | 1800-25300 | As above |
| 15-59 | 2005 | 5900 | (4500-7400) | 1200-14000 | As above |
| 15-59 | 2010 | 4900 | (3600-6400) | 1000-9000 | As above |
| 15-59 | 2015 | 4100 | (2800-5800) | 800-8000 | As above |
| 15-59 | 2020 | 3100 | (1800-5200) | 1800-7000 | As above |
| **Number of HIV-related deaths** | | | | | |
| 15-59 | 1990 | 1100 | (0-3200) | 0-8200 | [67] |
| 15-59 | 1995 | 3400 | (1600-6700) | 500-12300 | As above |
| 15-59 | 2000 | 6000 | (4100-8500) | 2100-15500 | As above |
| 15-59 | 2005 | 6400 | (5100-8000) | 2100-16000 | As above |
| 15-59 | 2010 | 4200 | (3300-5400) | 1300-9400 | As above |
| 15-59 | 2015 | 4300 | (3400-5400) | 1400-9400 | As above |

| | | | | | |
|---|---|---|---|---|---|
| 15-59 | 2020 | 3100 | (2200-4300) | 1500-5500 | As above |
| **Fraction of all females ever tested for HIV** | | | | | |
| 15-49 | 2001 | 4.2% | (3.5-5.0%) | 1.0-20.0% | [17] |
| 15-49 | 2009 | 14.4% | (13.3-15.6%) | 5.0-50.0% | [130] |
| 15-49 | 2015 | 19.9% | (18.0-21.9%) | 8.0-50.0% | [131] |
| 15-49 | 2018 | 17.9% | (15.8-20.1%) | 10.0-50.0% | [58] |
| **Fraction of all females not living with HIV ever tested for HIV** | | | | | |
| 15-49 | 2006 | 6.2% | (5.2-7.5%) | 2.0-25.0% | [16] |
| 15-49 | 2013 | 12.4% | (10.6-14.3%) | 5.0-45.0% | [15] |
| **Fraction of all females living with HIV ever tested for HIV** | | | | | |
| 15-49 | 2006 | 16.9% | N.A. | 5.0-50.0% | [126] |
| 15-49 | 2013 | 36.5% | N.A. | 20.0-60.0% | As above |
| 15-49 | 2020 | 54.7% | N.A. | 40.0-70.0% | As above |
| **Fraction of all males ever tested for HIV** | | | | | |
| 15-49 | 2001 | 9.0% | (7.3-10.9%) | 1.0-40.0% | [17] |
| 15-49 | 2015 | 17.3% | (15.7-19.0%) | 5.0-45.0% | [131] |
| 15-49 | 2018 | 14.7% | (32.1-37.1%) | 8.0-30.0% | [58] |
| **Fraction of all males not living with HIV ever tested for HIV** | | | | | |
| 15-49 | 2006 | 6.4% | (5.3-7.8%) | 2.0-30.0% | [16] |
| 15-49 | 2013 | 10.7% | (9.1-12.6%) | 4.0-30.0% | [15] |
| **Fraction of all males living with HIV ever tested for HIV** | | | | | |
| 15-49 | 2006 | 16.4% | N.A. | 8.0-45.0% | [126] |
| 15-49 | 2013 | 33.7% | N.A. | 15.0-60.0% | As above |
| 15-49 | 2020 | 49.8% | N.A. | 35.0-65.0% | As above |
| **Fraction of FSW ever tested for HIV** | | | | | |
| 15-59 | 2000 | 36.0% | N.A. | 2.0-70.0% | [98] |
| 15-59 | 2003 | 31.0% | N.A. | 10.0-60.0% | As above |
| 15-59 | 2006 | 51.0% | N.A. | 25.0-80.0% | As above |
| 15-59 | 2009 | 67.0% | N.A. | 30.0-90.0% | As above |
| 15-59 | 2018 | 45.8% | (34.8-57.3%) | 20.0-85.0% | [101] (All are living with HIV) |
| **Fraction of MSM ever tested for HIV** | | | | | |
| 15-59 | 2015 | 72.0% | N.A. | 50.0-95.0% | [129] |
| 15-59 | 2019 | 83.0% | (80.8-85.4%) | 60.0-90.0% | [32] |
| **Fraction of all females living with HIV which are diagnosed** | | | | | |
| 15-59 | 2015 | 47.0% | (39.0-56.0%) | 20.0-80.0% | [126] |
| 15-59 | 2016 | 50.0% | (42.0-60.0%) | 30.0-85.0% | As above |
| 15-59 | 2017 | 53.0% | (44.0-64.0%) | 30.0-85.0% | As above |
| 15-59 | 2018 | 56.0% | (47.0-68.0%) | 35.0-90.0% | As above |
| 15-59 | 2019 | 60.0% | (49.0-73.0%) | 40.0-90.0% | As above |
| 15-59 | 2020 | 64.0% | (53.0-78.0%) | 53.0-78.0% | As above |
| **Fraction of all males living with HIV which are diagnosed** | | | | | |
| 15-59 | 2015 | 36.0% | (30.0-43.0%) | 15.0-55.0% | [126] |
| 15-59 | 2016 | 38.0% | (32.0-45.0%) | 15.0-60.0% | As above |
| 15-59 | 2017 | 40.0% | (33.0-48.0%) | 20.0-60.0% | As above |
| 15-59 | 2018 | 43.0% | (35.0-51.0%) | 20.0-60.0% | As above |
| 15-59 | 2019 | 45.0% | (37.0-55.0%) | 25.0-65.0% | As above |
| 15-59 | 2020 | 49.0% | (39.0-59.0%) | 30.0-59.0% | As above |
| **Fraction of all FSW living with HIV with a diagnosed infection** | | | | | |
| 15-59 | 2018 | 45.8% | (34.8-57.3%) | 28.0-70.0% | [101] |
| **Fraction of all MSM living with HIV with a diagnosed infection** | | | | | |
| 15-59 | 2014 | 34.1% | (15.6-52.5%) | 15.0-65.0% | [47] |
| **Fraction of all females living with HIV with a treated infection** | | | | | |
| 15-49 | 2015 | 39.0% | (33.0-47.0%) | 20.0-60.0% | [67] |
| 15-49 | 2020 | 62.0% | (51.0-76.0%) | 45.0-82.0% | [67] |
| **Fraction of all males living with HIV with a treated infection** | | | | | |
| 15-49 | 2015 | 25.0% | (21.0-30.0%) | 10.0-50.0% | [67] |
| 15-49 | 2020 | 44.0% | (35.0-53.0%) | 30.0-60.0% | [67] |
| **Fraction of all MSM living with HIV with a treated infection** | | | | | |
| 15-59 | 2014 | 30.0% | (18.3-45.0%) | 10.0-60.0% | [47] |

| | | | | | |
|---|---|---|---|---|---|
| **Fraction of FSW living with HIV with a suppressed viral load** | | | | | |
| 15-49 | 2019 | 51.6% | (41.3-61.7%) | 30.0-70.0% | [14] |
| **Fraction of MSM living with HIV with a suppressed viral load** | | | | | |
| 15-49 | 2014 | 30.0% | (18.3-45.0%) | 10.0-60.0% | [47] |
| 15-24 | 2020 | 40.0% | (27.5-44.8%) | 20.0-65.0% | [32] |
| 25-49 | 2020 | 48.1% | (37.0-59.2%) | 30.0-70.0% | [32] |
| **Number of conventional HIV tests done each year (females and males combined)** | | | | | |
| 15-59 | 2015 | 393,007 | N.A. | (196,504-786,014) | Programmatic data reported by countries to UNAIDS's Shyny90[126] |
| 15-59 | 2016 | 400,005 | N.A. | (200,003-800,010) | As above |
| 15-59 | 2017 | 476,098 | N.A. | (238,049-952,196) | As above |
| 15-59 | 2018 | 565,838 | N.A. | (282,919-1,131,676) | As above |
| 15-59 | 2019 | 504,414 | N.A. | (252,207-1,008,828) | As above |
| **Fraction of conventional HIV tests done which are positive (females and males combined)** | | | | | |
| 15-59 | 2015 | 2.3% | N.A. | (1.2-4.6%) | Programmatic data reported by countries to UNAIDS's Shyny90[126] |
| 15-59 | 2016 | 1.7% | N.A. | (0.9-3.5%) | As above |
| 15-59 | 2017 | 2.6% | N.A. | (1.3-5.1%) | As above |
| 15-59 | 2018 | 2.3% | N.A. | (1.2-4.6%) | As above |
| 15-59 | 2019 | 2.5% | N.A. | (1.2-5.0%) | As above |

MSMW: men who have sex with men as well as female partners; MSME: men who have sex with men exclusively.
N.A.: Not available

#### Fitting data (Senegal)

| **Table S2c:** List of demographic, epidemiological, and intervention outcomes used for model fitting in Senegal | | | | | |
|---|---|---|---|---|---|
| **Population or age group** | **Year** | **Point estimate** | **Original 95%CI** | **Prior constraint** | **Reference** |
| **Population size** | | | | | |
| Total number of 15-59years-old | 1970<br>1980<br>1990<br>2000<br>2010<br>2020 | 2.18 million<br>2.74 million<br>3.64 million<br>4.92 million<br>6.56 million<br>8.81 million | N.A. | Initial value for 1970 and direct calibration using growth rate between 1970 and 2020 estimates | From [4] |
| **Age distribution among 15–59-year-old females** | | | | | |
| | 1970 | 15-24 years: 35.7%<br>25-49 years: 53.0%<br>50-59 years: 11.3% | N.A. | Used for comparison | From [4] |
| | 1980 | 15-24 years: 37.2%<br>25-49 years: 51.4%<br>50-59 years: 11.4% | N.A. | Used for comparison | As above |
| | 1990 | 15-24 years: 39.3%<br>25-49 years: 49.8%<br>50-59 years: 10.9% | N.A. | Used for comparison | As above |
| | 2000 | 15-24 years: 40.4%<br>25-49 years: 50.1%<br>50-59 years: 9.5% | N.A. | Used for comparison | As above |
| | 2010 | 15-24 years: 37.8%<br>25-49 years: 52.5%<br>50-59 years: 9.7% | N.A. | Used for comparison | As above |
| | 2020 | 15-24 years: 35.6%<br>25-49 years: 54.2%<br>50-59 years: 10.2% | N.A. | 27.4-46.3%<br>41.6-70.5%<br>6.8-15.3% | Fitted from [4] |
| **Age distribution among 15–59-year-old males** | | | | | |
| | 1970 | 15-24 years: 35.7%<br>25-49 years: 53.8%<br>50-59 years: 10.5% | N.A. | Initial value from data | From [4] |
| | 1980 | 15-24 years: 37.3%<br>25-49 years: 51.6%<br>50-59 years: 11.1% | N.A. | Used for comparison | As above |
| | 1990 | 15-24 years: 40.4%<br>25-49 years: 48.8%<br>50-59 years: 10.8% | N.A. | Used for comparison | As above |
| | 2000 | 15-24 years: 42.8%<br>25-49 years: 48.2%<br>50-59 years: 9.0% | N.A. | Used for comparison | As above |
| | 2010 | 15-24 years: 41.4%<br>25-49 years: 50.0%<br>50-59 years: 8.6% | N.A. | Used for comparison | As above |
| | 2020 | 15-24 years: 39.1%<br>25-49 years: 52.3%<br>50-59 years: 8.6% | N.A. | 30.1-50.8%<br>40.2-68.0%<br>5.7-12.9% | Fitted from [4] |
| **HIV prevalence among all adult females (except female sex workers)** | | | | | |
| 15-24 | 2005 | 0.5% | (0.1-0.8%) | 0.1-3.0% | [21] |
| 15-24 | 2011 | 0.3% | (0.1-0.5%) | 0.1-3.0% | [22] |

| | | | | | |
|---|---|---|---|---|---|
| 15-24 | 2017 | 0.2% | (0.0-0.5%) | 0.0-1.0% | [24] |
| 25-49 | 2005 | 0.8% | (0.4-1.3%) | 0.4-6.0% | [21] |
| 25-49 | 2011 | 0.8% | (0.4-1.1%) | 0.4-5.0% | [22] |
| 25-49 | 2017 | 0.7% | (0.5-1.3%) | 0.5-1.5% | [24] |
| **HIV prevalence among all adult males** | | | | | |
| 15-24 | 2005 | 0.1% | (0.0-0.2%) | 0.0-1.5% | [21] |
| 15-24 | 2011 | 0.1% | (0.0-0.1%) | 0.0-1.5% | [22] |
| 15-24 | 2017 | 0.1% | (0.0-0.2%) | 0.0-1.0% | [24] |
| 25-49 | 2005 | 0.7% | (0.1-1.2%) | 0.1-5.0% | [21] |
| 25-49 | 2011 | 0.7% | (0.3-1.2%) | 0.1-5.0% | [22] |
| 25-49 | 2017 | 0.6% | (0.3-1.0%) | 0.1-2.0% | [24] |
| 50-59 | 2011 | 0.8% | (0.1-1.4%) | 0.0-5.0% | [22] |
| 50-59 | 2017 | 1.2% | (0.3-2.1%) | 0.0-3.0% | [24] |
| **HIV prevalence among all female sex workers** | | | | | |
| 15-59 | 1988 | 16.1% | (14.5-18.0%) | 5.0-60.0% | [132] |
| 15-59 | 1989 | 3.1% | N.A. | 1.0-60.0% | [133] |
| 15-59 | 1994 | 10.1% | N.A. | 2.0-60.0% | [133] |
| 15-59 | 2000 | 20.1% | (18.0-22.4%) | 5.0-60.0% | [134] |
| 15-59 | 2006 | 19.8% | (16.8-23.1%) | 5.0-60.0% | [76] |
| 15-59 | 2010 | 18.5% | N.A. | 5.0-50.0% | [75] |
| 15-59 | 2015 | 6.6% | (5.4-8.7%) | 2.0-30.0% | [74] |
| 15-59 | 2015 | 3.3% | (1.5-5.2%) | 2.0-30.0% | [78] |
| 15-59 | 2017 | 8.1% | N.A. | 3.0-40.0% | [107] |
| 15-59 | 2019 | 5.8% | (4.0-7.0%) | 10.0-40.0% | [73] |
| **HIV prevalence among clients of sex workers** | | | | | |
| 15-59 | 1999 | 4.4% | (3.3-5.8%) | 1.0-30.0% | [135] |
| 15-59 | 2015 | 1.2% | (0.2-2.5%) | 0.5-10.0% | IBBS surveys [78] (personal communication of estimated from the unpublished client survey) |
| **HIV Prevalence among MSM** | | | | | |
| All 15-59 | 2012 | 38.6% | (30.2-47.8%) | 15.0-60.0% | [136] |
| All 15-59 | 2016 | 23.5% | (18.6-28.4%) | 10.0-40.0% | [78] (RDS-adjusted estimates) |
| All 15-24 | 2014 | 17.7% | (14.9-20.8%) | 5.0-30.0% | [54] |
| All 15-24 | 2017 | 19.4% | (16.6-22.5%) | 5.0-35.0% | [55] (data fitted among MSMW/MSME separately) |
| All 25-49 | 2014 | 18.1% | (14.5-22.4%) | 5.0-50.0% | [54] |
| All 25-49 | 2017 | 39.5% | (35.0-44.1%) | 16.0-60.0% | [55] |
| All MSMW | 2004 | 20.2% | (16.6-24.4%) | 5.0-60.0% | [50] |
| All MSMW | 2007 | 19.3% | (15.5-23.7%) | 5.0-50.0% | [79] |
| All MSMW | 2017 | 24.1% | (21.4-27.0%) | 15.0-35.0% | [55] |
| All MSME | 2004 | 34.1% | (21.5-49.4%) | 5.0-70.0% | [50] |
| All MSME | 2007 | 29.0% | (21.9-37.3%) | 13.0-70.0% | [79] |
| All MSME | 2017 | 37.7% | (32.0-43.7%) | 25.0-50.0% | [55] |
| **HIV incidence rate (per 100 susceptible-year)** | | | | | |
| 15-59 | 1990 | 0.036 | (0.028-0.046) | 0.010-0.12 | [67] |
| 15-59 | 1995 | 0.072 | (0.058-0.088) | 0.018-0.160 | As above |
| 15-59 | 2000 | 0.094 | (0.080-0.112) | 0.030-0.192 | As above |
| 15-59 | 2005 | 0.064 | (0.052-0.076) | 0.022-0.146 | As above |
| 15-59 | 2010 | 0.018 | (0.014-0.022) | 0.007-0.052 | As above |
| 15-59 | 2015 | 0.010 | (0.008-0.013) | 0.004-0.045 | As above |
| 15-59 | 2020 | 0.010 | (0.088-0.014) | 0.004-0.030 | As above |
| **Number of new HIV infections** | | | | | |
| 15-59 | 1990 | 1400 | (1100-1800) | 300-4500 | [67] |
| 15-59 | 1995 | 3300 | (2600-3900) | 1600-7000 | As above |
| 15-59 | 2000 | 4900 | (4100-5800) | 2100-9800 | As above |
| 15-59 | 2005 | 3800 | (3100-4500) | 2100-8500 | As above |
| 15-59 | 2010 | 1200 | (0-1500) | 0-3500 | As above |
| 15-59 | 2015 | 500 | (0-1000) | 0-3000 | As above |
| 15-59 | 2020 | 700 | (0-1300) | 0-2600 | As above |

| | | | | | |
|---|---|---|---|---|---|
| **Number of HIV-related deaths** | | | | | |
| 15-59 | 1990 | 100 | (0-500) | 0-2000 | [67] |
| 15-59 | 1995 | 500 | (0-1000) | 0-3000 | As above |
| 15-59 | 2000 | 1300 | (1100-1700) | 500-3700 | As above |
| 15-59 | 2005 | 2400 | (2000-2900) | 1000-3900 | As above |
| 15-59 | 2010 | 1200 | (0-1600) | 0-3000 | As above |
| 15-59 | 2015 | 1600 | (1200-2000) | 600-4000 | As above |
| 15-59 | 2020 | 800 | (0-1000) | 0-1500 | As above |
| **Fraction of all females ever tested for HIV** | | | | | |
| 15-49 | 2000 | 3.0% | (2.4-3.8%) | 0.0-20.0% | [137] |
| 15-49 | 2014 | 42.6% | (40.0-45.2%) | 25.0-80.0% | [27] |
| 15-49 | 2015 | 42.6% | (40.1-45.1%) | 25.0-80.0% | [26] |
| 15-49 | 2016 | 39.8% | (36.8-42.8%) | 10.0-50.0% | [25] |
| **Fraction of all females not living with HIV ever tested for HIV** | | | | | |
| 15-49 | 2005 | 2.8% | (2.2-3.6%) | 0.5-20.0% | [21] |
| 15-49 | 2010 | 27.3% | (25.3-29.4%) | 15.0-70.0% | [138] |
| 15-49 | 2017 | 46.1% | (44.1-48.1%) | 32.0-80.0% | [24] |
| **Fraction of all females living with HIV ever tested for HIV** | | | | | |
| 15-49 | 2005 | 6.7% | N.A. | 2.0-40.0% | [126] |
| 15-49 | 2010 | 50.0% | N.A. | 25.0-80.0% | As above |
| 15-49 | 2017 | 82.2% | N.A. | 70.0-99.0% | As above |
| **Fraction of all males ever tested for HIV** | | | | | |
| 15-49 | 2014 | 20.0% | (17.5-22.7%) | 12.0-70.0% | [27] |
| 15-49 | 2015 | 21.8% | (19.2-24.7%) | 12.0-70.0% | [26] |
| 15-49 | 2016 | 19.8% | (17.3-22.4%) | 10.0-70.0% | [25] |
| 15-49 | 2018 | 17.9% | (15.8-20.1%) | 10.0-60.0% | [23] |
| **Fraction of all males not living with HIV ever tested for HIV** | | | | | |
| 15-49 | 2005 | 3.7% | (2.8-4.9%) | 1.0-20.0% | [21] |
| 15-49 | 2010 | 16.7% | (15.1-18.6%) | 5.0-60.0% | [138] |
| 15-49 | 2017 | 19.0% | (17.3-20.8%) | 15.0-70.0% | [24] |
| **Fraction of all males living with HIV ever tested for HIV** | | | | | |
| 15-49 | 2005 | 4.9% | N.A. | 1.0-40.0% | [126] |
| 15-49 | 2010 | 30.0% | N.A. | 10.0-80.0% | As above |
| 15-49 | 2017 | 55.7% | N.A. | 35.0-90.0% | As above |
| **Fraction of FSW ever tested for HIV** | | | | | |
| 15-59 | 2006 | 63.2% | N.A. | 10.0-90.0% | [76] |
| 15-59 | 2010 | 73.6% | N.A. | 20.0-95.0% | [75] |
| 15-59 | 2013 | 58.0% | N.A. | 25.0-90.0% | [39] |
| 15-59 | 2015 | 72.4% | (68.5-76.0%) | 30.0-95.0% | [78] |
| 15-59 | 2018 | 79.4% | (72.3-85.0%) | 40.0-95.0% | [139] |
| **Fraction of MSM ever tested for HIV** | | | | | |
| 15-59 | 2003 | 13.3% | (8.9-19.5%) | 3.0-60.0% | [104] |
| 15-59 | 2004 | 10.8% | (8.1-14.0%) | 3.0-50.0% | [50] |
| 15-59 | 2007 | 34.1% | (30.1-38.4%) | 10.0-70.0% | [74,79] |
| 15-59 | 2012 | 86.6% | (79.3-91.6%) | 55.0-99.0% | [136] |
| 15-59 | 2014 | 72.6% | N.A. | 30.0-90.0% | [140] |
| 15-59 | 2014 | 69.1% | N.A. | 50.0-90.0% | [74] |
| 15-59 | 2015 | 70.2% | (66.7-74.4%) | 50.0-95.0% | [78] |
| 15-59 | 2017 | 82.6% | (80.2-84.7%) | 60.0-95.0% | [55] |
| 15-59 | 2018 | 54.0% | (46.6-61.3%) | 40.0-90.0% | [139] |
| **Fraction of all females living with HIV which are diagnosed** | | | | | |
| 15-59 | 2015 | 70.0% | (62.0-78.0%) | 45.0-90.0% | [126] |
| 15-59 | 2016 | 74.0% | (66.0-83.0%) | 50.0-90.0% | As above |
| 15-59 | 2017 | 79.0% | (71.0-89.0%) | 55.0-95.0% | As above |
| 15-59 | 2018 | 86.0% | (77.0-96.0%) | 65.0-99.0% | As above |
| 15-59 | 2019 | 92.0% | (83.0-99.0%) | 70.0-99.0% | As above |
| 15-59 | 2020 | 95.0% | (85.0-99.0%) | 80.0-99.0% | As above |
| **Fraction of all males living with HIV which are diagnosed** | | | | | |
| 15-59 | 2015 | 48.0% | (42.0-54.0%) | 25.0-75.0% | [126] |

| | | | | | |
|---|---|---|---|---|---|
| 15-59 | 2016 | 52.0% | (46.0-58.0%) | 30.0-75.0% | As above |
| 15-59 | 2017 | 56.0% | (50.0-63.0%) | 35.0-80.0% | As above |
| 15-59 | 2018 | 61.0% | (54.0-67.0%) | 40.0-85.0% | As above |
| 15-59 | 2019 | 65.0% | (58.0-72.0%) | 45.0-90.0% | As above |
| 15-59 | 2020 | 68.0% | (61.0-77.0%) | 56.0-85.0% | As above |
| **Fraction of all FSW living with HIV with a diagnosed infection** | | | | | |
| 15-59 | 2000 | 5.1% | (1.4-16.9%) | 0.0-40.0% | [141] |
| 15-59 | 2002 | 29.3% | (23.6-35.8%) | 5.0-70.0% | [142] |
| 15-59 | 2010 | 12.5% | (7.8-19.3%) | 5.0-80.0% | [75] |
| 15-59 | 2015 | 53.8% | (37.4-69.6%) | 20.0-90.0% | [139] |
| 15-59 | 2015 | 67.5% | (52.0-79.9%) | 20.0-90.0% | [143] |
| 15-59 | 2016 | 55.0% | (39.8-69.3%) | 30.0-80.0% | [78] |
| **Fraction of all MSM living with HIV with a diagnosed infection** | | | | | |
| 15-59 | 2014 | 48.8% | (34.2-63.5%) | 20.0-85.0% | [136] |
| 15-59 | 2016 | 13.2% | (9.4-18.4%) | 5.0-80.0% | [78] |
| 15-59 | 2018 | 63.4% | N.A. | 40.0-90.0% | [6] |
| **Fraction of all females living with HIV with a treated infection** | | | | | |
| 15-49 | 2015 | 56.0% | (50.0-63.0%) | 20.0-90.0% | [67] |
| 15-49 | 2020 | 95.0% | (85.0-99.0%) | 85.0-99.0% | [67] |
| **Fraction of all males living with HIV with a treated infection** | | | | | |
| 15-49 | 2015 | 35.0% | (31.0-39.0%) | 10.0-80.0% | [67] |
| 15-49 | 2020 | 61.0% | (54.0-69.0%) | 45.0-75.0% | [67] |
| **Fraction of all FSW living with HIV with a treated infection** | | | | | |
| 15-59 | 2016 | 37.5% | (24.2-53.0%) | 20.0-53.0% | [78] |
| **Fraction of all MSM living with HIV with a treated infection** | | | | | |
| 15-59 | 2016 | 10.0% | (24.2-53.0%) | 20.0-53.0% | [78] |
| 15-59 | 2019 | 38.0% | N.A. | 10.0-60.0% | [6] |
| **Fraction of FSW living with HIV with a suppressed viral load** | | | | | |
| 15-49 | 2019 | 48.0% | (38.5-57.7%) | 35.0-60.0% | [73] |
| **Number of conventional HIV tests done each year (females and males combined)** | | | | | |
| 15-59 | 2016 | 611,175 | N.A. | (305,588-1,222,350) | Programmatic data reported by countries to UNAIDS's Shyny90[126] |
| 15-59 | 2017 | 550,386 | N.A. | (275,193-1,100,772) | As above |
| 15-59 | 2018 | 669,438 | N.A. | (334,719-1,338,876) | As above |
| 15-59 | 2019 | 684,635 | N.A. | (342,318-1,369,270) | As above |
| **Fraction of conventional HIV tests done which are positive (females and males combined)** | | | | | |
| 15-59 | 2016 | 1.5% | N.A. | (0.7-2.9%) | Programmatic data reported by countries to UNAIDS's Shyny90[126] |
| 15-59 | 2017 | 1.6% | N.A. | (0.8-3.3%) | As above |
| 15-59 | 2018 | 1.1% | N.A. | (0.5-2.1%) | As above |
| 15-59 | 2019 | 1.2% | N.A. | (0.6-2.4%) | As above |

MSMW: men who have sex with men as well as female partners; MSME: men who have sex with men exclusively.
N.A.: Not available

### Methods: simulated HIVST scenarios

*Number of HIVST distributed by ATLAS over 2019-2021*

A total of 187,914 HIVST were distributed by ATLAS in Côte d'Ivoire over 2019-2021, including 105,904 tests (56%) through FSW channels, 53,864 (29%) through MSM channels, and 28,146 (15%) through other channels (48% of them within STI clinics, 48% to partners of PLHIV, and 3% to people who use drugs (PwUD)). Individuals reached by ATLAS received an average of two test kits, and it was expected that they would distribute some or sometimes all (e.g., PLHIV already aware of their status) their tests to their sexual partners or friends.

This analysis only used programmatic data on number of HIVST distributed through the FSW and MSM channels. We used the channel-specific age-distribution of individuals receiving the tests from the ATLAS programmatic data. The age distribution of HIVST distributed over 2019-2021 through the FSW and MSM channels of the three ATLAS countries, under our assumptions, is shown in **Table S3** and **Figure S2**.

**Table S3:** Number of HIV self-test (HIVST) kits distributed over 2019-2021 in each ATLAS country by distribution channel and age (% by group for each country)

| | Female sex workers (FSW) channel | | | Men who have sex with men (MSM) channel | | |
|---|---|---|---|---|---|---|
| Age (years) | 15-24 | 25-49 | 50+ | 15-24 | 25-49 | 50+ |
| Côte d'Ivoire | 46 300 (29%) | 58 900 (37%) | 800 (1%) | 26 200 (16%) | 27 400 (17%) | 200 (<1%) |
| Mali | 48 200 (37%) | 58 900 (45%) | 3100 (2%) | 8800 (7%) | 11 900 (9%) | 200 (<1%) |
| Senegal | 8100 (18%) | 20 500 (45%) | 1300 (3%) | 5900 (13%) | 9800 (21%) | 200 (<1%) |

*Primary and secondary distribution*

The first stage of an ATLAS phone survey, carried out between March and June 2021, administered sociodemographic questionnaires by telephone to individuals reporting having received ATLAS HIVST kits[144]. In the survey, the HIVST distribution channel corresponding to the test used by a participant was identified using a survey serial number printed on survey flyers distributed with HIVST kits. The proportion of tests being received by individuals from the different modelled risk groups was calculated using two scenarios that reflect uncertainties related to the characteristics of individuals receiving the tests. This uncertainty was partly due to the poor reporting of specific behaviours such as paying or receiving money for sex, or sex between men in the region.

Both scenarios assumed that the age group of the individual (directly or indirectly) receiving a test is the same age group as the individual that directly received the test in the programmatic data (groups are 15-24, 25-49, 50+ years old). A specific sensitivity analysis is looking at the predicted impact of HIVST scale-up when using the age distribution from the phone survey and not the programmatic data (see Figure S2). The model assumes that the fraction of HIVST used among those received is the same across risk groups (80% in most scenarios, see specific section), thus the age/risk distribution of people who received the tests is the same as the one who used the tests.

*HIVST distribution under the base-case scenarios*

All our base case scenarios used the sex distribution of the phone survey (1st stage) participants by HIVST distribution channel and reflected the distribution of HIVST within the different risk groups (through primary and secondary distribution) accounting for reporting biases described in qualitative studies. The **Table S4a** reports the fraction of HIVST distributed within each channel by risk group under the base case scenario.

In the phone survey, 52% of respondents receiving HIVST from a FSW channel reported being females, and 48% being males. Our base case scenarios assumed that 90% of females receiving the test through the FSW channels were female sex workers, with the remaining 10% being non-FSW females having multiple sexual partners in the model ("intermediate-risk females"). For the tests received by male participants trough an FSW channel, we assumed that the 67% (2/3) were received by clients of FSW (as these males are likely to have received the tests from a FSW through secondary distribution), and the remaining 33% non-client males that have 3+ partners per year in the model to ("intermediate risk males").

In the phone survey data, 91% of respondents using a test delivered through an MSM channel reported being males. We further assumed that all males receiving HIVST through the MSM channels were MSM and that 66% of them (60% of the total) also had female partners (MSMW). This assumption was based on country-level data on the fraction of MSMW among MSM, as well as the fraction (68%, 334/494) of male phone survey participants from the MSM channel reporting having both male and female partners among those reporting male partners. Out of the 9% of females receiving HIVST from the MSM channel, we assumed that 60% were lower-risk females, 40% were intermediate-risk females, and none were FSW. This was based on the likeliness of MSM having monogamous female partners, whilst in the phone survey, ~55% of females having received HIVST through an MSM channel reported 1 or 2 partners, and 30% reported having 3+ partners.

**Table S4a:** Fraction of tests distributed through the ATLAS FSW and MSM channels across risk groups in the base-case scenarios, based on the ATLAS phone survey (1st stage) data (all ATLAS countries combined)

| | All females (52%) | | | All males (48%) | | | | |
|---|---|---|---|---|---|---|---|---|
| FSW channel | Lower-risk females | Intermediate-risk females | FSW | Lower-risk males | Intermediate-risk males | Clients of FSW | MSMW | MSME |
| | 0% | 5% | 47% | 0% | 16% | 32% | 0% | 0% |
| | All females (9%) | | | All males (91%) | | | | |
| MSM channel | Lower-risk females | Intermediate-risk females | FSW | Lower-risk males | Intermediate-risk males | Clients of FSW | MSMW | MSME |
| | 5% | 4% | 0% | 0% | 0% | 0% | 61% | 30% |

FSW: female sex workers; MSMW: men who have sex with men as well as female partners; MSME: men who have sex with men exclusively.

*HIVST distribution under the sensitivity scenario*

A sensitivity scenario reflected the empirical outcomes of the ATLAS phone survey (1st stage) more accurately without accounting for likely reporting biases more self-tests distributed to non-KP. The **Table S4b** reports the fraction of HIVST distributed within each channel by risk group under the sensitivity scenario.

For the FSW channel, only 40% (250/620) of female phone survey respondents reported having received money in exchange for sex in the past year and were considered as FSW (only 25% of females reported having 3 or more partners). The fraction of tests distributed through the FSW ultimately received by lower risk females was assumed to be 28% as it is the fraction of phone survey female participants (FSW channel) reporting not having partners. Regarding the tests received by males trough an FSW channel, only 24% of males reported having paid for sex in the last year, and were considered to be FSW clients in the model. Around 6% of males reported only having male partners, and 6% both male and female partners, proportions which were used to quantify the fraction of tests from FSW channels given to MSME and MSMW. Around 16% of males survey participants reported having no partners and were considered to correspond to lower-risk populations in the model. The fraction of HIVST from the FSW channel which ended up being received by intermediate risk females and males were calculated from the total number of tests distributed to each gender, and the fraction received by the other groups.

The sensitivity scenario assumed that 91% of individuals receiving HIVST from the MSM channels were males, still based on the phone survey (1$^{st}$ stage). Around 16% (160/997) and 34% (334/997) of men respondents reported having only male partners, and both male and female partners, respectively. The fraction of HIVST received by clients of FSW (12%) was assumed to be half the fraction of men reporting having paid for sex in the past 12 months (236/947/2), as reporting exchanging sex for money between two male partners is not uncommon in the region. The fraction of tests distributed through the MSM channels that were received by lower-risk (heterosexual) male was calculated as the fraction of men reporting no sexual partners (14%, 139/997). Around 24% (22/92) of female participants having received a test through the MSM channel reported having received money for sex in the past year and were considered FSW, whereas 15% reported not having any partner and were considered as low-risk females.

**Table S4b:** Fraction of tests distributed through the ATLAS FSW and MSM channels across risk groups in the sensitivity scenario (presented in the sensitivity analysis), based on the ATLAS phone survey data (all ATLAS countries combined)

| | All females (52%) | | | All males (48%) | | | | |
|---|---|---|---|---|---|---|---|---|
| FSW channel | Lower-risk females | Intermediate-risk females | FSW | Lower-risk males | Intermediate-risk males | Clients of FSW | MSMW | MSME |
| | 15% | 17% | 20% | 8% | 23% | 12% | 3% | 3% |
| | All females (9%) | | | All males (91%) | | | | |
| MSM channel | Lower-risk females | Intermediate-risk females | FSW | Lower-risk males | Intermediate-risk males | Clients of FSW | MSMW | MSME |
| | 2% | 5% | 2% | 13% | 22% | 11% | 30% | 15% |

FSW: female sex workers; MSMW: men who have sex with men as well as female partners; MSME: men who have sex with men exclusively.

*National HIVST scale-up from January 2022*

Within each HIVST distribution scenario, we modelled the scale-up of the WHO objectives of 95% of KP testing twice a year for HIV[145], but only considering HIVST. We calculated the number of HIVST needed to be distributed each year through FSW and MSM channels so that 95% of FSW and 95% of MSM not living with HIV or living with HIV but not on ART (=”eligible” population) receive 2 HIVST each year from 2025. Our analysis assumed a linear scale-up from January 2022 and a plateau from 2025. The size of the target population and number of HIVST to distribute each year were calculated in January 2022 and varied across simulations because sizes of KPs and coverage of ART vary from one simulation to another. Population growth was not accounted for when calculating the size of the target population in future years (but this assumption was tested in our sensitivity analysis), nor was the potential increase in ART coverage over time. The actual coverages in the model are different from the coverages as estimated in January 2022 because 1) large fractions of self-tests distributed through a KP channel were not kept by the target population (e.g. half of HIVST distributed to FSW end up in the hands of males), and 2) the size of the target population may change over time, partly due to the intervention.

As an example, if 50,000 FSW are not living with HIV or living with HIV but not on ART in January 2022, the model will represent a scale-up of HIVST over time so that 95% of 55,000 FSW (size of target population assuming population growth) receive two tests in 2025 through a FSW HIVST distribution channel (0.95 x 55,000 x 2=104,500 HIVST to distribute into the FSW channel).

All model scenarios assumed the same number of tests distributed into each channel each year, except in a specific sensitivity analysis scenario where it increases over time, according to population growth.

*Other HIVST assumptions*

Assumptions regarding HIVST uptake and linkage to care are described in **Figure 1** and **Table S5**. The independent influence of several assumptions on the estimated HIVST scale-up impacts were tested within specific sensitivity analyses.

Our main scenarios assumed that:

- 80% of distributed HIVST were used, based on the experience of STAR[146], although an higher fraction was estimated during a study among FSW in Senegal[139]. This fraction was the same across all population strata.
- 20% (Côte d'Ivoire), 30% (Mali), and 40% (Senegal) of HIVST were "substitute" tests, which were discounted from the number of conventional tests (see specific section on HIVST assumptions) performed during the year by the same population strata. This was based on analysis of subnational-level programmatic data during ATLAS[108], although these reductions were not statistically significant in the analysis. The fraction in Mali was simply calculated as the average between the fractions in Côte d'Ivoire and Senegal.
- Uptake of HIVST within a specific risk/age group combination did not depend on the history of testing nor HIV status, except that PLHIV on ART did not receive HIVST. A sensitivity analysis assumed that people that have never tested were twice more likely to receive a self-test compared to people with the same characteristics who ever had an HIV test.
- Only half of people had a confirmation test following a reactive test, with an average time of two months between the tests, whereas those subsequently receiving a positive confirmation test initiated ART after a month. These assumptions were derived from ATLAS phone survey (2$^{nd}$ stage), which aimed at measuring linkage to care among people experiencing a reactive self-test[110]. A sensitivity analysis looked at the change in estimated impact if the fraction of confirmed reactive tests was increased from 50% to 80%.
- The sensitivity and specificity of the "OraQuick HIV Self-Tests" used during ATLAS were assumed to be 92% and 99%, according to manufacturer data[109]. Real-world studies have estimated lower sensitivity[147,148] when the tests are performed by the intended users, due to variable literacy levels and limited exposure to HIVST. Since we assumed that only 50% of reactive HVIST would be followed by a confirmation test, further reducing the sensitivity had a minimal impact.

**Table S5:** main assumptions of scenarios.

| Parameter | Assumptions | | Comment |
|---|---|---|---|
| | Base-case scenario | Sensitivity analysis scenario) | |
| Number of HIVST kits distributed over 2019-2021 | "Counterfactual": No HIVST distributed<br><br>"ATLAS-only" and "HIVST scale-up": HIVST distribution | | The number of kits distributed each year matches ATLAS programmatic data for FSW and MSM outreach channels (shown in Table S3) |
| Number of HIVST kits distributed over 2022-2038 | "Counterfactual" and "ATLAS-only": No HIVST distributed<br><br>"HIVST scale-up": linear increase up to 2025, then plateau | | "HIVST scale-up": target in 2025 defined as two self-tests to 95% of "eligible" HIVST users (i.e. people without HIV or untreated PLHIV, regardless of their awareness of HIV status) |
| Numbers of kits distributed to KP every year from 2025 account for growth of population size | No i.e. annual number of kits distributed constant over time | Yes, the annual number of kits distributed from 2025 increases by ~3% annually | Based on country-specific UNPD estimates of population growth [4] |
| Fraction of kits distributed through the FSW and MSM channels received by each risk group | Shown in Table S4a: assumption accounting for biases in reported high risk behaviours in the ATLAS phone survey[144], where < 20% of distributed kits end up | Shown in Table S4b: assumption not accounting for biases in the ATLAS phone survey ($1^{st}$ stage) [144], where around 50% of distributed kits end up in the hands of non-KP | Assumption reflecting most plausible secondary distribution of self-tests among ATLAS participants. See Tables S4a,b |

| | | | |
|---|---|---|---|
| | in the hands of non-KP | | |
| Age distribution of people receiving self-tests kits | Matches programmatic data shown in Table S3 | Shown in Figure S2: Matches ATLAS phone survey data among HIVST users[144] | Age distributions are comparable in the two datasets overall, but MSM participating to the phone survey are slightly younger than in the programmatic data, see Figure S2. |
| Fraction of all tests used | 80% (constant across population strata) | 100% | Based on STAR Malawi (80%) [146].<br>Sensitivity assumption based on a survey among FSW in Senegal (~94% HIVST used) [139]. |
| HIVST distributed to PLHIV on ART | No | No | Assumed |
| HIVST uptake (likelihood of receiving a kit) by HIV infection, diagnosis, and treatment status. | Uptake independent of history of testing diagnosis and treatment. | 4 specific uptake scenarios:<br>1) twice higher among those never tested vs ever tested<br>2) PLHIV with a diagnosed infection don't receive tests<br>3) twice higher among PLHIV on ART vs others<br>4) twice higher among PLHIV with an undiagnosed infection vs others | Uptake assumptions modelled as independent relative risks all = 1 in the base-case. |
| Fraction of reactive HIVST of undiagnosed PLHIV followed by a confirmatory test | 50% | 80% | Base-case assumption derived from ATLAS phone survey data[110].<br>Sensitivity analysis assumption assumes improvements in use of HIVST (improved kit user manual, experience with COVID self-tests kits) |
| Time from reactive to confirmatory test (for those eventually confirming) and from confirmatory test to ART initiation | 2 months and 1 month | | ATLAS phone survey data (2nd stage)[110] |

| | | | |
|---|---|---|---|
| Test substitution fraction: HIVST substitute/replace conventional tests | 20% (Côte d'Ivoire)<br>30% (Mali)<br>40% (Senegal) | No substitution | Analysis of subnational programmatic data in Côte d'Ivoire and Senegal[108] during ATLAS. Fraction for Mali was assumed to be the average of the estimates from the two other countries. Note that the estimated levels of substitution were not statistically significant. |
| HIVST sensitivity/specificity | 92% / 99% | 100% | Manufacturer data [109]. |

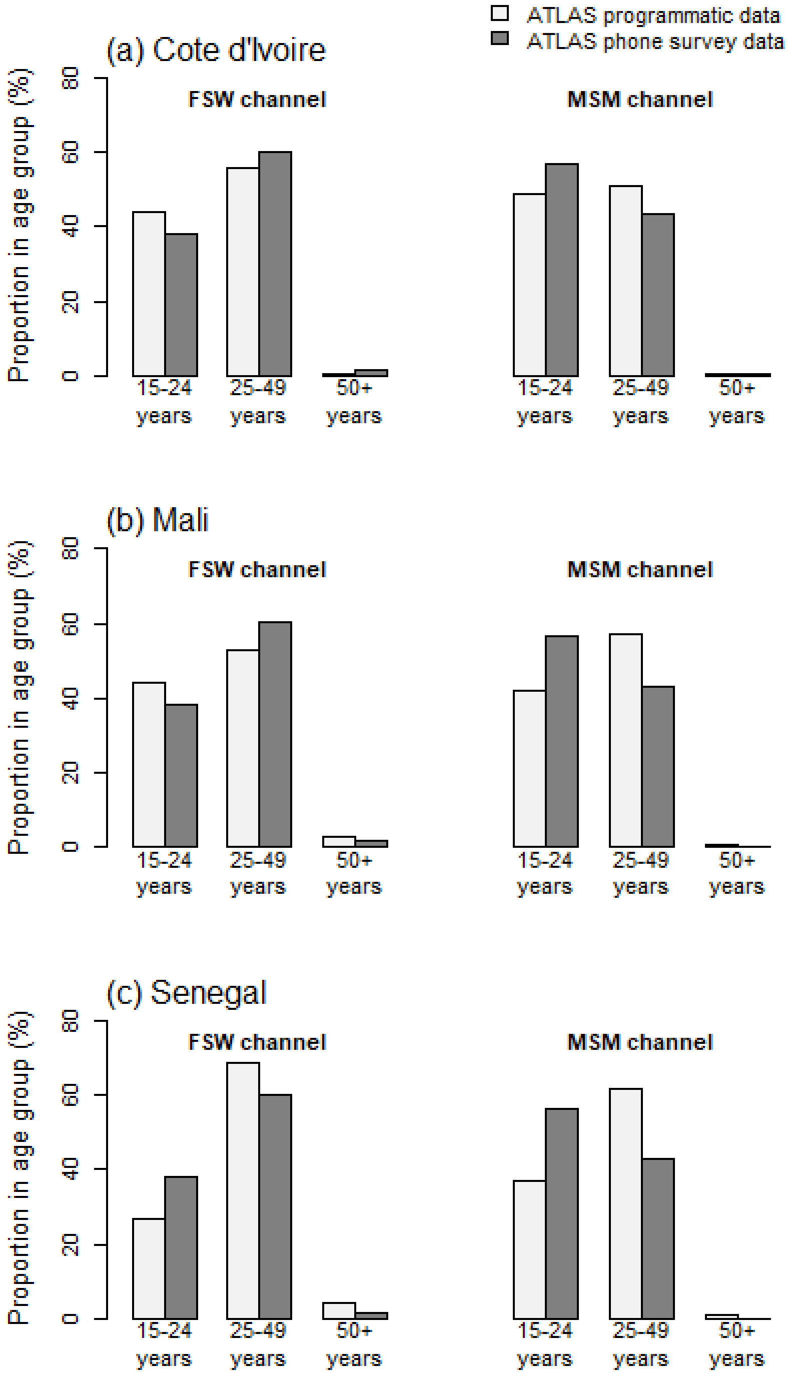

**Figure S2:** Proportions of HIV self-tests distributed through FSW and MSM outreach to different age groups in the programmatic data (white bars, base-case scenarios) in each ATLAS country, and in the ATLAS phone survey (stage 1) data (dark grey bars, sensitivity analysis scenarios) all countries combined.

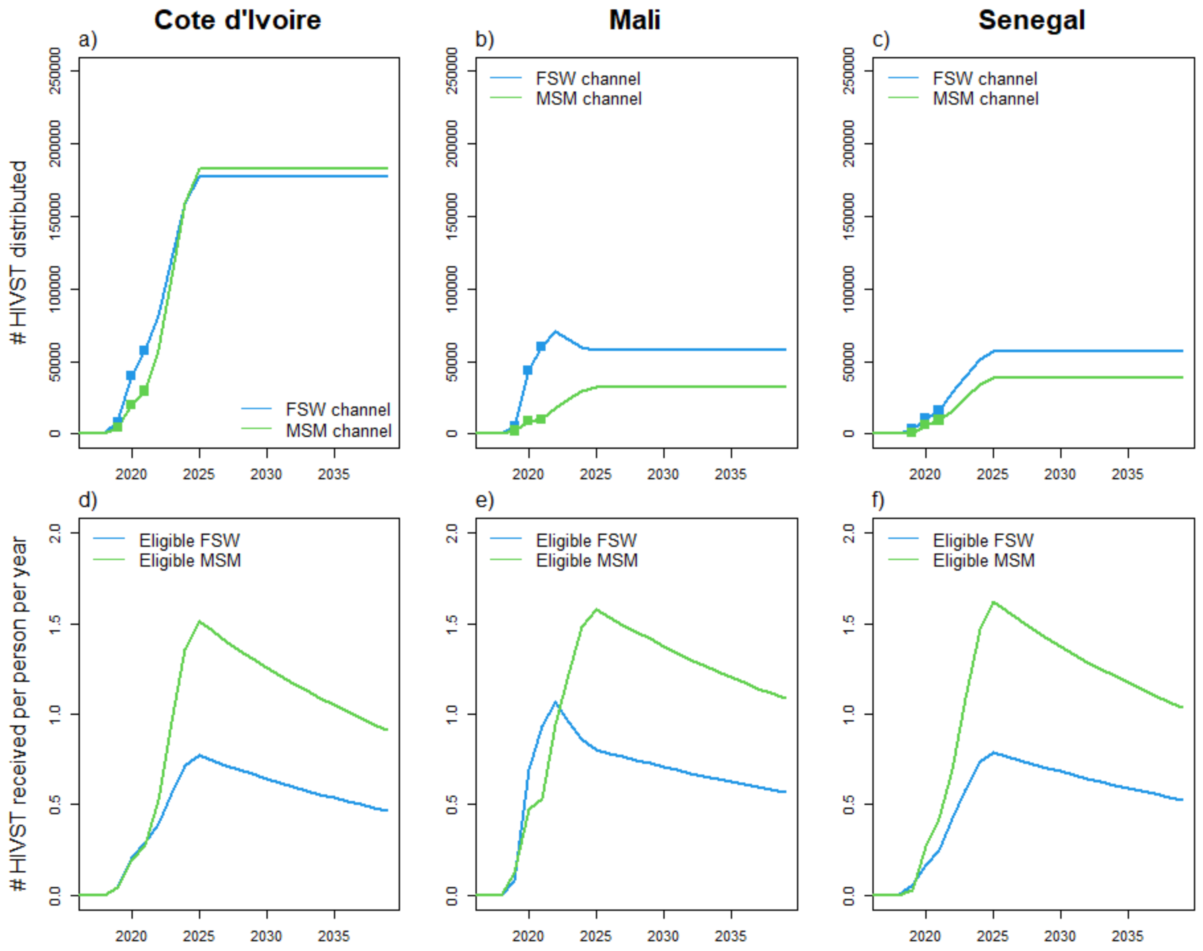

**Figure S3:** HIVST distribution from mid-2019 in each ATLAS country. Panels a-c) Number of HIVST distributed by ATLAS over 2019-2021 and over 2022-2038 during a hypothetical scale-up of HIVST through female sex workers (blue), and men who have sex with men (green) outreach channels. Panels d-f) Median number of HIVST received each year per FSW (blue) and MSM (green) not living with HIV or living with an untreated HIV infection. The decrease in numbers per capita on panels d-f) from 2025 is because the population is assumed to grow at a constant rate whereas the number of kits distributed over time is constant.

### Results: model fits (Côte d’Ivoire)

#### Demography

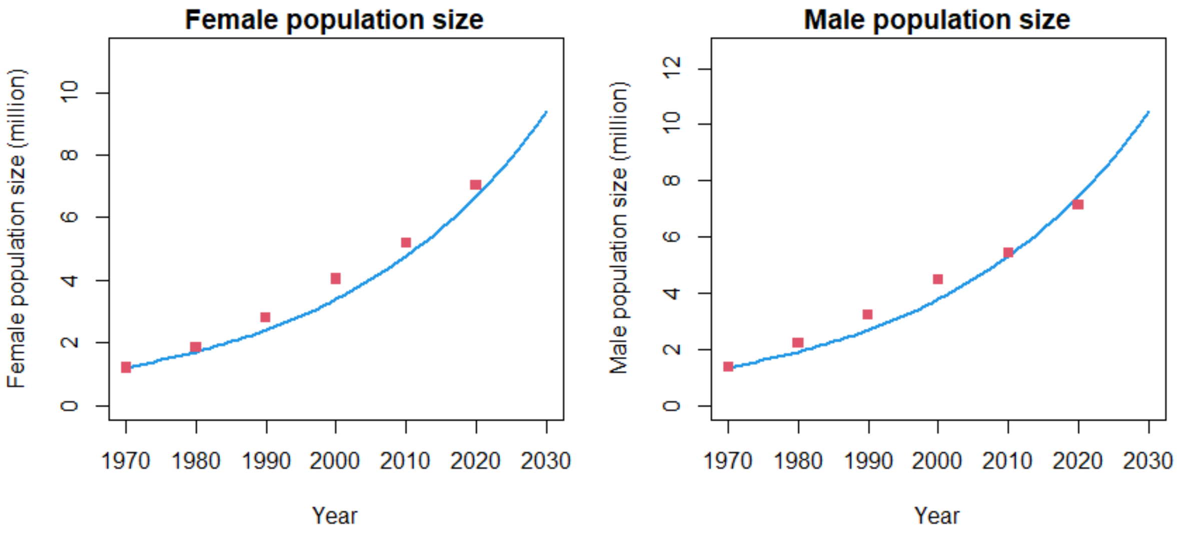

**Figure S4a:** Côte d’Ivoire model fitting to the size of the female and male population aged 15-59 years old over time. Blue curves represent model estimates and red squares the estimates from UNPD[4].

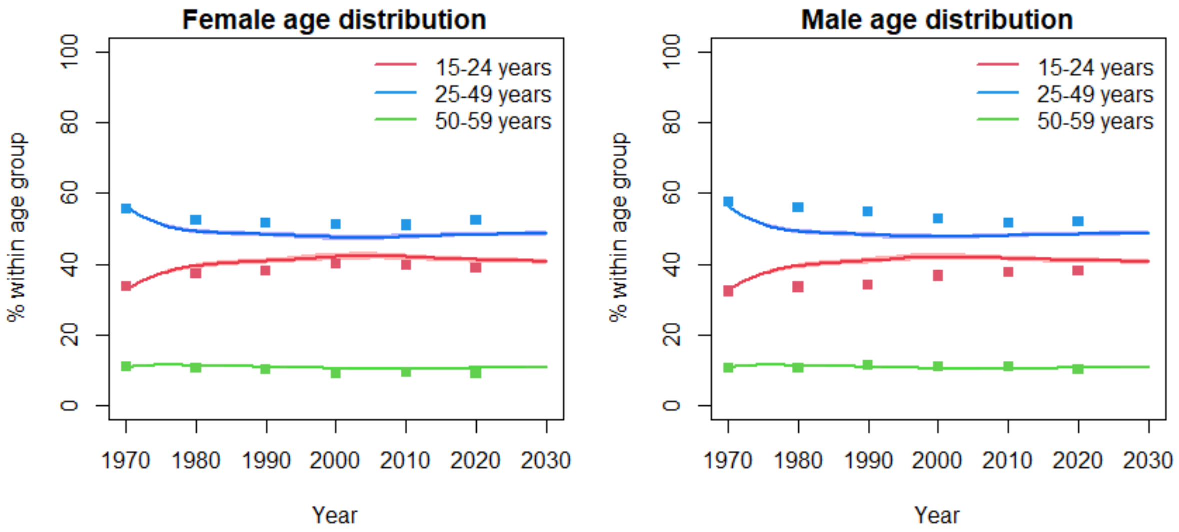

**Figure S4b:** Côte d’Ivoire model fitting to the age distribution of the female and male population aged 15-59 years old over time. Curves represent model estimates and the squares the estimates from UNPD.

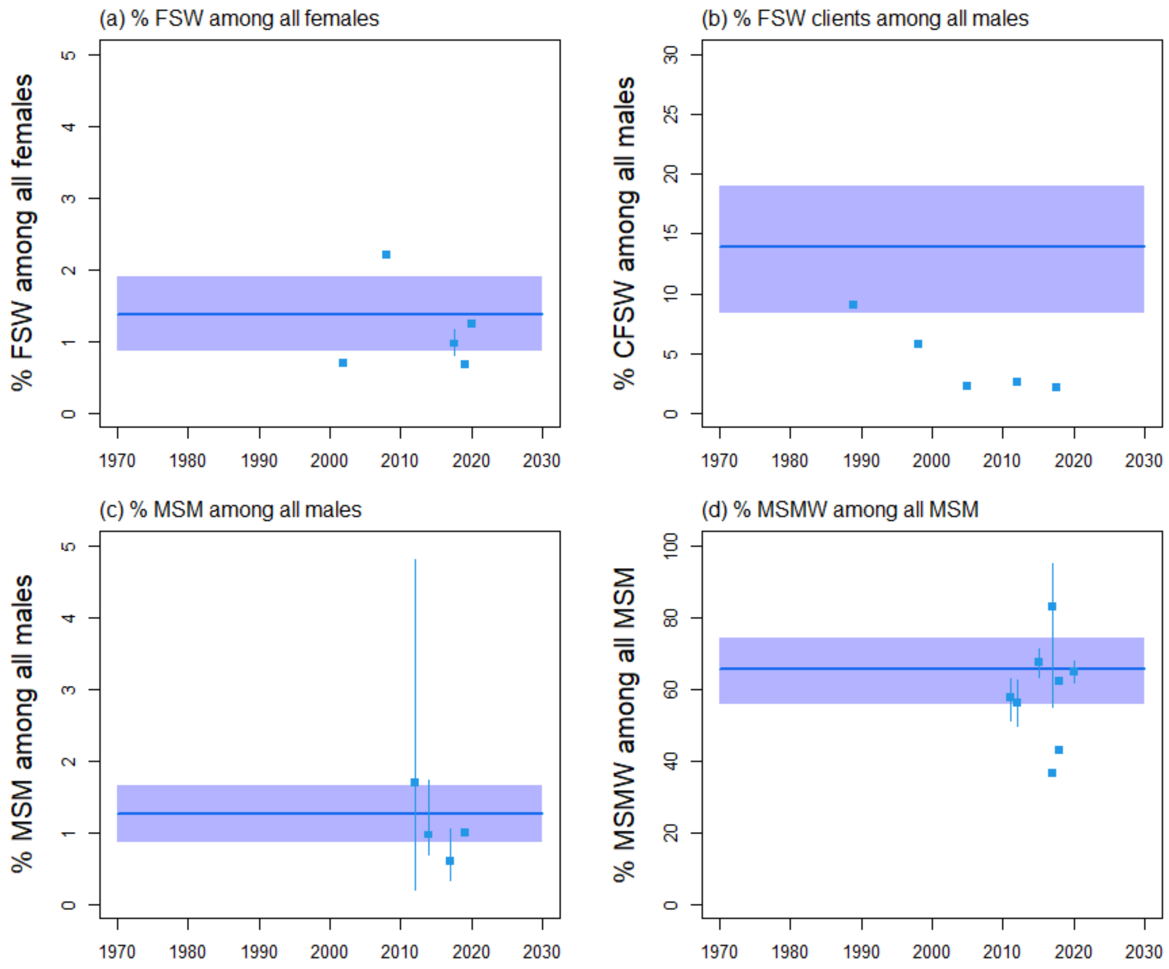

**Figure S4c:** Côte d'Ivoire model fitting to the size of key populations and their clients, with fractions of a) FSW among all females aged 15-59 years, b) FSW clients among all males aged 15-59 years, c) MSM among all males aged 15-59 years, and d) MSMW (men who have sex with men and women) among all MSM aged 15-59 years old over time. Blue curves and shades represent median and 90% UI (5th$^{th}$ and 95$^{th}$ percentiles across model fits), whereas squares and intervals represent empirical estimates (with 95% CI). Estimates in panel b) were reported from household surveys and were only use for comparison, whereas the FSW clients population size in the model was calculated using the multiplier method as in[1].

#### HIV epidemiology

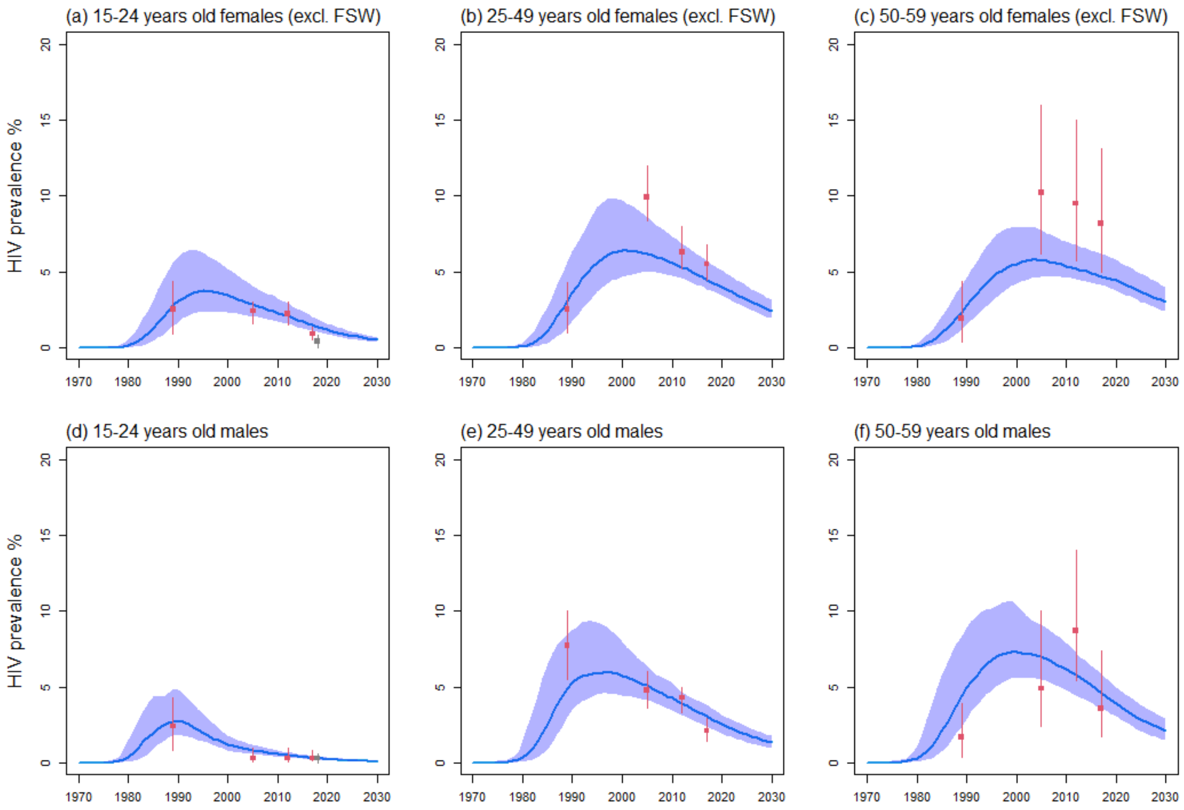

**Figure S4d:** Côte d'Ivoire model fitting to the HIV prevalence among all females aged a) 15-24, b) 25-49, and c) 50-59 years old (excluding FSW), as well as all males aged d) 15-24, e) 25-49 years, and f) 50-59 years old. Blue curves and shades represent median and 90% UI (5$^{th}$ and 95$^{th}$ percentiles across model fits), red squares and intervals represent empirical estimates used for model fitting (with 95% CI), and the grey square and intervals in panel a) are estimates from a recent survey about violence against children and youth in Côte d'Ivoire[112] included for comparison (not used to fit the model).

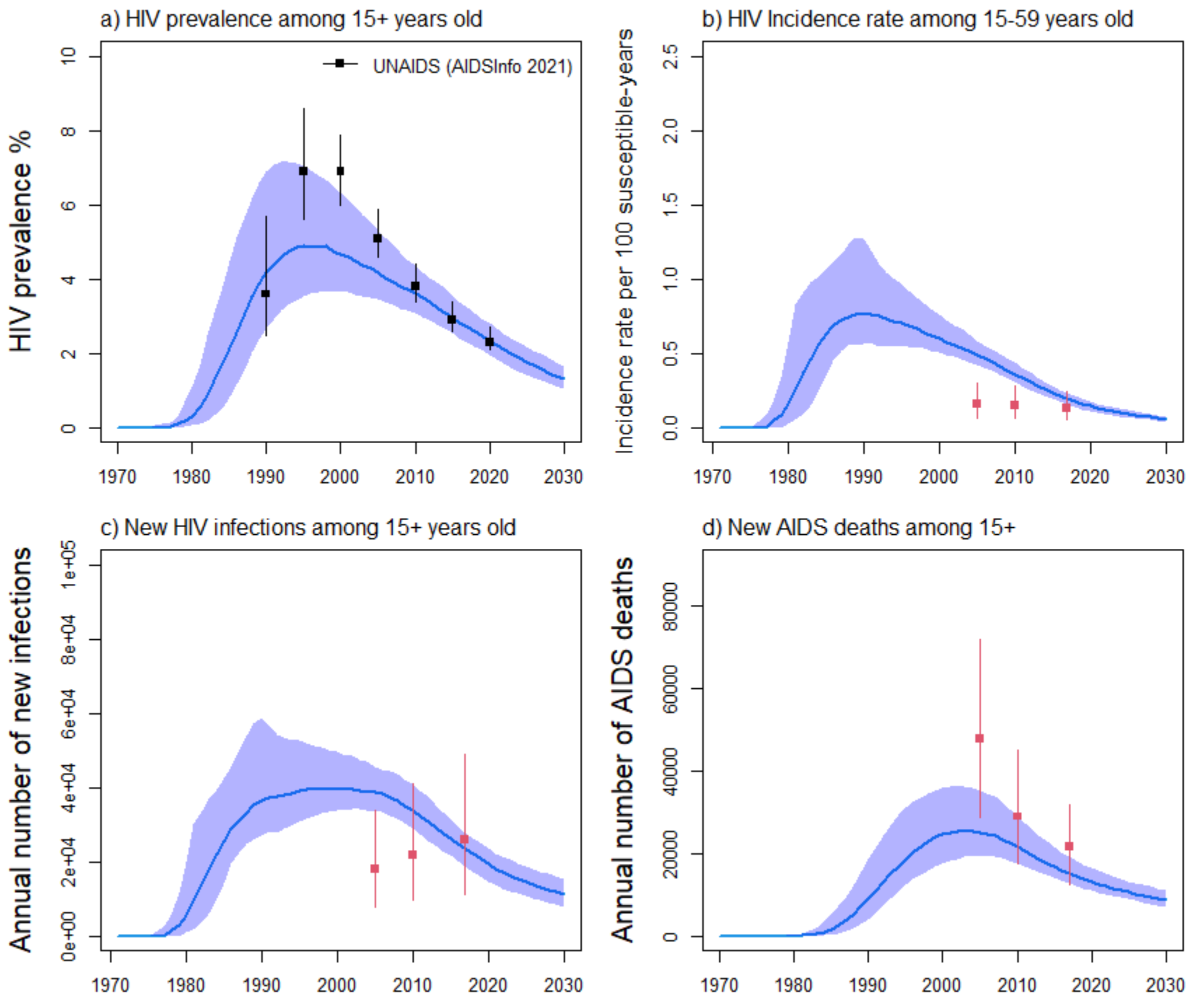

**Figure S4e:** Estimates of HIV prevalence among a) all adults aged over 15 years old, fits to b) HIV incidence rate per susceptible, c) annual number of new HIV infections and d) annual HIV-related deaths in Côte d'Ivoire from UNAIDS Data 2018. Blue curves and shades represent median and 90% UI (5$^{th}$ and 95$^{th}$ percentiles across model fits), red squares and intervals represent empirical estimates used for model fitting (with 95%CI), whereas the dark squares and intervals in panel a) represent estimates from UNAIDS used for comparison.

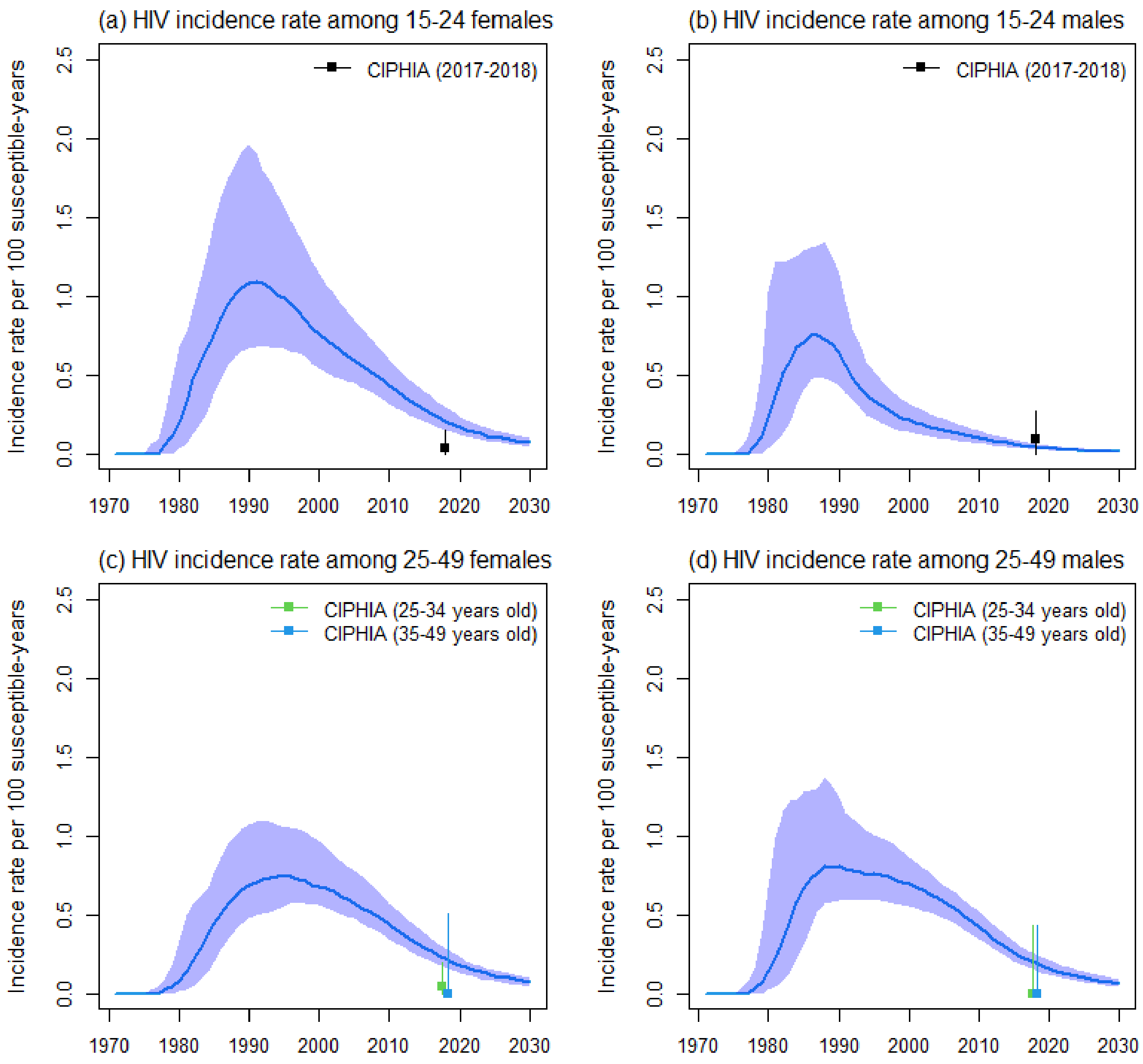

**Figure S4f:** Comparison of estimates of HIV incidence in the Côte d'Ivoire PHIA survey among a) females aged 15-24 years old, b) males aged 15-24 years old, c) females aged 25-49 years old, d) males aged 25-49 years old. Blue curves and shades represent median and 90% UI ($5^{th}$ and $95^{th}$ percentiles across model fits), whereas squares and intervals represent empirical estimates from the PHIA survey (with 95%CI).

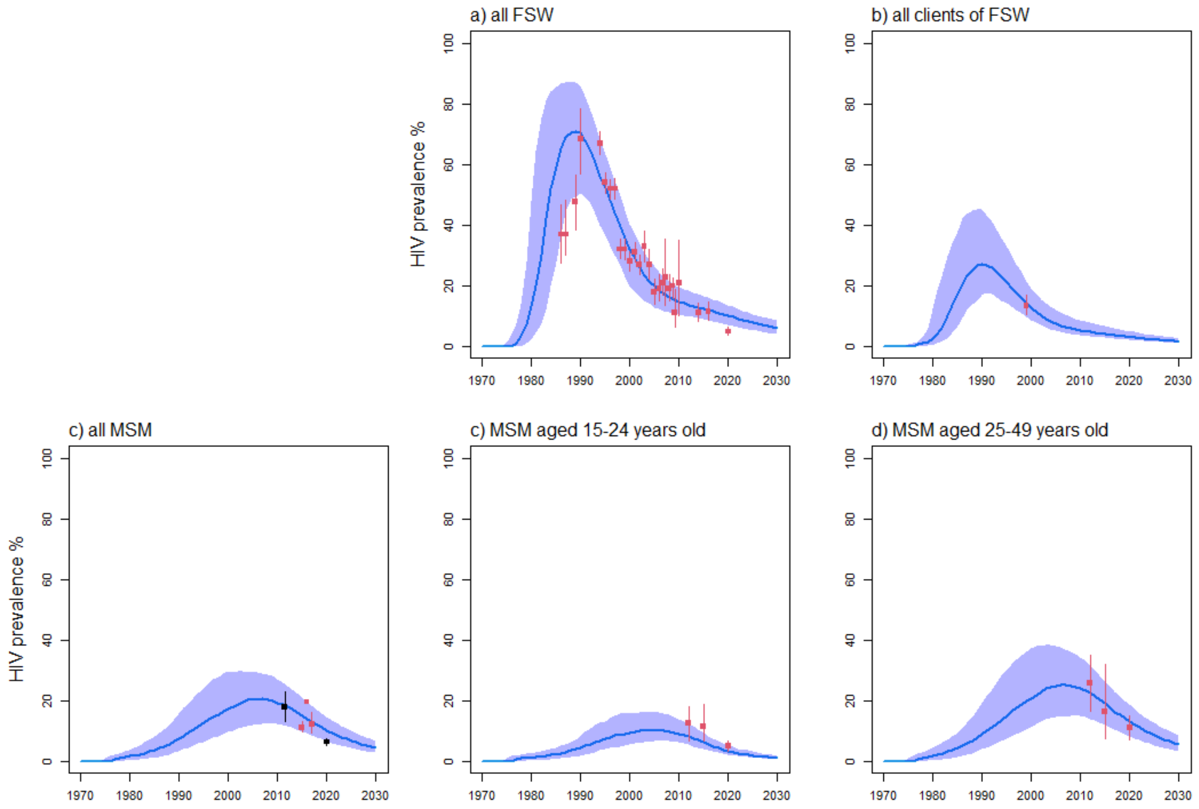

**Figure S4g:** Côte d'Ivoire model fitting to HIV prevalence estimates among all a) FSW, b) clients of FSW, c) MSM, as well as c) MSM aged 15-24 years old, and d) aged 25-49 years old. Blue curves and shades represent median and 90% UI (5[th] and 95[th] percentiles across model fits), red squares and intervals represent empirical estimates used for model fitting (with 95%CI), whereas dark estimates in panel c) represent estimates fitted among MSMW and MSME separately (see Figure S4h).

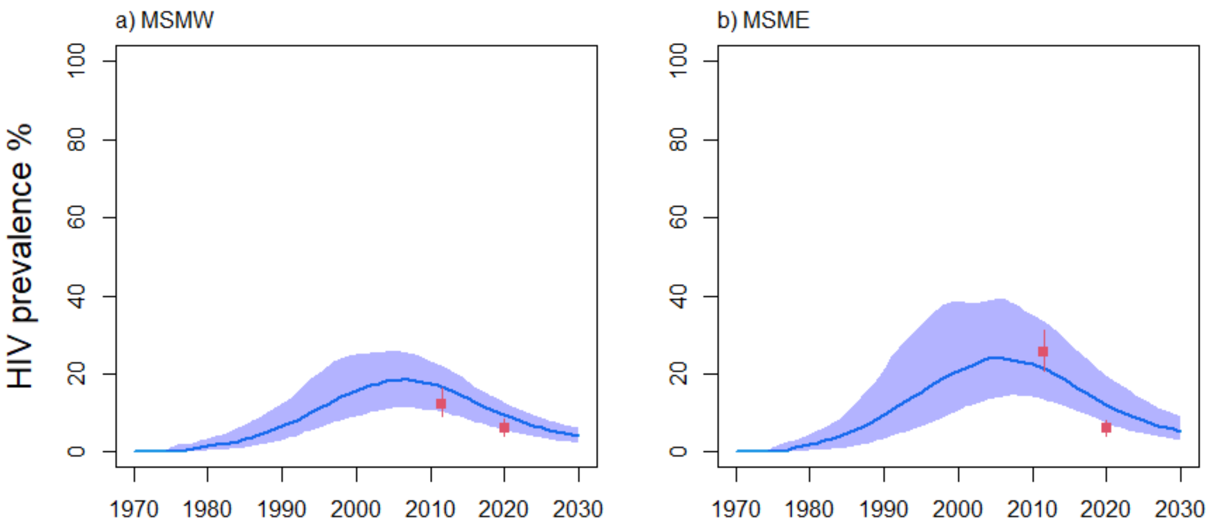

**Figure S4h:** Côte d'Ivoire model fitting to HIV prevalence estimates among all a) MSMW (men having sex with both men and women) and b) MSME (men having sex with another men) MSM. Blue curves and shades represent median and 90% UI (5th and 95th percentiles across model fits), whereas red squares and intervals represent empirical estimates (with 95%CI).

#### HIV treatment cascade

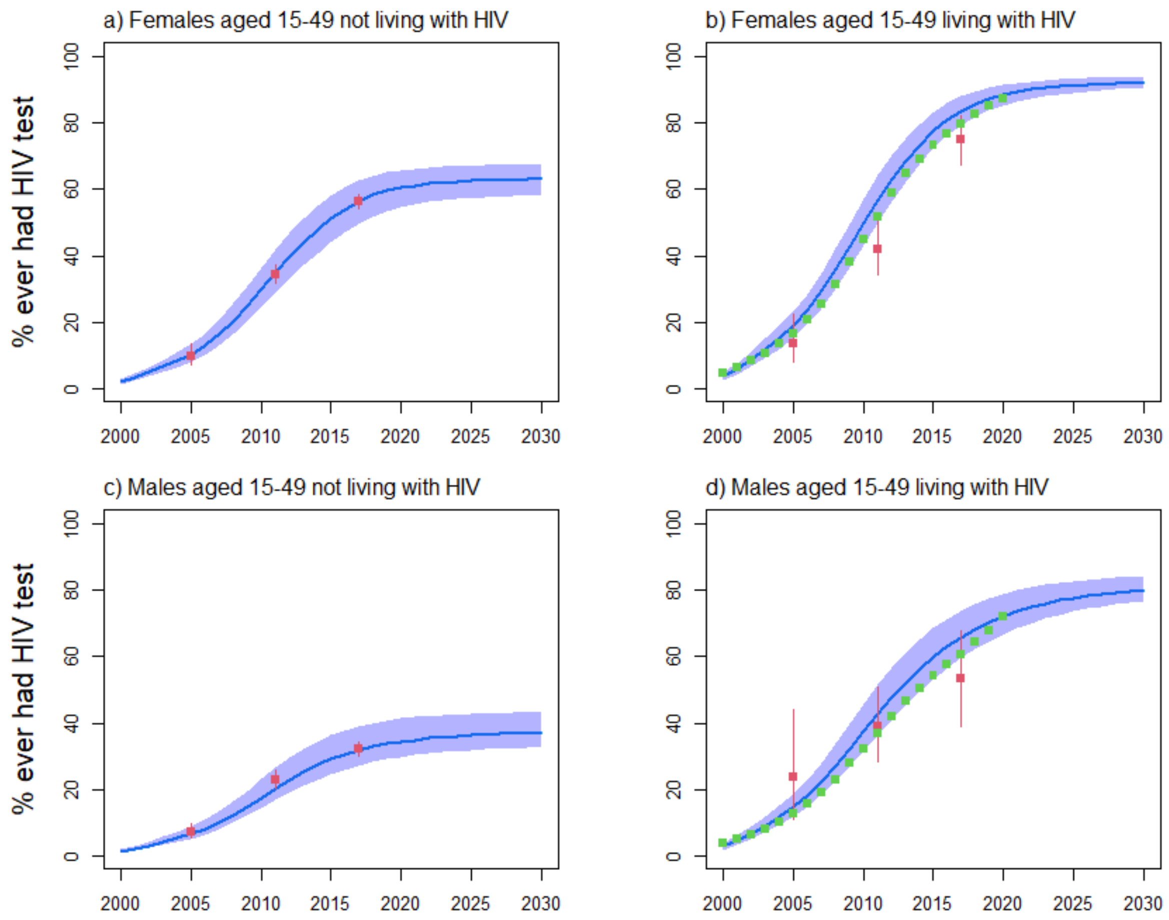

**Figure S4i:** Côte d'Ivoire model fitting to the fraction ever having tested for HIV among all females aged 15-49 years old a) not living with HIV, and b) living with HIV, and males aged 15-49 years old c) not living with HIV, and d) living with HIV. Blue curves and shades represent median and 90% UI (5th and 95th percentiles across model fits), red squares and intervals represent empirical estimates used for model fitting (with 95%CI), whereas green squares represent estimates from UNAIDS Shiny90 which were only used for comparison.

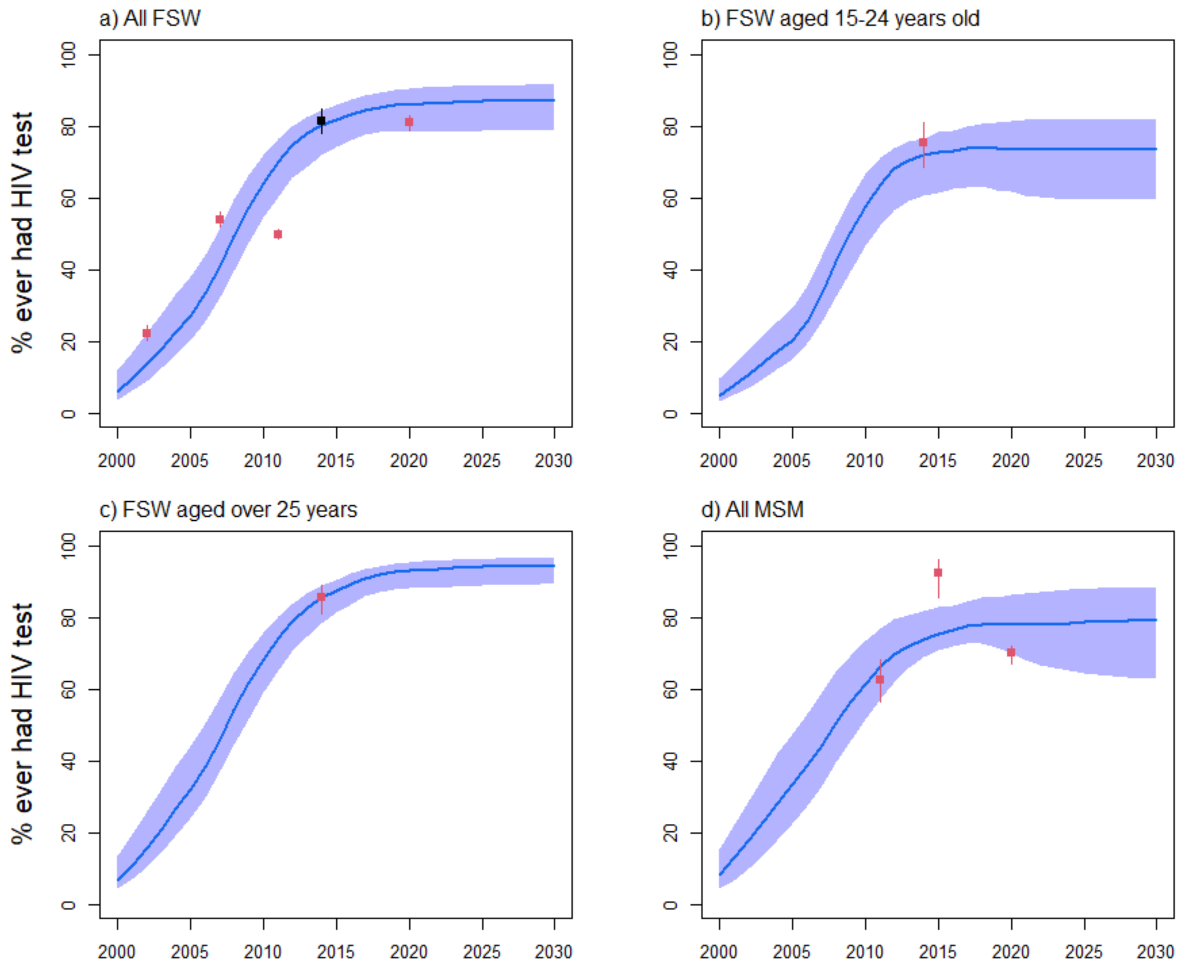

**Figure S4j:** Côte d'Ivoire model fitting to the fraction ever having tested for HIV among a) all FSW, b) FSW aged 15-24 years, c) FSW aged 25-49 years, and d) MSM. Blue curves and shades represent median and 90% UI (5th and 95th percentiles across model fits), red squares and intervals represent empirical estimates used for model fitting (with 95%CI), whereas dark squares in panel a) represent overall fraction from study outcomes shown in panels b) and c).

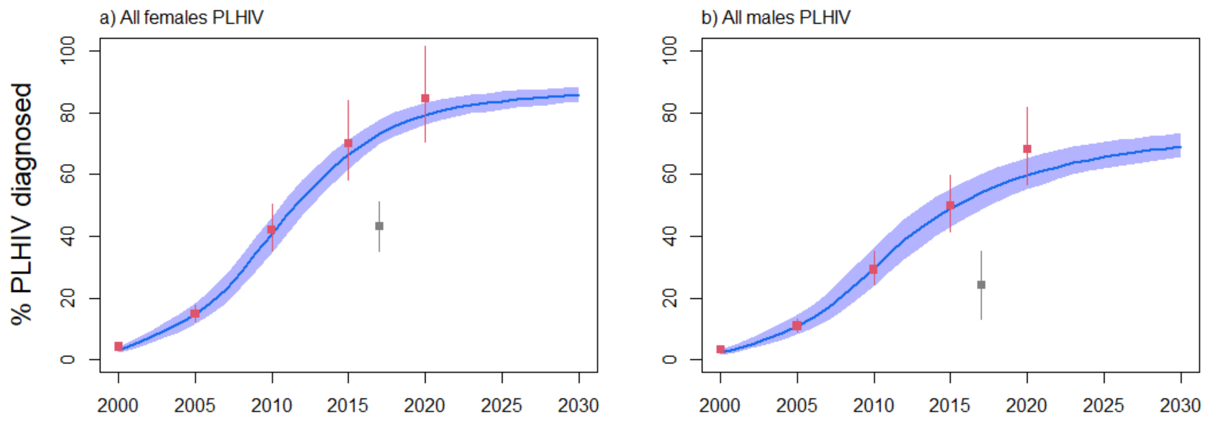

**Figure S4k:** Côte d'Ivoire model fitting to the fraction of a) all females living with HIV and b) all males living with HIV diagnosed (aware of their status). Blue curves and shades represent median and

90% UI (5th and 95th percentiles across model fits), red squares and intervals represent estimates from UNAIDS Shiny90 used for model fitting[126], whereas grey squares and intervals represent estimates from PHIA[31], which were underestimates because this fraction of people diagnosed (reporting being aware of their positive status) lower than the fraction of PLHIV having traces of ARV drugs in their blood in the survey.

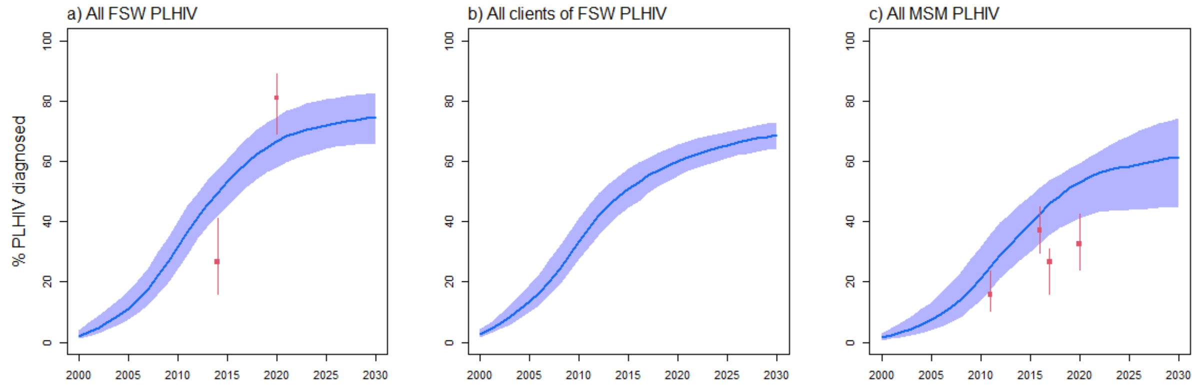

**Figure S4l:** Côte d’Ivoire model fitting to the fraction of a) all FSW living with HIV, b) all male clients of FSW living with HIV, and c) all MSM living with HIV diagnosed. Blue curves and shades represent median and 90% UI (5th and 95th percentiles across model fits), whereas red squares and intervals represent estimates from empirical surveys (with 95%CI).

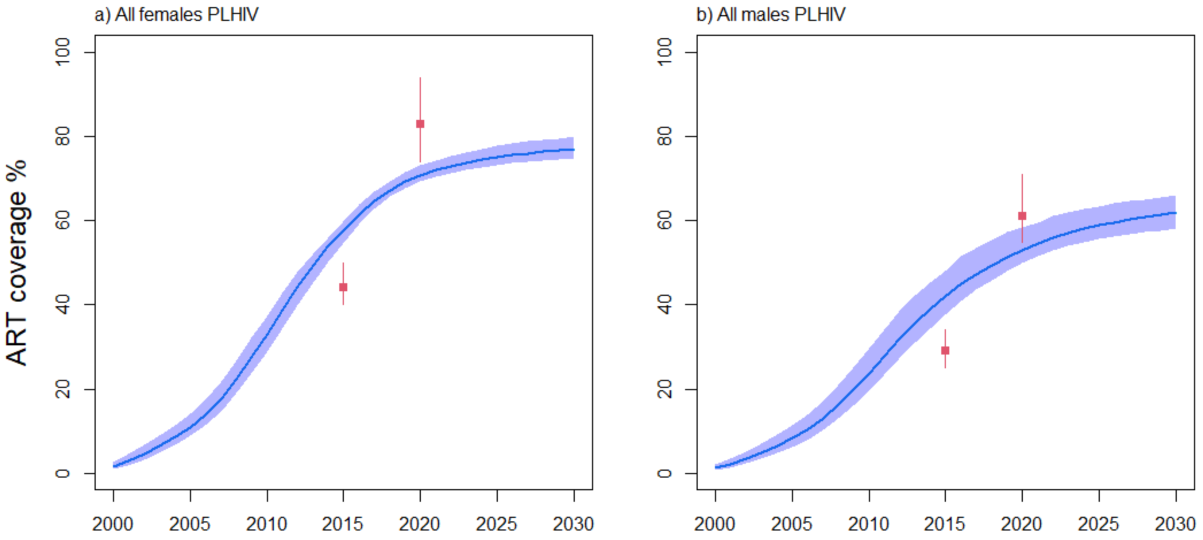

**Figure S4m:** Côte d’Ivoire model fitting to ART coverage among a) all females and b) all males aged 15-59 years old living with HIV. Blue curves and shades represent median and 90% UI (5thth and 95th percentiles across model fits), whereas red squares and intervals represent estimates from UNAIDS (with 95%CI).

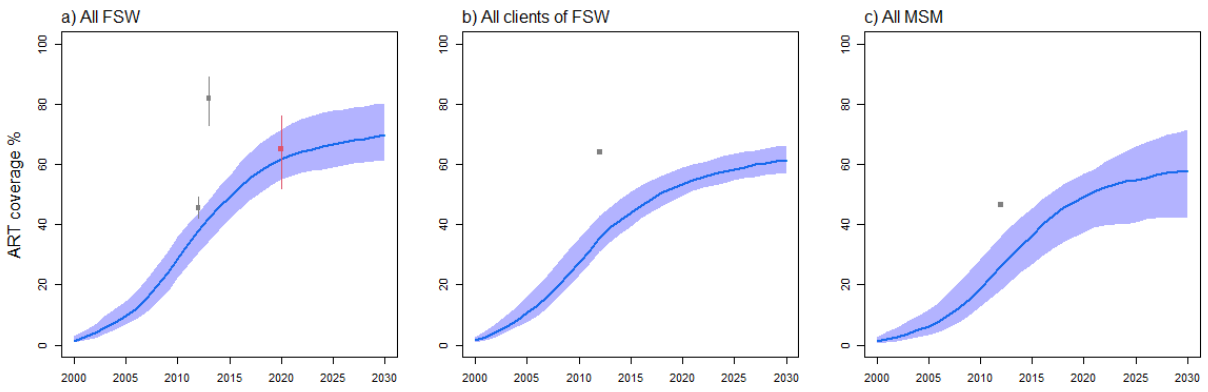

**Figure S4n:** Côte d'Ivoire model fitting to ART coverage among a) all FSW, b) all male clients of FSW, and c) all MSM living with HIV. Blue curves and shades represent median and 90% UI (5th and 95th percentiles across model fits), whereas red squares and intervals represent empirical estimates from local surveys (with 95%CI)[118]. Estimates in grey represent estimates from an STI clinic[115], which were assumed to be overestimates and not included for model fitting but shown for comparison.

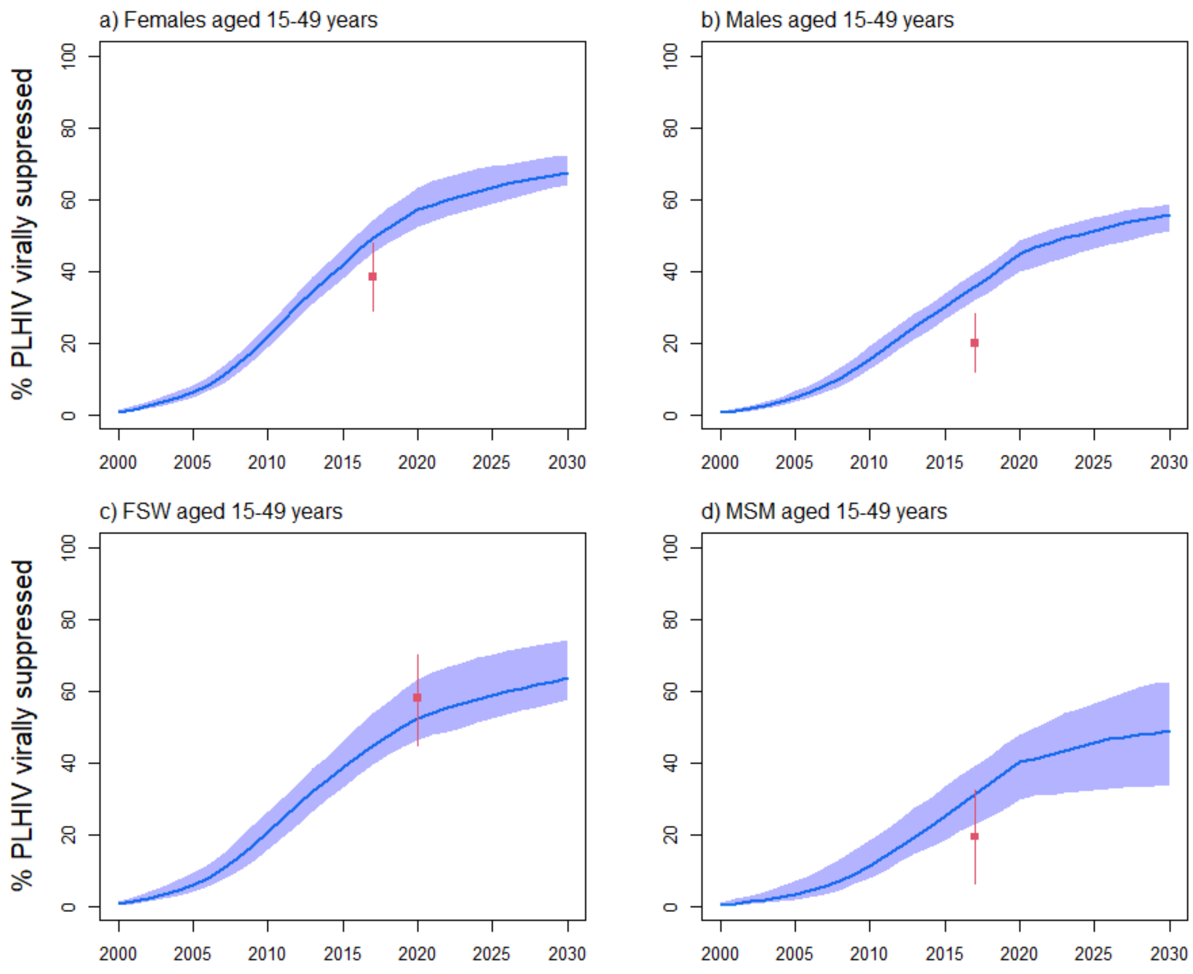

**Figure S4o:** Côte d'Ivoire model fitting to HIV viral load suppression coverage among a) all females, b) all males, c) all FSW, and d) all MSM living with HIV aged 15-49 years. Blue curves and shades represent median and 90% UI (5th and 95th percentiles across model fits), whereas red squares and intervals represent empirical estimates from local surveys.

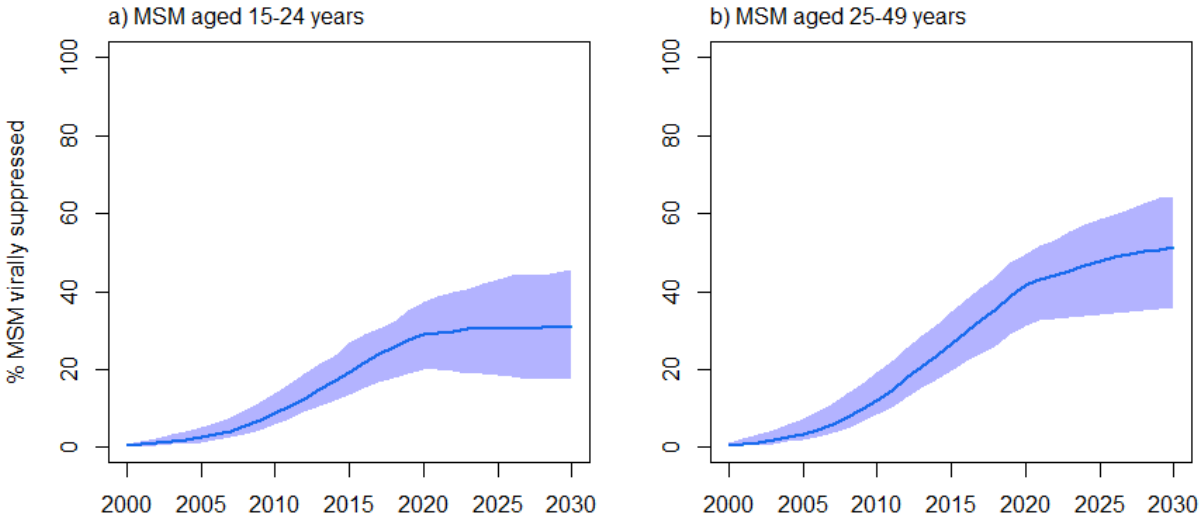

**Figure S4p:** Côte d'Ivoire model fitting to HIV viral load suppression coverage among MSM aged a) 15-24 years and b) 25-49 years living with HIV. Blue curves and shades represent median and 90% UI (5th and 95th percentiles across model fits). No data was available.

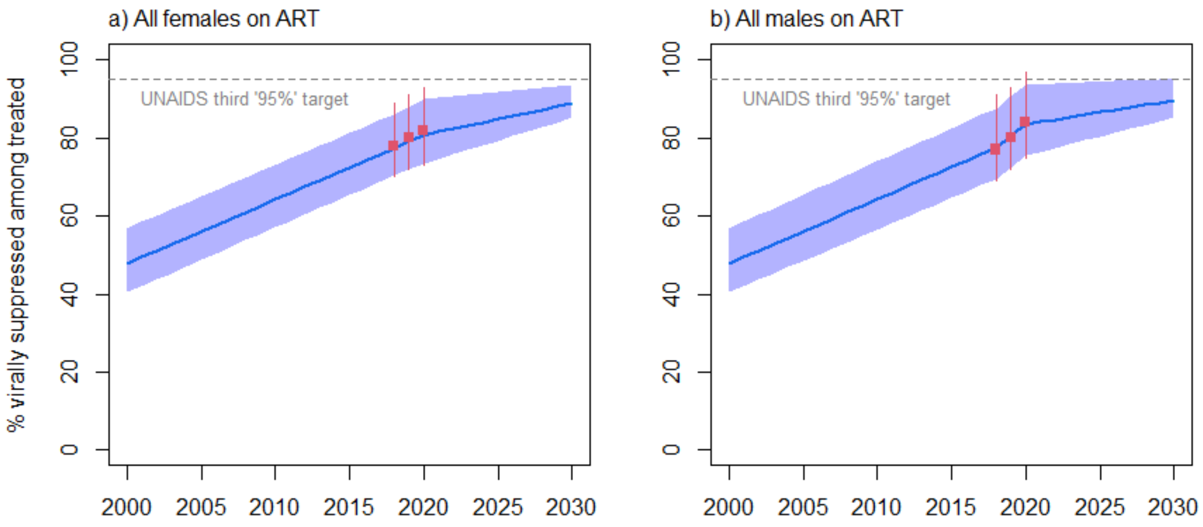

**Figure S4q:** Côte d'Ivoire model fitting to fractions of a) females and b) males PLHIV on ART which are virally suppressed (the third UNAIDSs "95%" indicator) over time. Blue curves and shades represent median and 90% UI (5th and 95th percentiles across model fits), whereas red squares and intervals represent estimates from UNAIDS used as parameters and not at fitting targets. The grey dashed line corresponds to the UNAIDS's third "95%" target whereby 95% of PLHIV on ART should be virally suppressed in 2025.

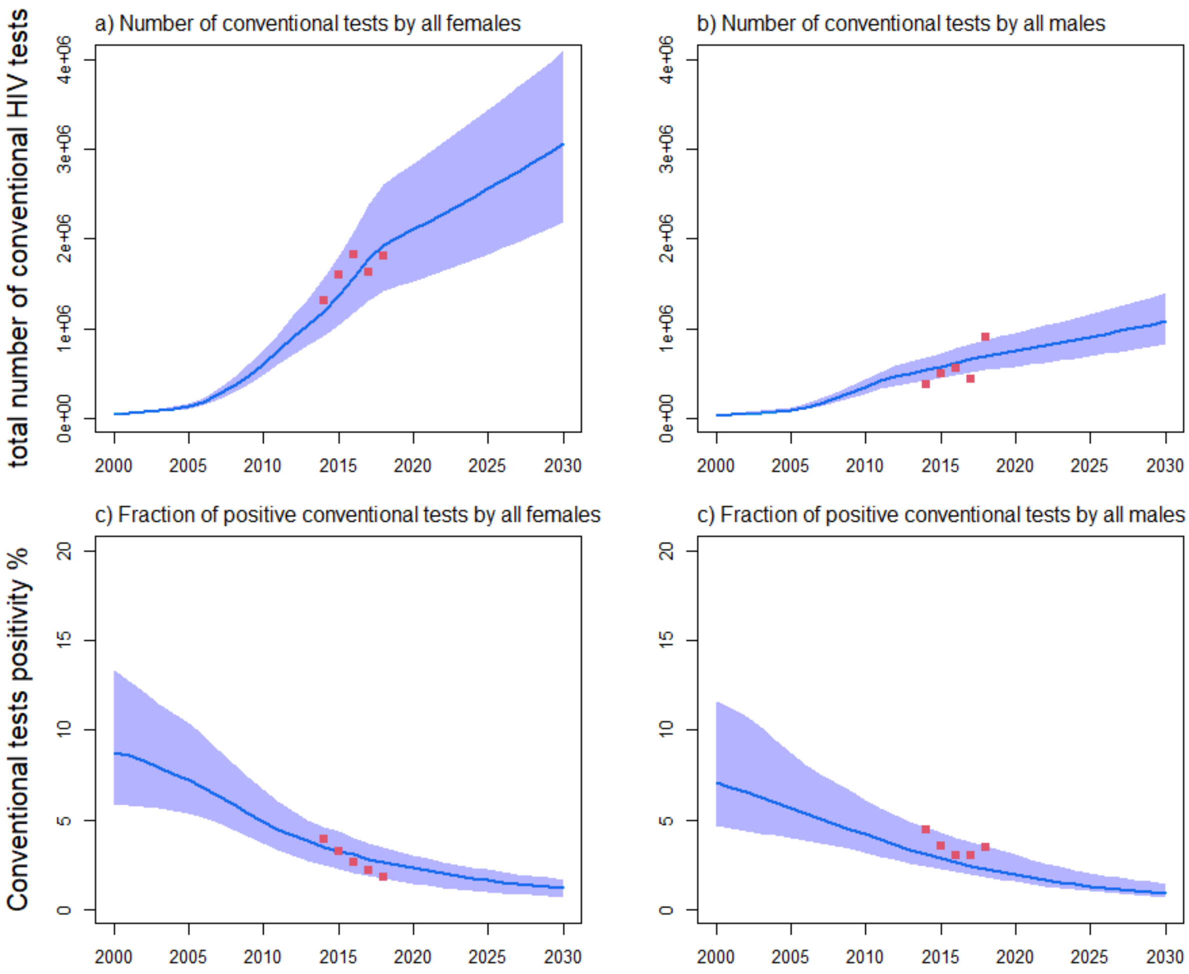

**Figure S4r:** Côte d'Ivoire model fitting to programmatic data[67] on the total number of conventional tests among a) all females and b) all males. Proportions of positive conventional tests among c) all females, and d) all males. Blue curves and shades represent median and 90% UI (5th and 95th percentiles across model fits), whereas red squares represent programmatic data communicated by UNAIDS.

### Results: model fits (Mali)

#### Demography

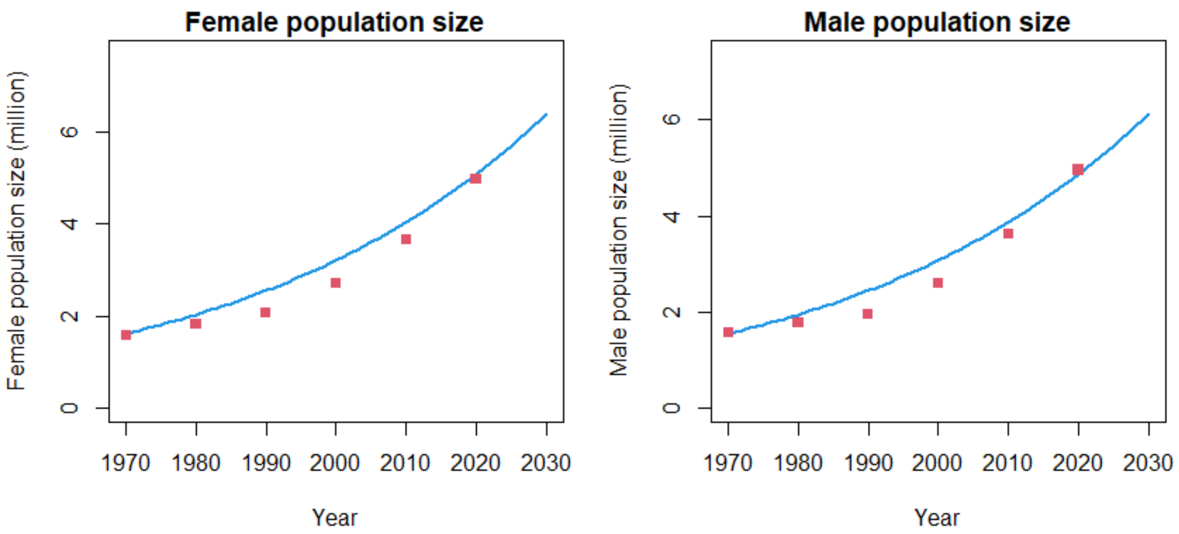

**Figure S5a:** Mali model fitting to the size of the female and male population aged 15-59 years old over time. Blue curves represent model estimates and red squares the estimates from UNPD[4].

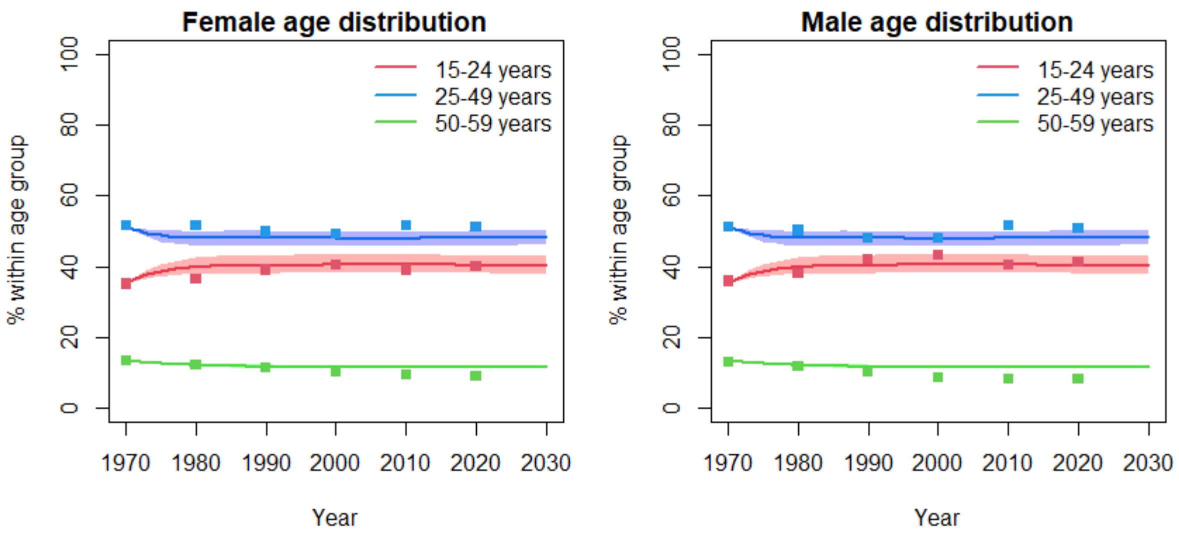

**Figure S5b:** Mali model fitting to the size of the female and male population aged 15-59 years old over time. Blue curves represent model estimates and red squares the estimates from UNPD[4].

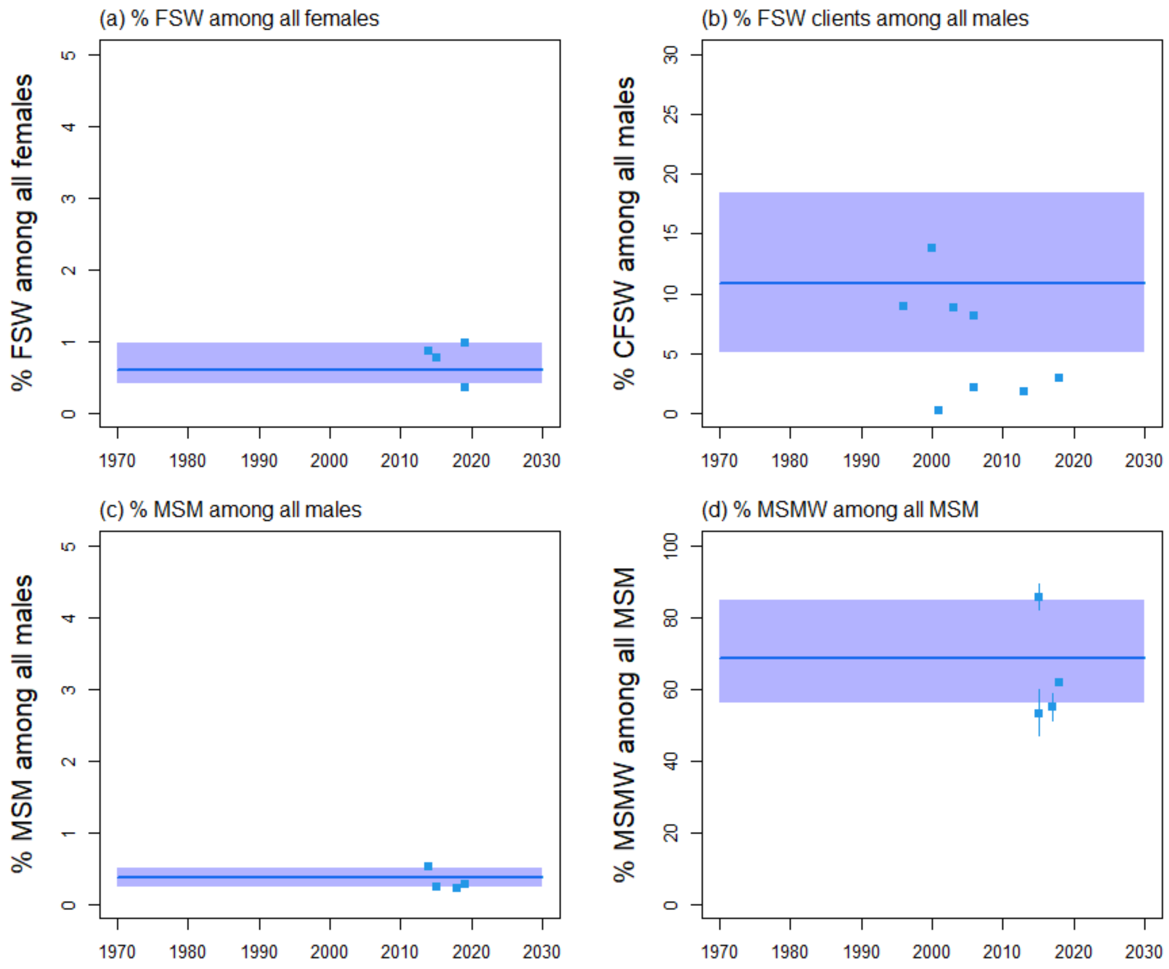

**Figure S5c:** Mali model fitting to the size of key populations and their clients, with fractions of a) FSW among all females aged 15-59 years, b) FSW clients among all males aged 15-59 years, c) MSM among all males aged 15-59 years, and d) MSMW (men who have sex with men and women) among all MSM aged 15-59 years old over time. Blue curves and shades represent median and 90% UI (5th[th] and 95[th] percentiles across model fits), whereas squares and intervals represent empirical estimates (with 95% CI). Estimates in panel b) were reported from household surveys and were only use for comparison, whereas the FSW clients population size in the model was calculated using the multiplier method as in[1].

#### HIV epidemiology

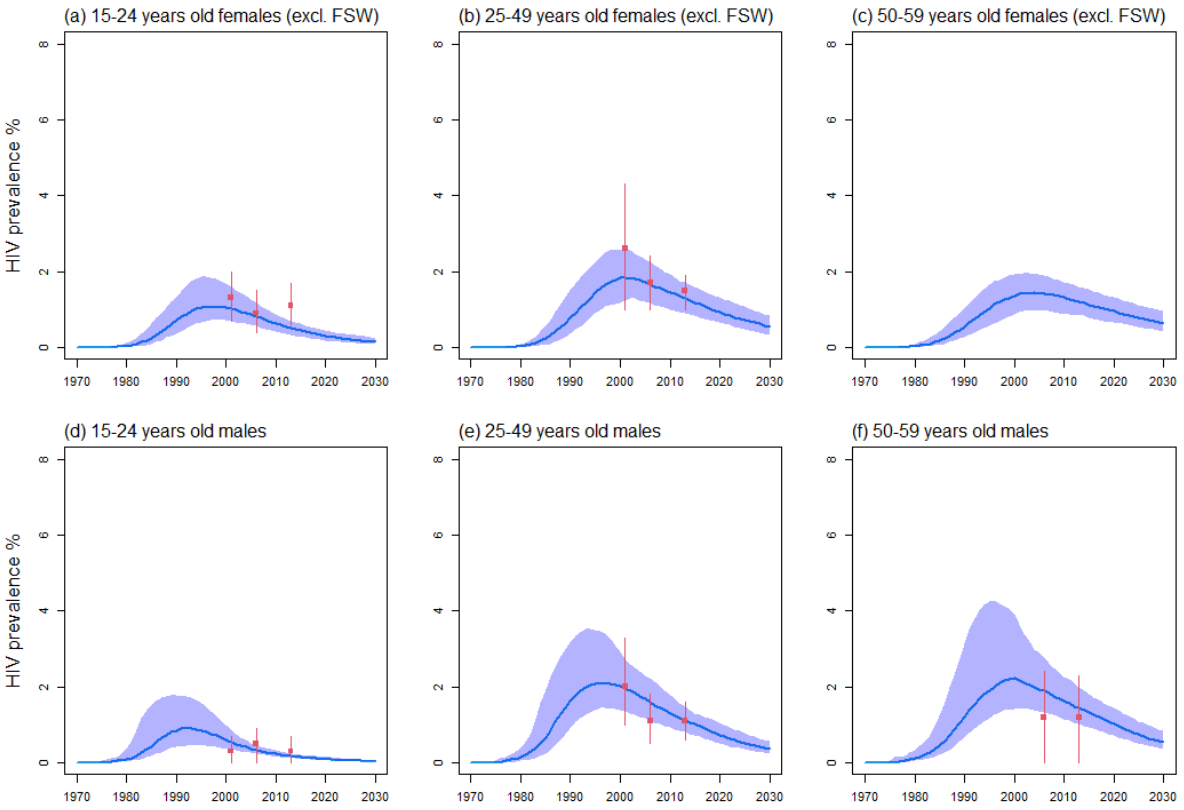

**Figure S5d:** Mali model fitting to the HIV prevalence among all females aged a) 15-24, b) 25-49, and c) 50-59 years old (excluding FSW), as well as all males aged d) 15-24, e) 25-49 years, and f) 50-59 years old. Blue curves and shades represent median and 90% UI (5$^{th}$ and 95$^{th}$ percentiles across model fits), red squares and intervals represent empirical estimates used for model fitting (with 95% CI).

.

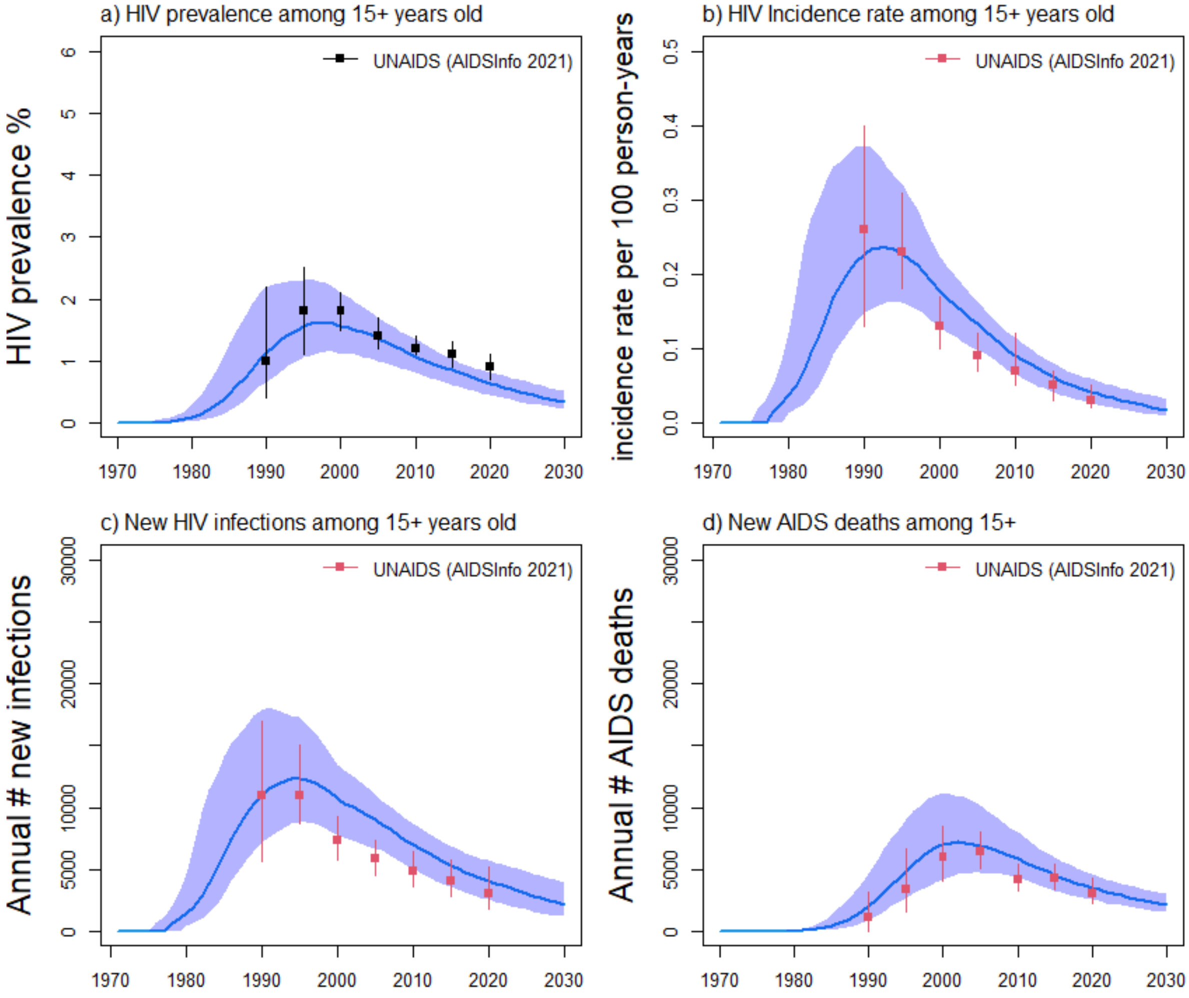

**Figure S5e:** Estimates of HIV prevalence among a) all adults aged over 15 years old, fits to b) HIV incidence rate, c) annual number of new HIV infections and d) annual HIV-related deaths in Mali from UNAIDS. Blue curves and shades represent median and 90% UI (5$^{th}$ and 95$^{th}$ percentiles across model fits), red squares and intervals represent empirical estimates used for model fitting, whereas the dark squares and intervals in panel a) represent estimates from UNAIDS only used for comparison.

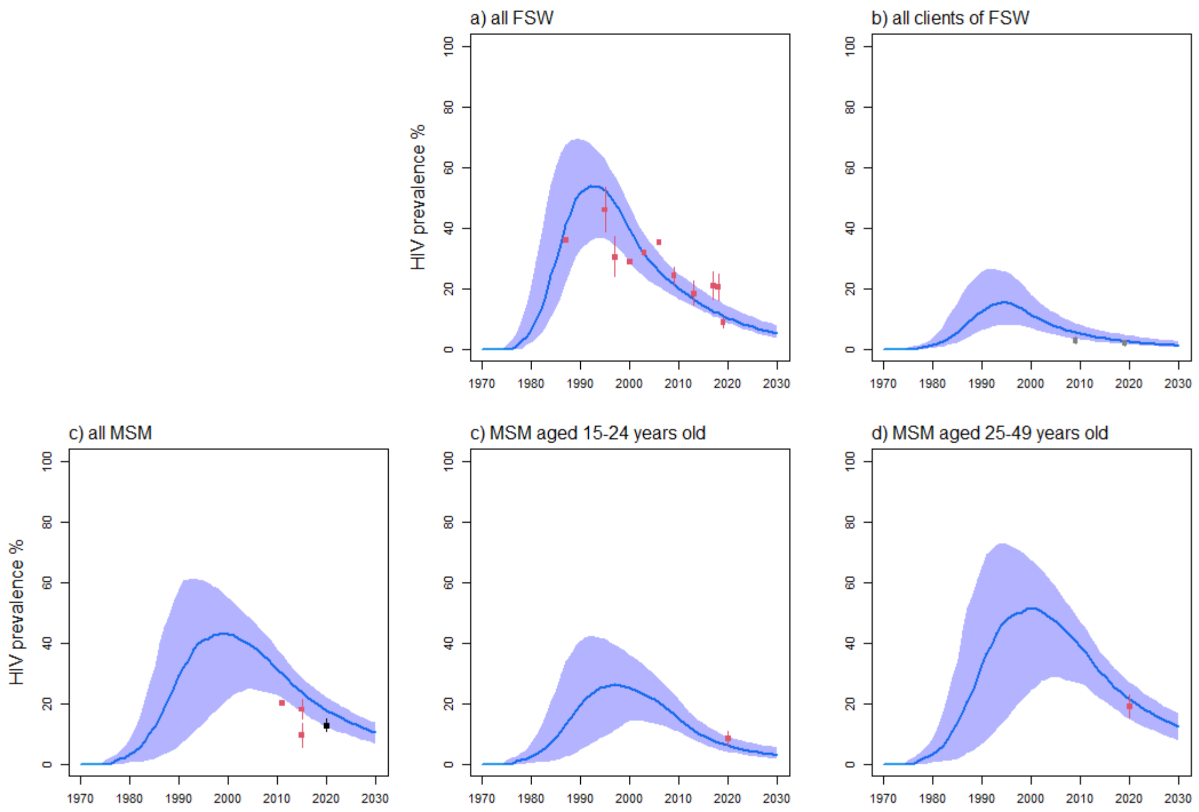

**Figure S5f:** Mali model fitting to HIV prevalence estimates among all a) FSW, b) clients of FSW, c) MSM, as well as c) MSM aged 15-24 years old, and d) aged 25-49 years old. Blue curves and shades represent median and 90% UI (5th and 95th percentiles across model fits), red squares and intervals represent empirical estimates used for model fitting, whereas the dark square on panel c) represents the estimate fitted by age.

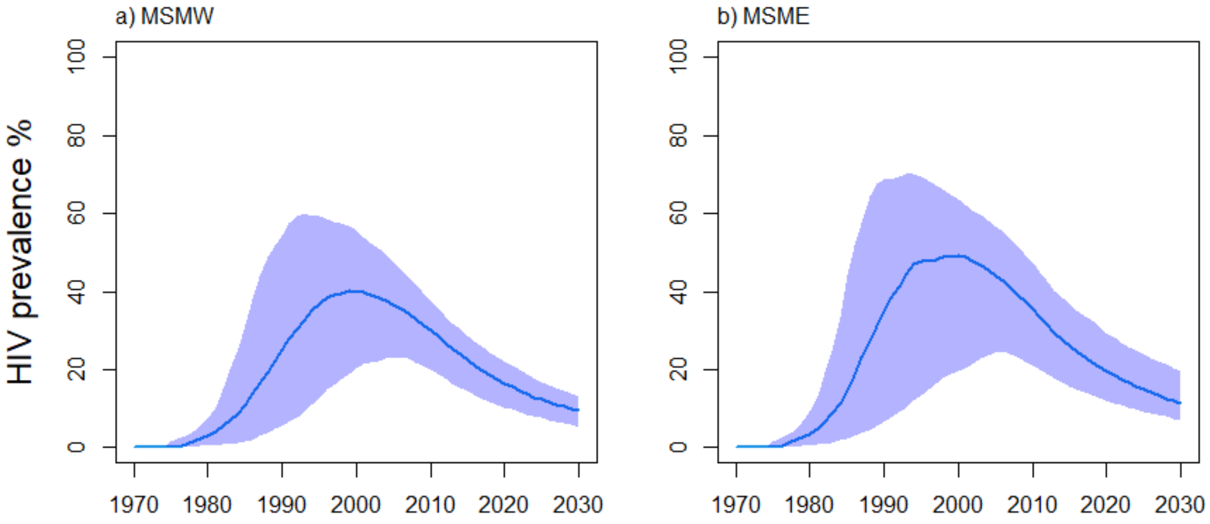

**Figure S5g:** Mali model estimates of the HIV prevalence among all a) MSMW (men having sex with both men and women) and b) MSME (men having sex with another men) MSM. Blue curves and shades represent median and 90% UI (5th and 95th percentiles across model fits). No data was available.

### HIV treatment cascade

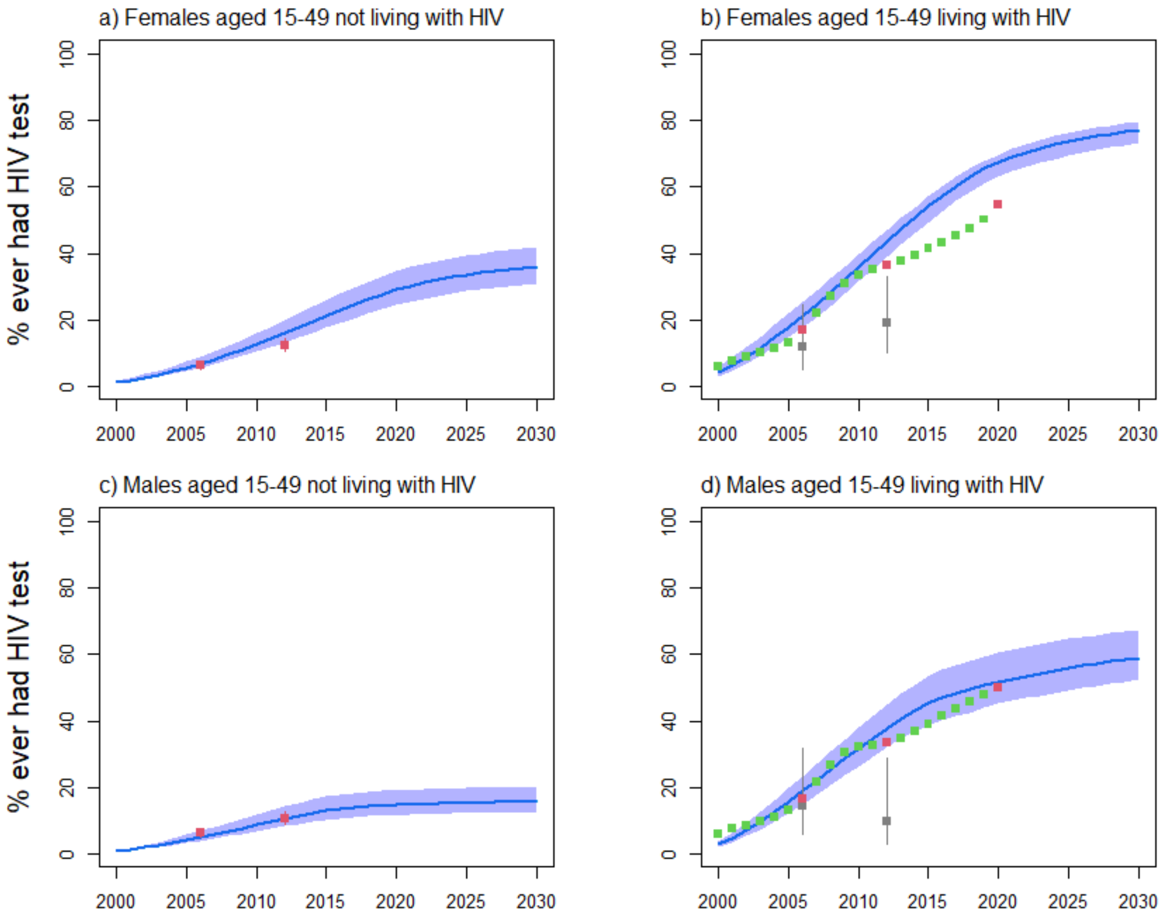

**Figure S5h:** Mali model fitting to the fraction ever having tested for HIV among all females aged 15-49 years old a) not living with HIV, b) living with HIV, and males aged 15-49 years old c) not living with HIV, d) living with HIV. Blue curves and shades represent median and 90% UI ($5^{th}$ and $95^{th}$ percentiles across model fits), red squares and intervals represent estimates from UNAIDS Shiny90 [126] used for model fitting (green=Shiny90 estimates used for comparison), whereas grey squares represent estimates from PLHIV from DHS surveys.

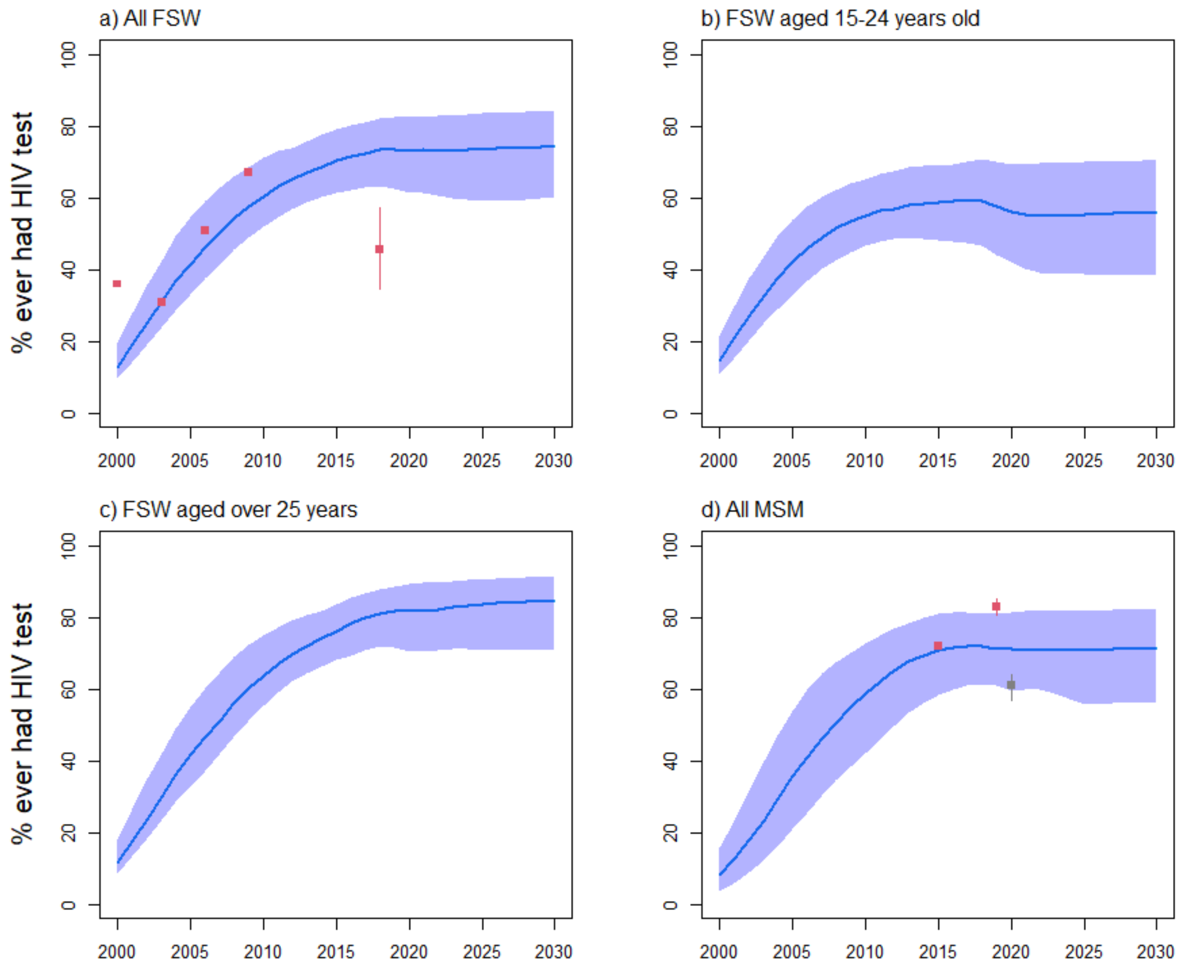

**Figure S5i:** Mali model fitting to the fraction ever having tested for HIV among a) all FSW, b) FSW aged 15-24 years, c) FSW aged 25-49 years, and d) MSM. Blue curves and shades represent median and 90% UI (5th and 95th percentiles across model fits), red squares and intervals represent empirical estimates used for model fitting, the grey square for 2020 in panel d) was an estimate of the fraction of MSM having had an HIV test in the last year, which was used for comparison.

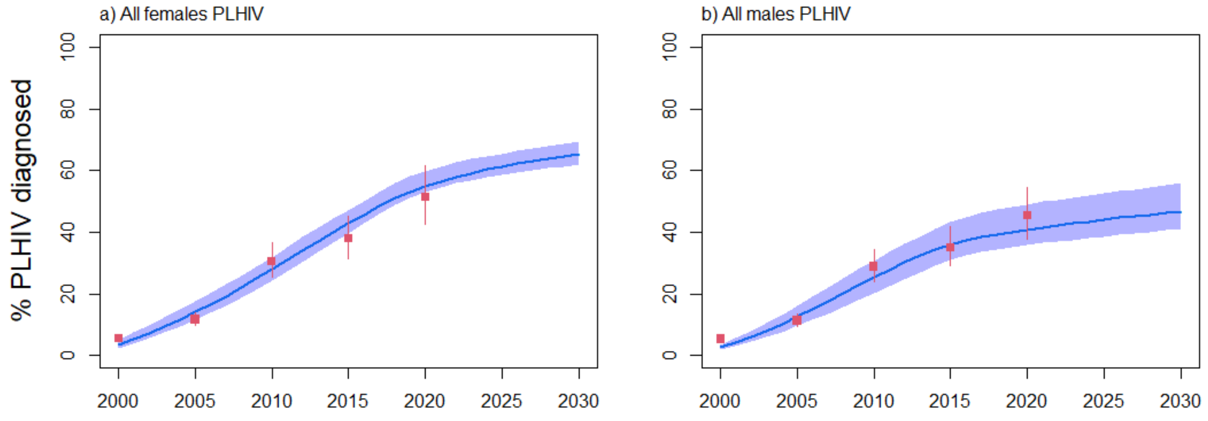

**Figure S5j:** Mali model fitting to the fraction of a) all females living with HIV and b) all males living with HIV being diagnosed. Blue curves and shades represent median and 90% UI (5th and 95th percentiles across model fits), red squares and intervals represent estimates from UNAIDS Shiny90 used for model fitting.

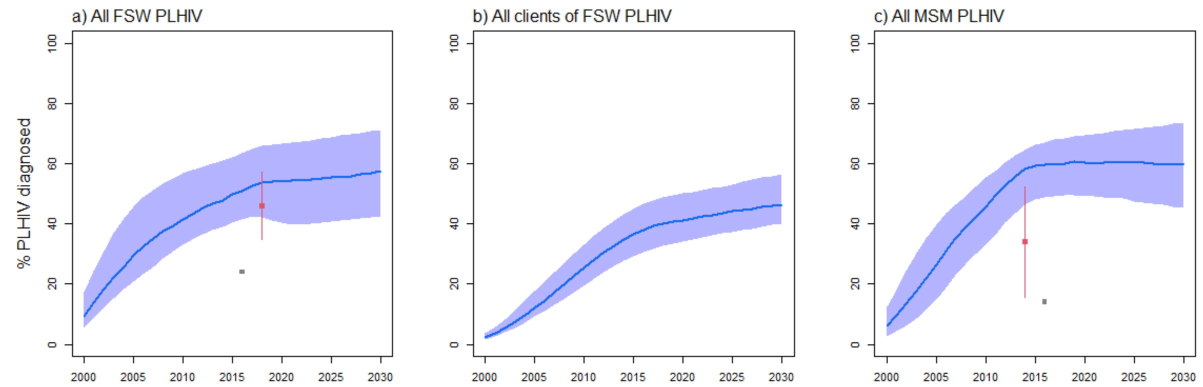

**Figure S5k:** Mali model fitting to the fraction of a) all FSW living with HIV, b) all male clients of FSW living with HIV, and c) all MSM living with HIV diagnosed. Blue curves and shades represent median and 90% UI (5th and 95th percentiles across model fits), red squares and intervals represent estimates from empirical surveys[47,101], whereas grey squares represent national estimates for which no report or underlying study could be identified, and which were only used for comparison.

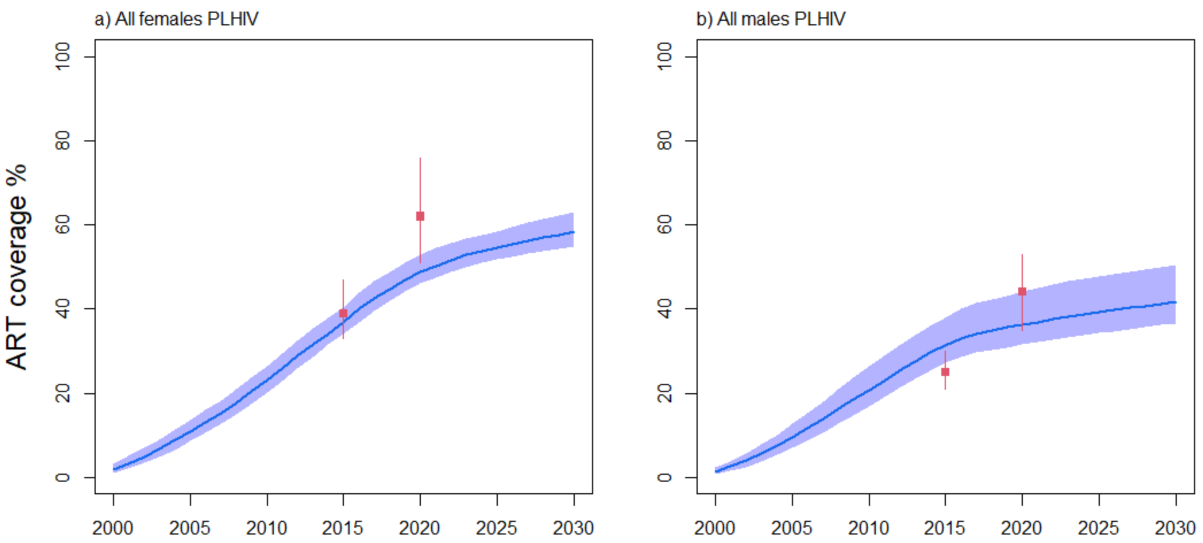

**Figure S5l:** Mali model fitting to ART coverage among a) all females and b) all males living with HIV. Blue curves and shades represent median and 90% UI (5th and 95th percentiles across model fits), whereas red squares and intervals represent estimates from UNAIDS.

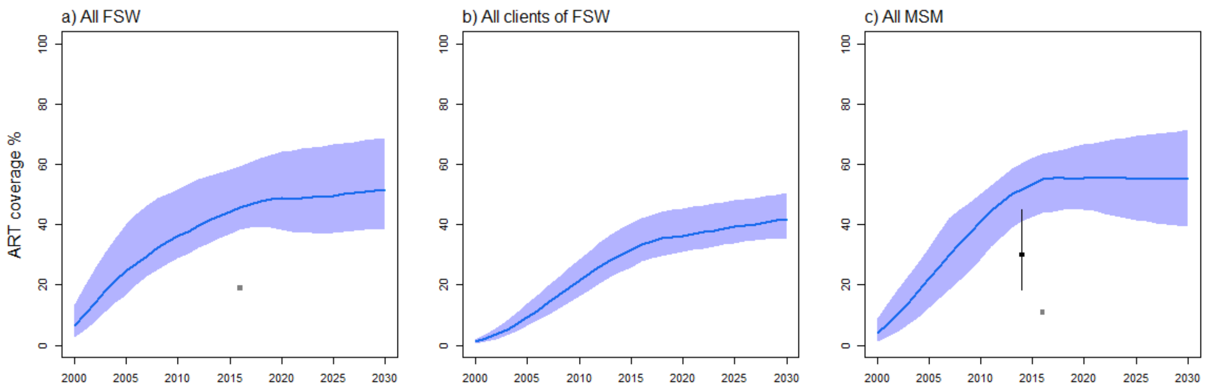

**Figure S5m:** Mali model fitting to ART coverage among a) all FSW, b) all male clients of FSW, and c) all MSM living with HIV. Blue curves and shades represent median and 90% UI (5th and 95th percentiles across model fits), whereas red squares and intervals represent empirical estimates from local surveys. Dark point and interval in panel c) represent self-reported use of ART, and all study participants reporting being on ART were virally suppressed (which was fitted by age in our model). Grey squares in panels a) and c) represent estimates for which no report or underlying study could be identified. Although no ART coverage data was used, we fitted the Mali model to estimates of viral suppression in the country.

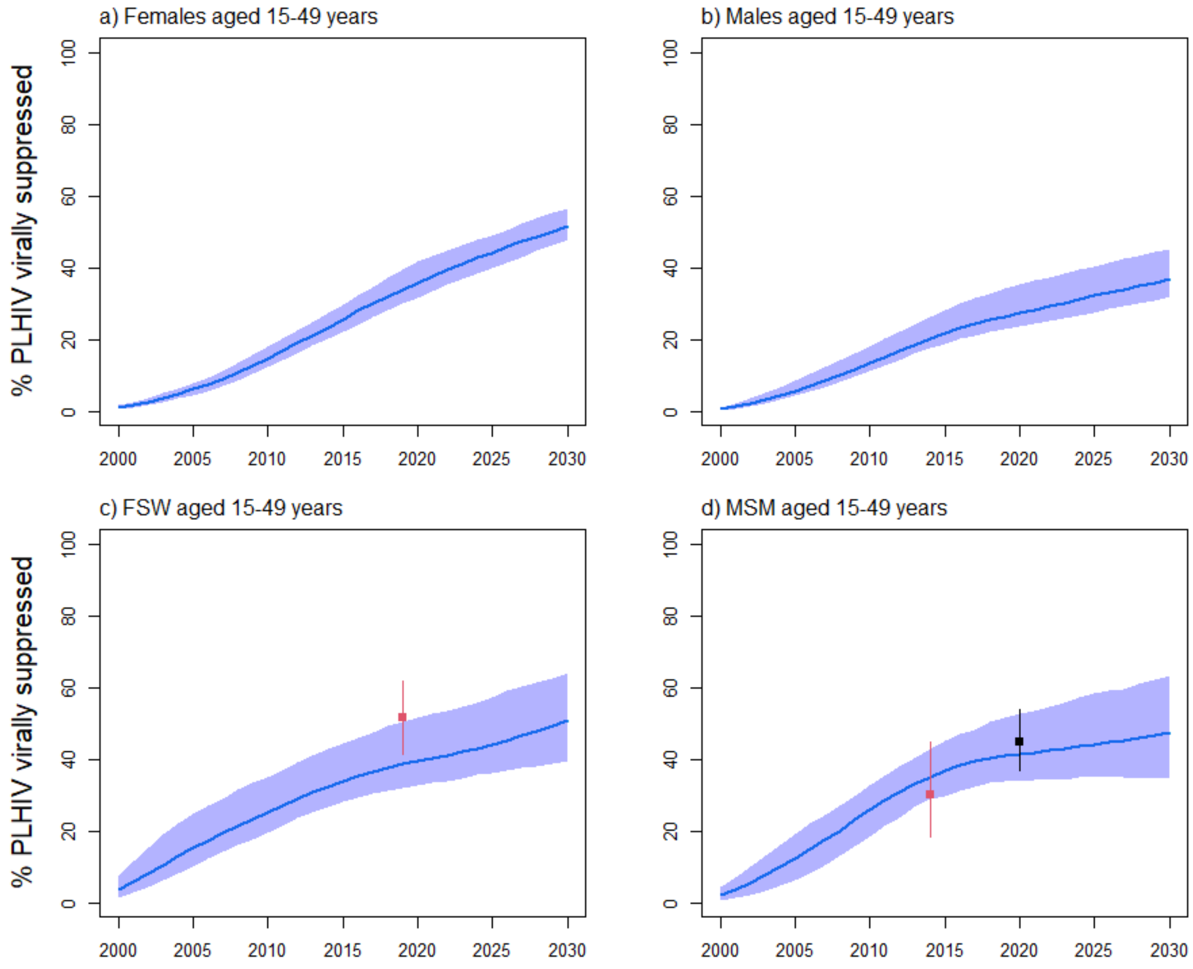

**Figure S5n:** Mali model fitting to HIV viral load suppression coverage among a) all females, b) all males, c) all FSW, and d) all MSM aged 15-49 years living with HIV. Blue curves and shades represent median and 90% UI (5th and 95th percentiles across model fits), whereas red squares and intervals represent empirical estimates from local surveys. The black square in panel d) was only used for comparison; it was aggregated from the age-stratified 2020 estimates used at the fitting stage (see figure S5o).

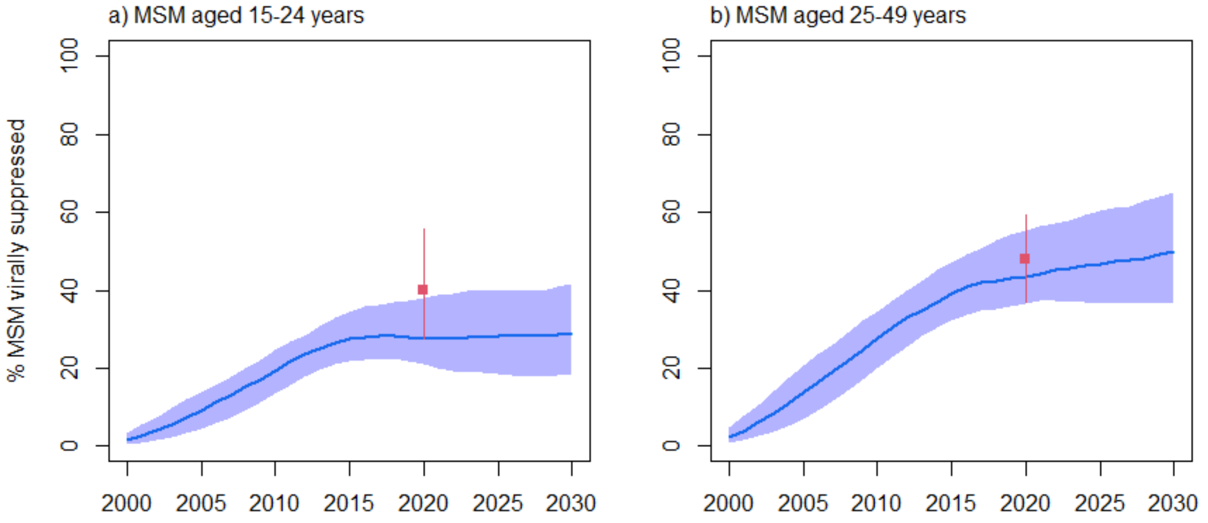

**Figure S5o:** Mali model fitting to HIV viral load suppression coverage among MSM living with HIV aged a) 15-24 years and b) 25-49 years. Blue curves and shades represent median and 90% UI (5th and 95th percentiles across model fits), whereas red squares and intervals represent empirical estimates.

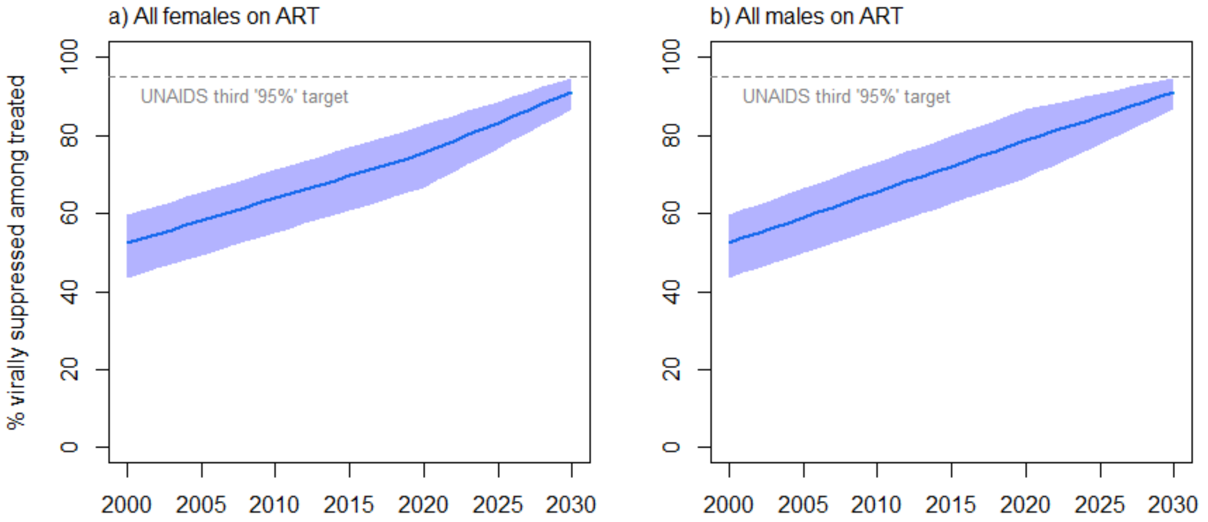

**Figure S5p:** Mali model estimates of the fraction of a) females and b) males living with HIV on ART which are virally suppressed (the third UNAIDSs "95%" indicator) over time, which is used as a parameter in our model. There were no available estimates of this fraction, and the plausible fractions were estimated by using the relationship between the 1st and 3rd "95%" indicators in Côte d'Ivoire and Senegal and applying this relationship to the estimate of the 1st "95%" indicator in Mali. The grey dashed line corresponds to the UNAIDS's third "95%" target whereby 95% of PLHIV on ART should be virally suppressed in 2025.

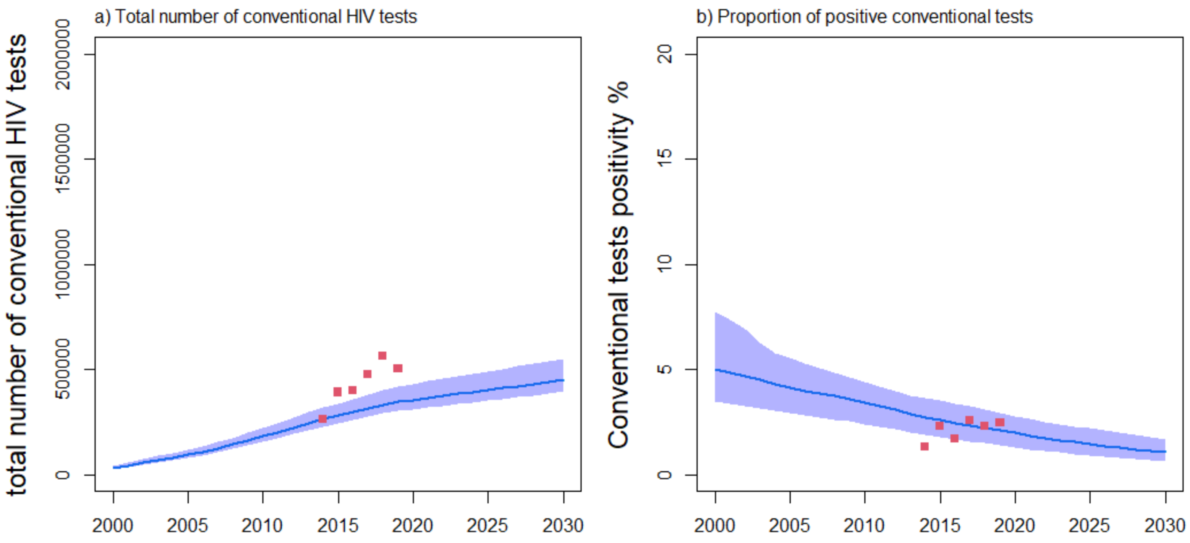

**Figure S5q:** Mali model fitting to programmatic data[67] on a) the total number of conventional tests and b) proportion of these tests which were positive. Blue curves and shades represent median and 90%UI (5th and 95th percentiles across model fits), whereas red squares represent programmatic data from UNAIDS[67].

### Results: model fits (Senegal)

#### Demography

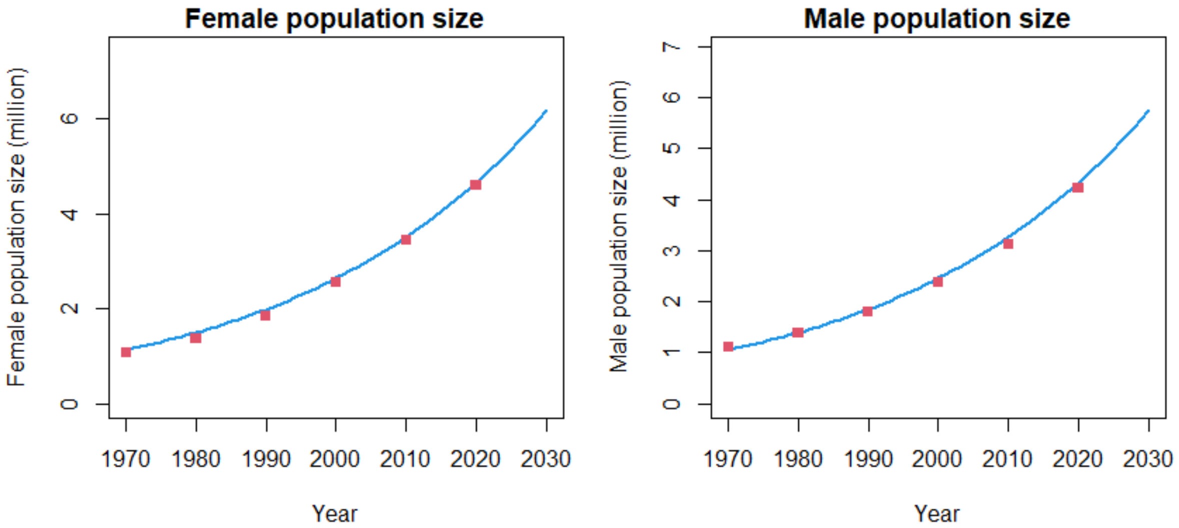

**Figure S6a:** Senegal model fitting to the size of the (left) female and (right) male populations aged 15-59 years old over time. Blue curves represent model estimates, while red squares show estimates from UNPD.

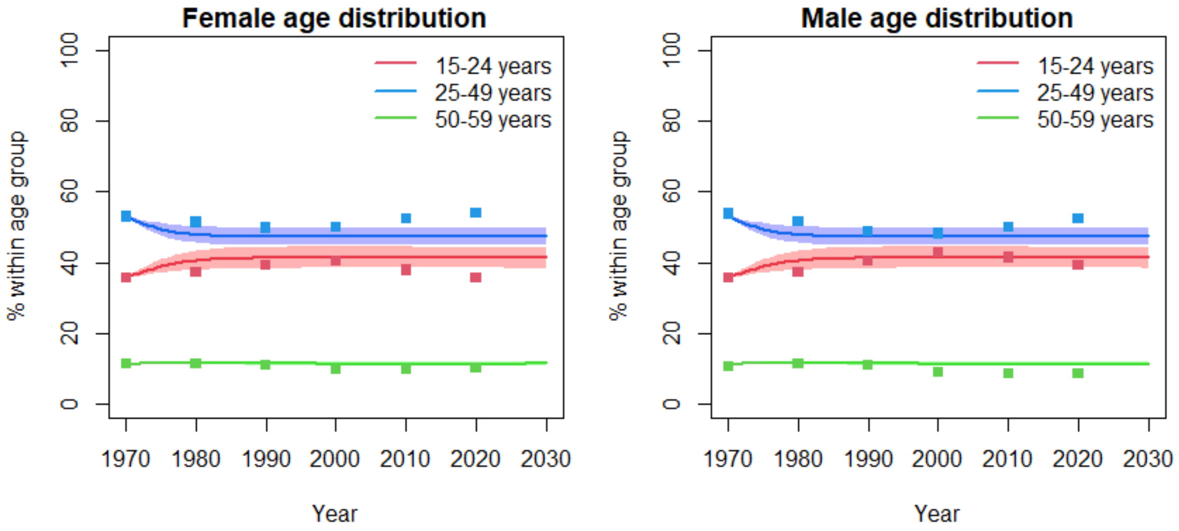

**Figure S6b:** Senegal model fitting to the age distribution of (left) females and (right) male populations aged 15-59 years old over time. Curves represent model estimates while squares show estimates from UNPD.

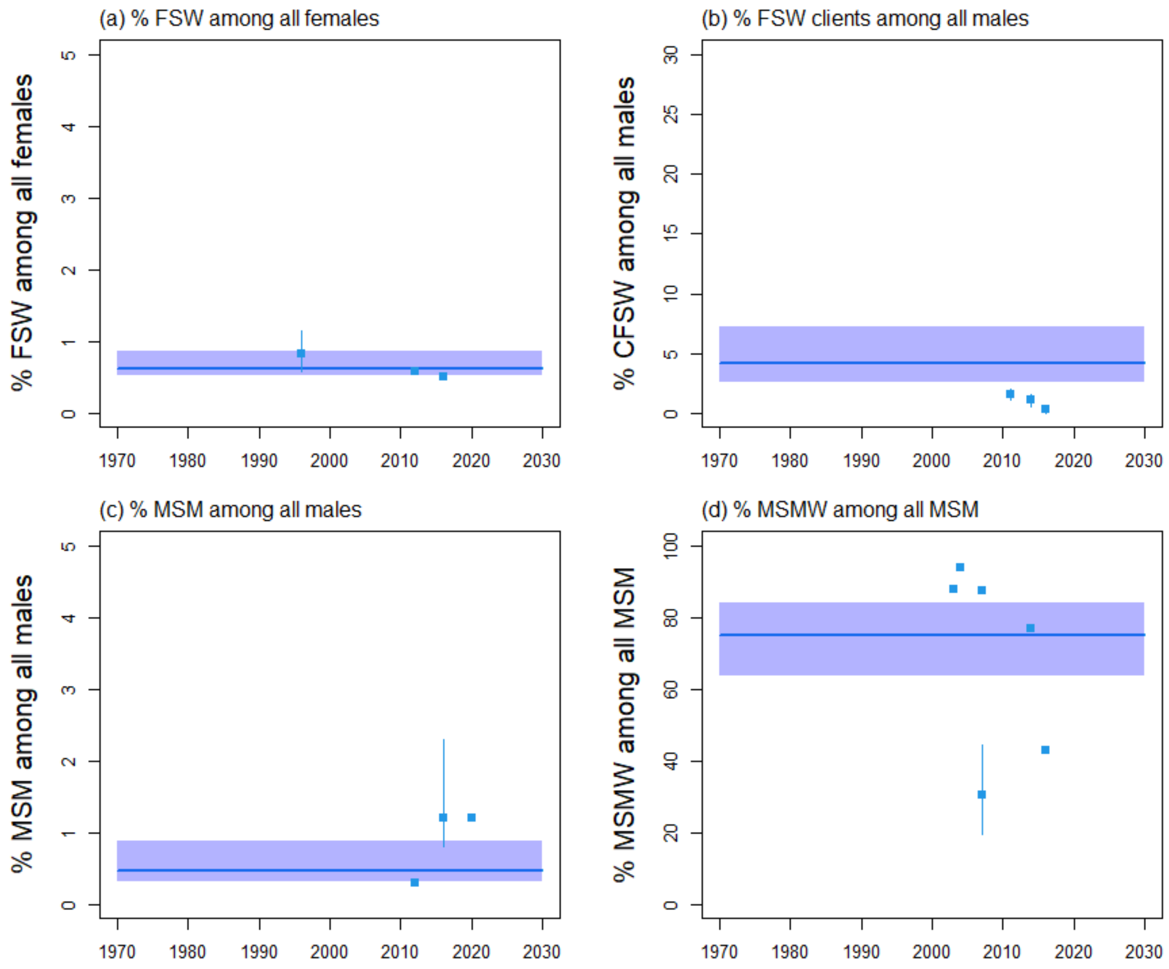

**Figure S6c:** Senegal model fitting to the size of key populations and their clients, with the fraction of a) FSW among all females aged 15-59 years, b) FSW clients among all males aged 15-59 years, c) MSM among all males aged 15-59 years, and d) MSMW (men who have sex with men and women) among all MSM aged 15-59 years old over time. Blue curves and shades represent median and 90% UI (5th$^{th}$ and 95$^{th}$ percentiles across model fits), whereas squares and intervals represent empirical estimates.

#### HIV epidemiology

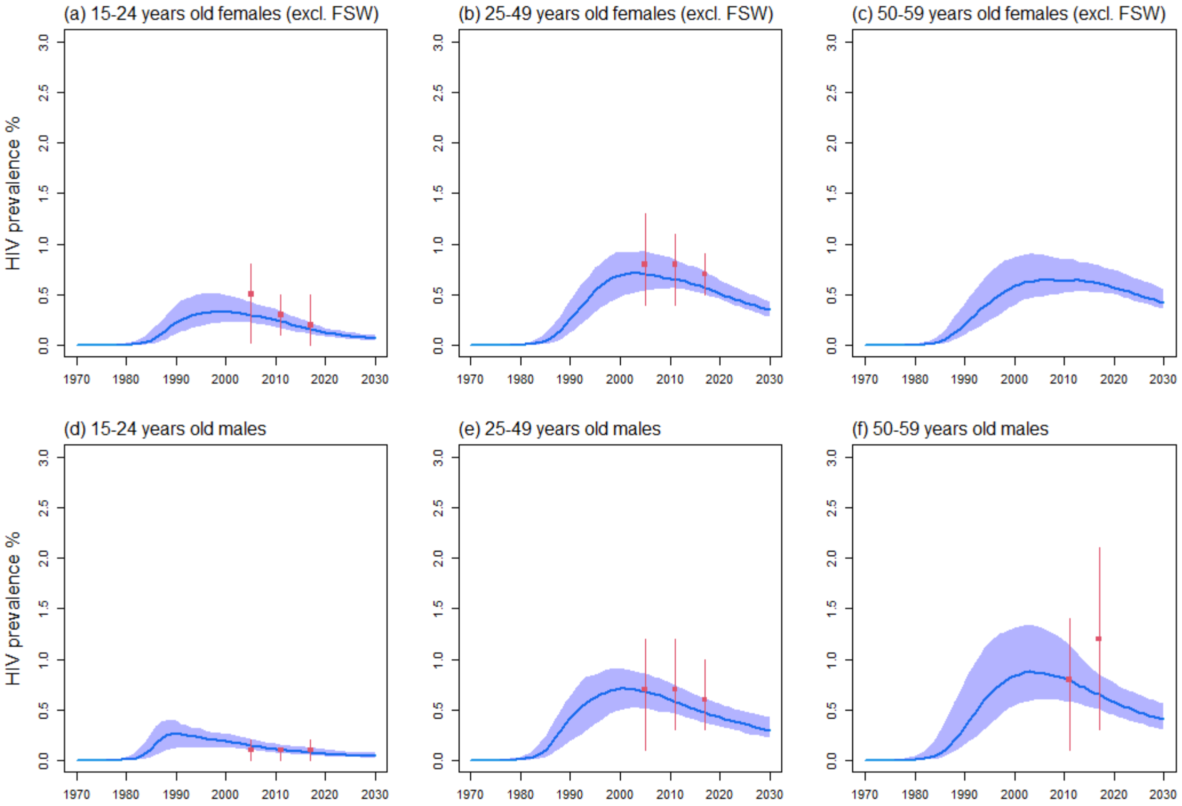

**Figure S6d:** Senegal model fitting to the HIV prevalence among all females aged a) 15-24, b) 25-49, and c) 50-59 years old (excluding FSW), as well as all males aged d) 15-24, e) 25-49 years, and f) 50-59 years old. Blue curves and shades represent median and 90% UI (5$^{th}$ and 95$^{th}$ percentiles across model fits), whereas red squares and intervals represent empirical estimates used for model fitting.

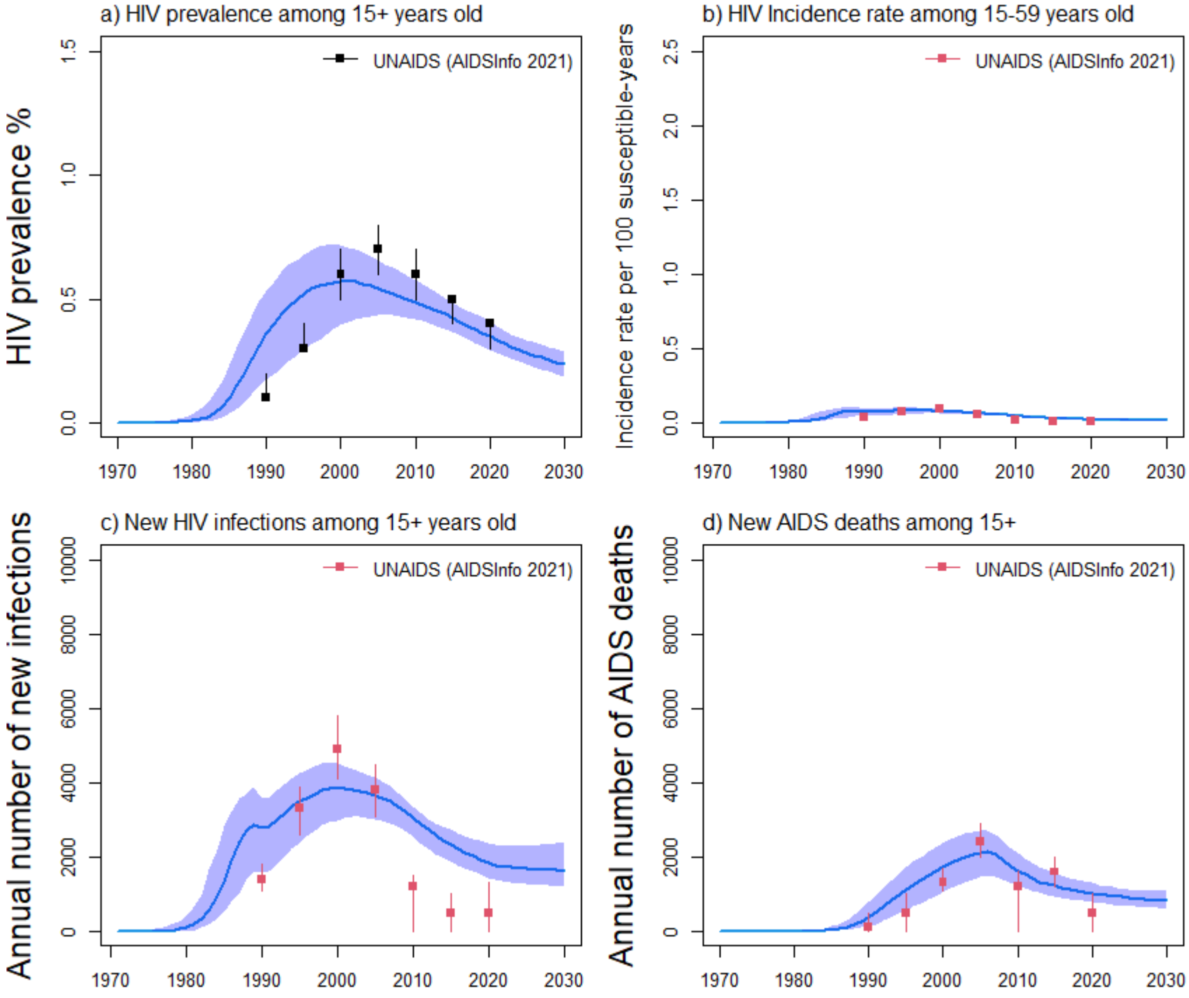

**Figure S6e:** Estimates of HIV prevalence among a) all adults aged over 15 years old, fits to b) HIV incidence rate, c) annual number of new HIV infections and d) annual HIV-related deaths in Senegal from UNAIDS. Blue curves and shades represent median and 90% UI ($5^{th}$ and $95^{th}$ percentiles across model fits), red squares and intervals represent empirical estimates used for model fitting, whereas the dark squares and intervals in panel a) represent estimates from UNAIDS only used for comparison.

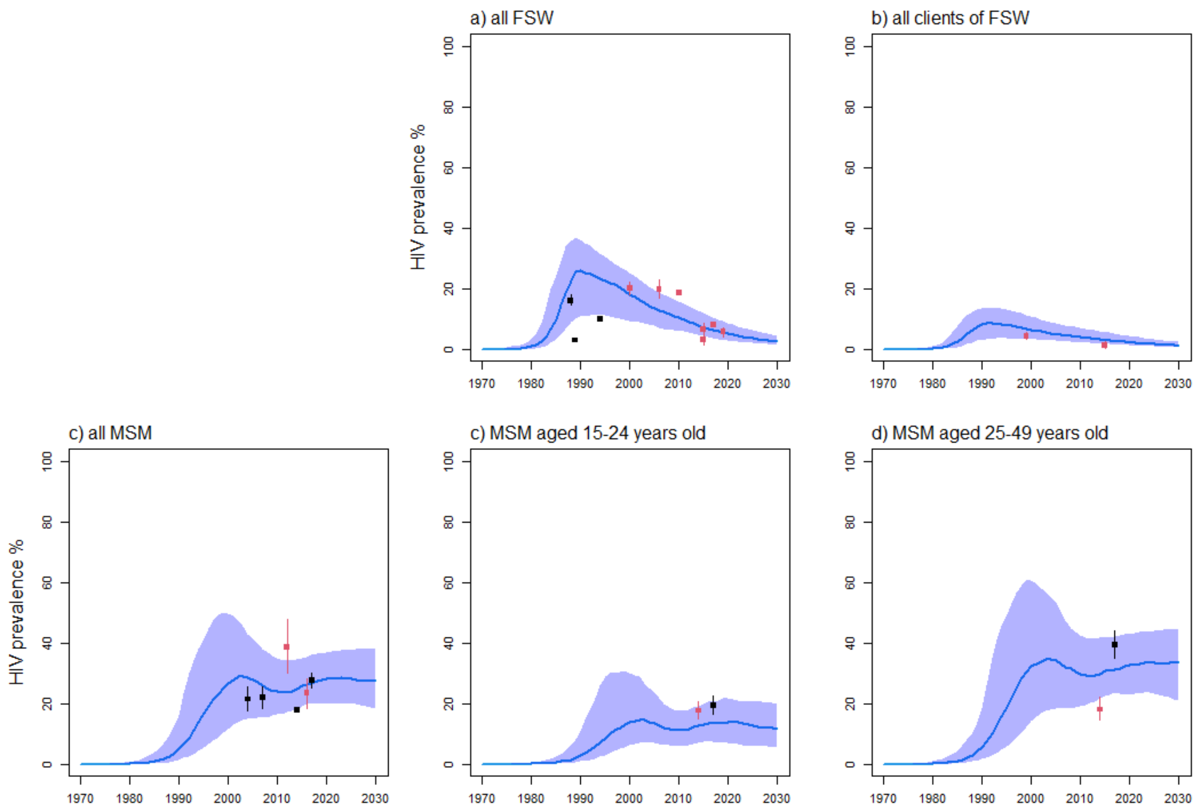

**Figure S6f:** Senegal model fitting to HIV prevalence estimates among a) all FSW, b) all clients of FSW, c) MSM, as well as c) MSM aged 15-24 years old, and d) aged 25-49 years old. Blue curves and shades represent median and 90% UI (5$^{th}$ and 95$^{th}$ percentiles across model fits), red squares and intervals represent empirical estimates used for model fitting. Dark squares for FSW correspond to estimates which could not be sourced to a particular study or report and were only used for comparison, whereas those on panel c-d) represent estimates fitted by age and for bisexuals/exclusive MSM separately.

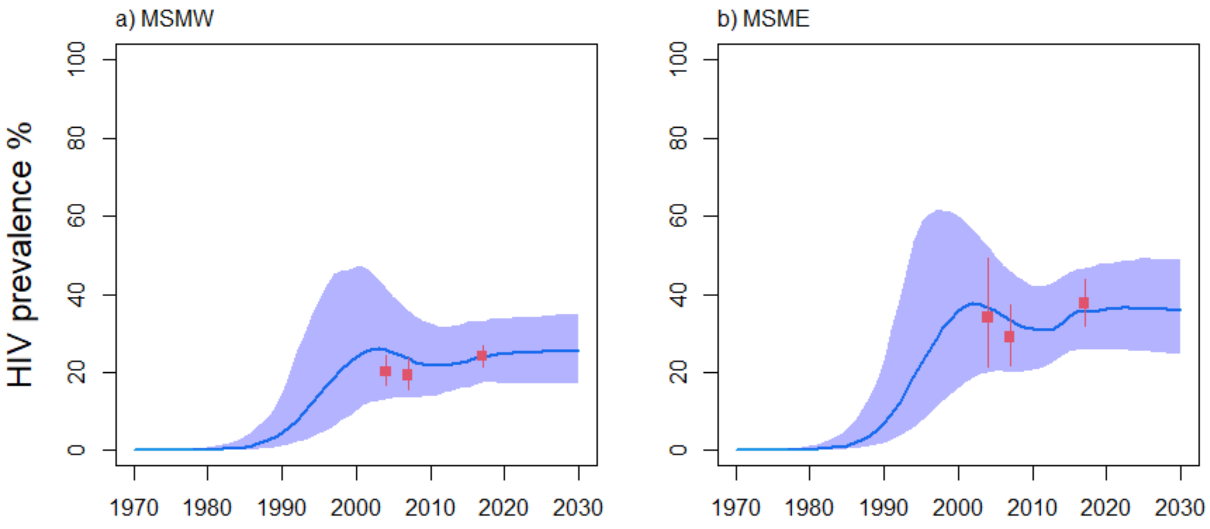

**Figure S6g:** Senegal model fitting to HIV prevalence estimates among all a) MSMW (men having sex with both men and women) and b) MSME (men having sex with other men exclusively) MSM. Blue curves and shades represent median and 90% UI (5th and 95th percentiles across model fits), whereas red squares and intervals represent empirical estimates.

#### HIV treatment cascade

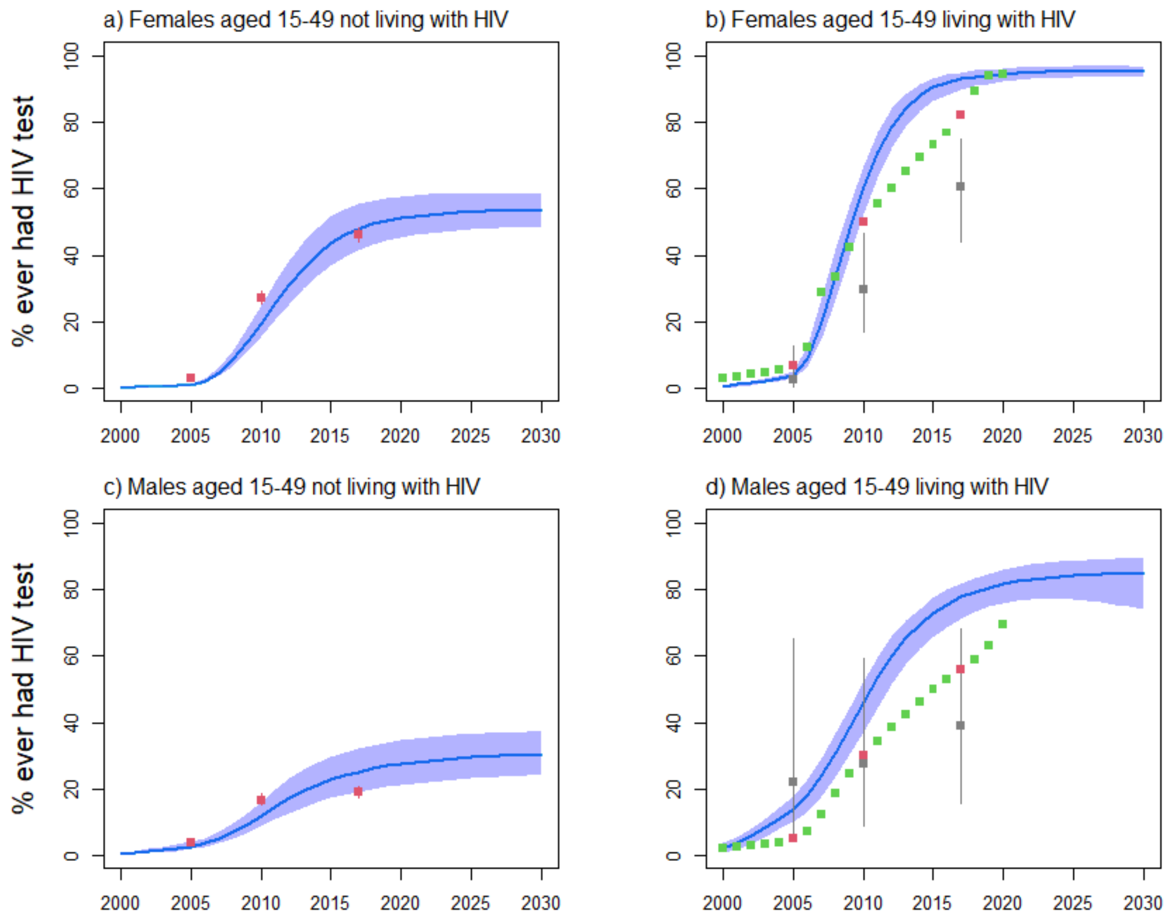

**Figure S6h:** Senegal model fitting to the fraction ever having tested for HIV among all females aged 15-49 years old a) not living with HIV, and b) living with HIV, and males aged 15-49 years old c) not living with HIV, and d) living with HIV. Blue curves and shades represent median and 90% UI (5th and 95th percentiles across model fits), red squares and intervals represent estimates from UNAIDS Shiny90 used for model fitting (green=those used for comparison), whereas grey squares represent estimates among PLHIV from DHS surveys. Our model estimates among PLHIV were higher than empirical estimates because of high coverage of ART in the data.

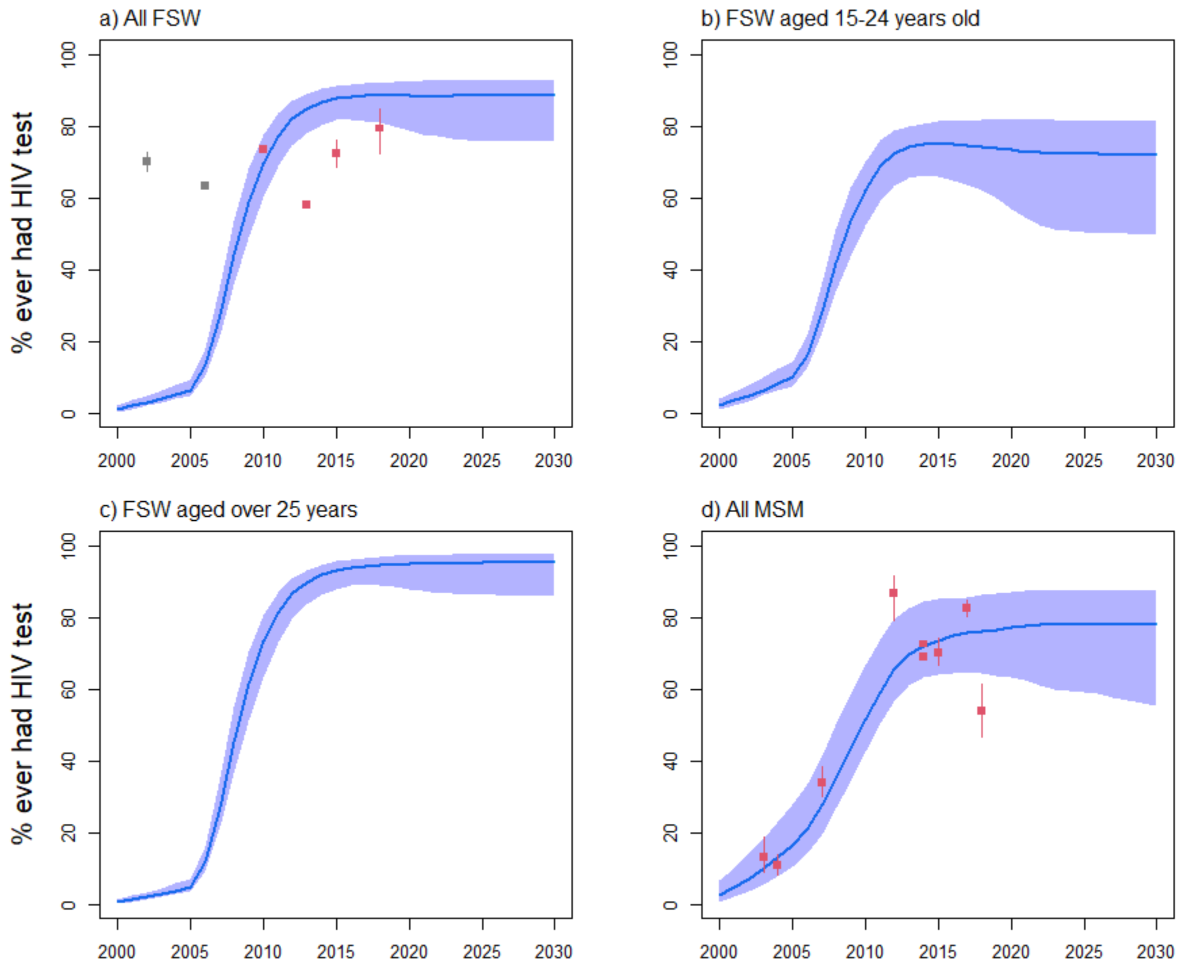

**Figure S6i:** Senegal model fitting to the fraction ever having tested for HIV among a) all FSW, b) FSW aged 15-24 years, c) FSW aged 25-49 years, and d) all MSM. Blue curves and shades represent median and 90% UI (5$^{th}$ and 95$^{th}$ percentiles across model fits), red squares and intervals represent empirical estimates used for model fitting, whereas the grey squares in panel a) corresponded to studies from STI clinics, which were only used for comparison.

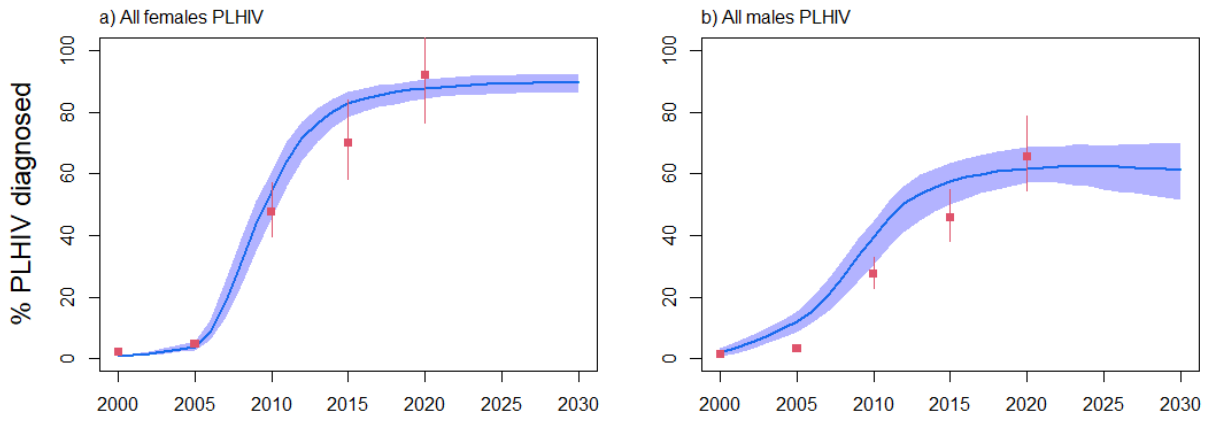

**Figure S6j:** Senegal model fitting to the fraction of a) all females living with HIV and b) all males living with HIV being diagnosed. Blue curves and shades represent median and 90% UI (5th and 95th percentiles across model fits), red squares and intervals represent estimates from UNAIDS Shiny90 used for model fitting.

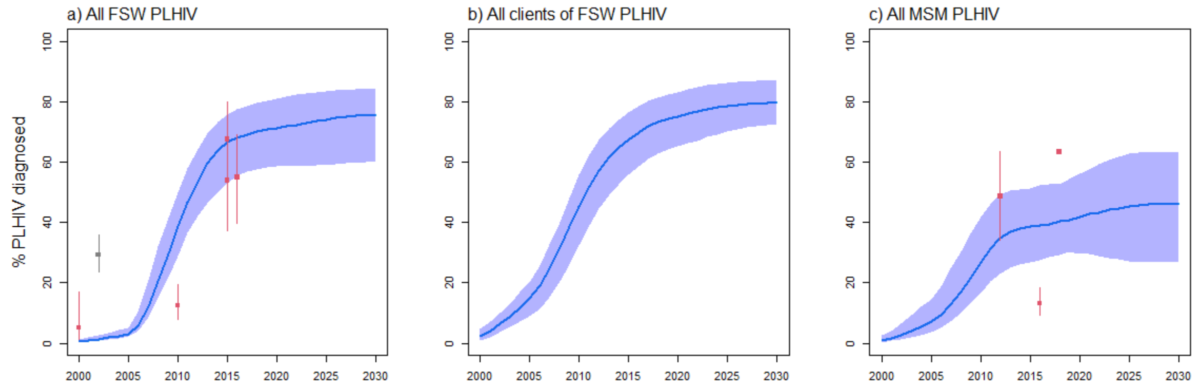

**Figure S6k:** Senegal model fitting to the fraction of a) all FSW living with HIV, b) all male clients of FSW living, and c) all MSM living with HIV being diagnosed. Blue curves and shades represent median and 90% UI (5th and 95th percentiles across model fits), red squares and intervals represent estimates from empirical surveys, whereas grey square in panel a) represent estimates from an STI clinic.

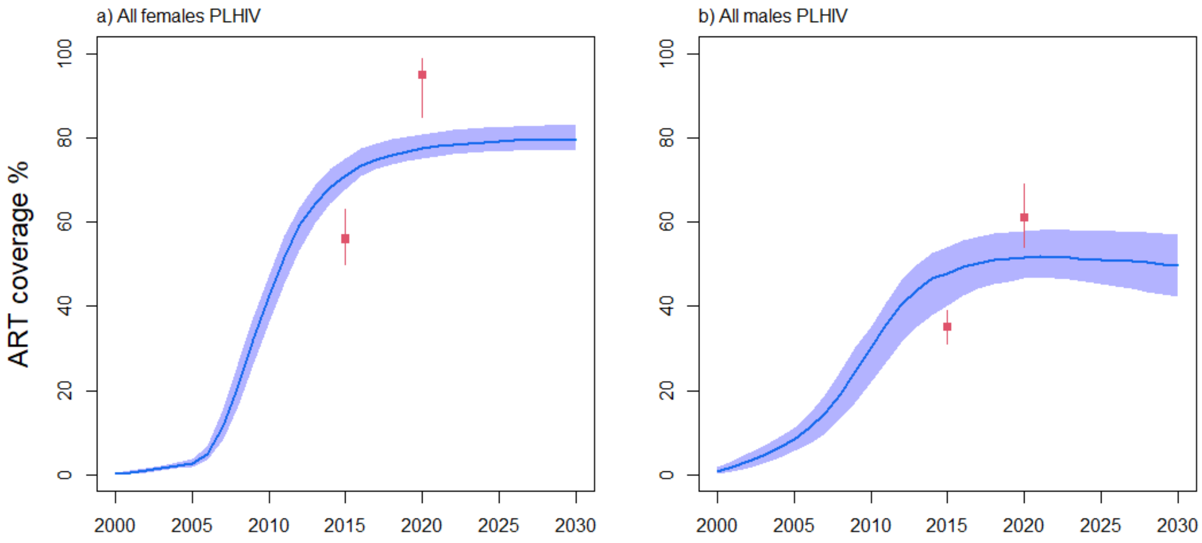

**Figure S6l:** Senegal model fitting to ART coverage among a) all females and b) all males. Blue curves and shades represent median and 90% UI (5th and 95th percentiles across model fits), whereas red squares and intervals represent estimates from UNAIDS.

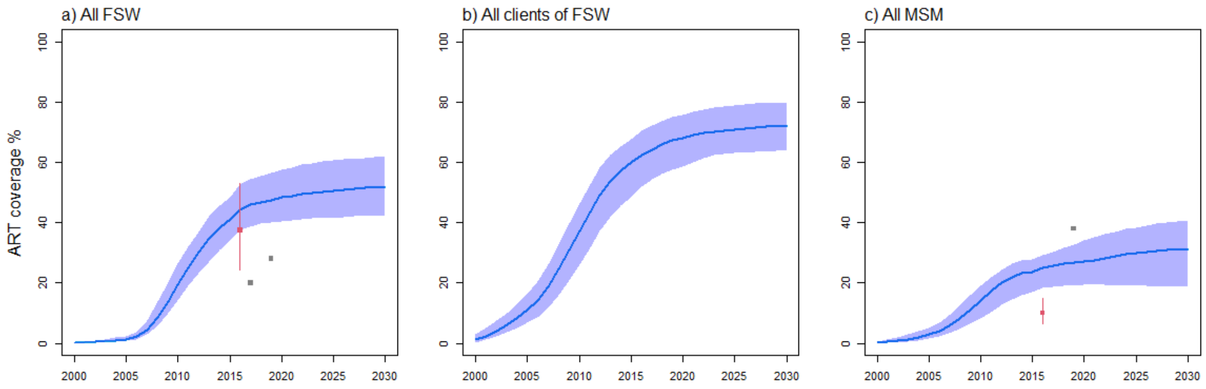

**Figure S6m:** Senegal model fitting to ART coverage among a) all FSW, b) all male clients of FSW, and c) all MSM. Blue curves and shades represent median and 90% UI (5th and 95th percentiles across model fits), whereas red squares and intervals represent empirical estimates from local surveys. Dark point and interval in panel c) represent self-reported use of ART (as opposed to estimates using viral load data), and all study participants reporting being on ART were virally suppressed (which was fitted by age in our model). Grey squares in panels a) and c) represent estimates from the UNAIDS key population atlas for which no report or underlying study could be identified.

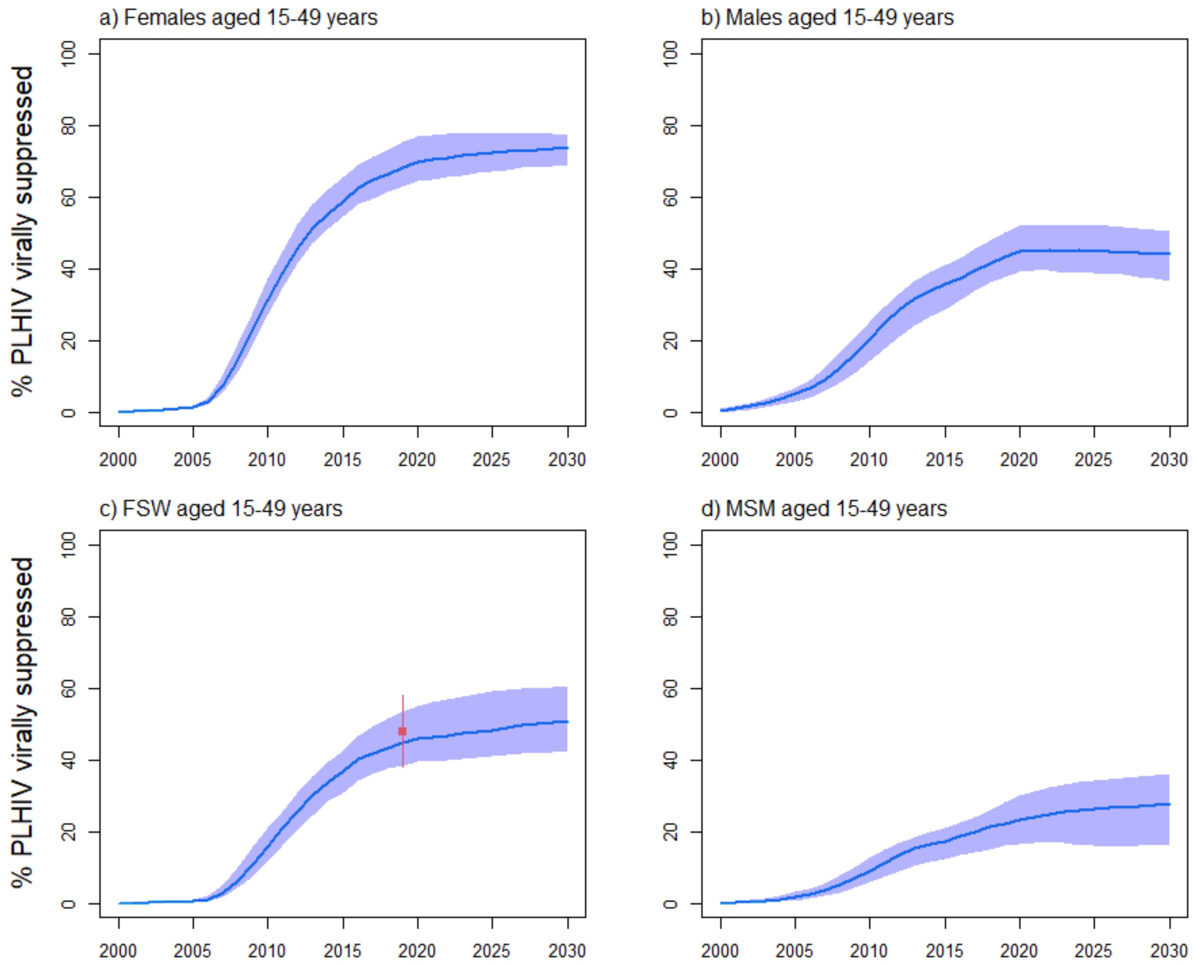

**Figure S6n:** Senegal model fitting to HIV viral load suppression coverage among a) all females, b) all males, c) all FSW, and d) all MSM aged 15-49 years living with HIV. Blue curves and shades represent median and 90% UI (5th and 95th percentiles across model fits), whereas red squares and intervals in panel c) represent empirical estimate from a local survey.

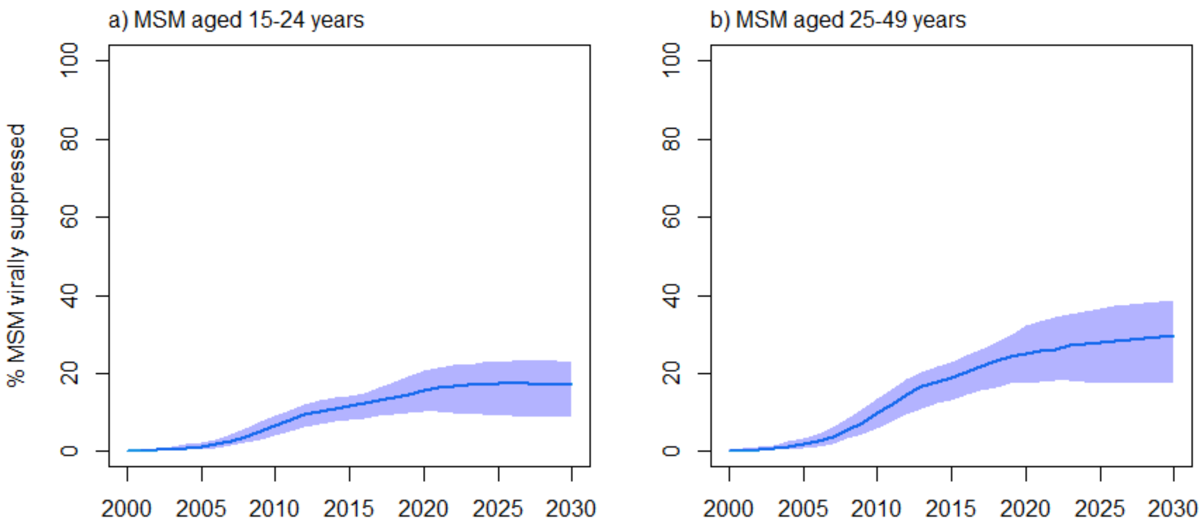

**Figure S6o:** Senegal model fitting to HIV viral load suppression coverage among all MSM aged a) 15-24 years and b) 25-49 years. Blue curves and shades represent median and 90% UI (5th and 95th percentiles across model fits. No data was available.

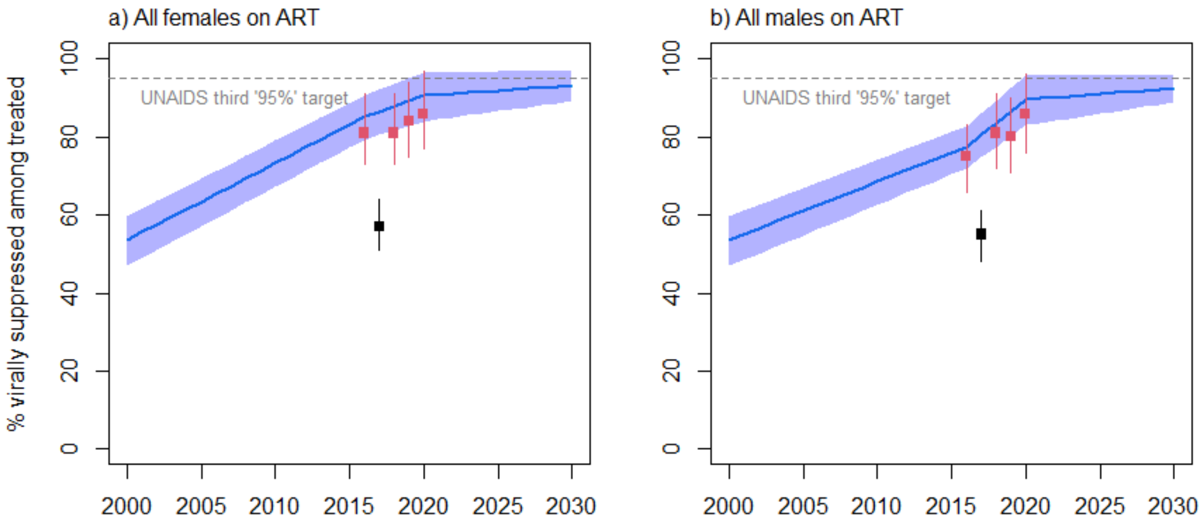

**Figure S6p:** Senegal model fitting to fractions of a) females and b) males living with HIV on ART which are virally suppressed (the third UNAIDSs "95%" indicator) over time. Blue curves and shades represent median and 95% UI (5th and 95th percentiles across model fits), whereas red squares and intervals represent estimates from UNAIDS used as parameters. The grey dashed line corresponds to the UNAIDS's third "95%" target whereby 95% of PLHIV on ART should be virally suppressed in 2025. The estimate for 2017 was assumed to be an outlier and not considered in our analysis.

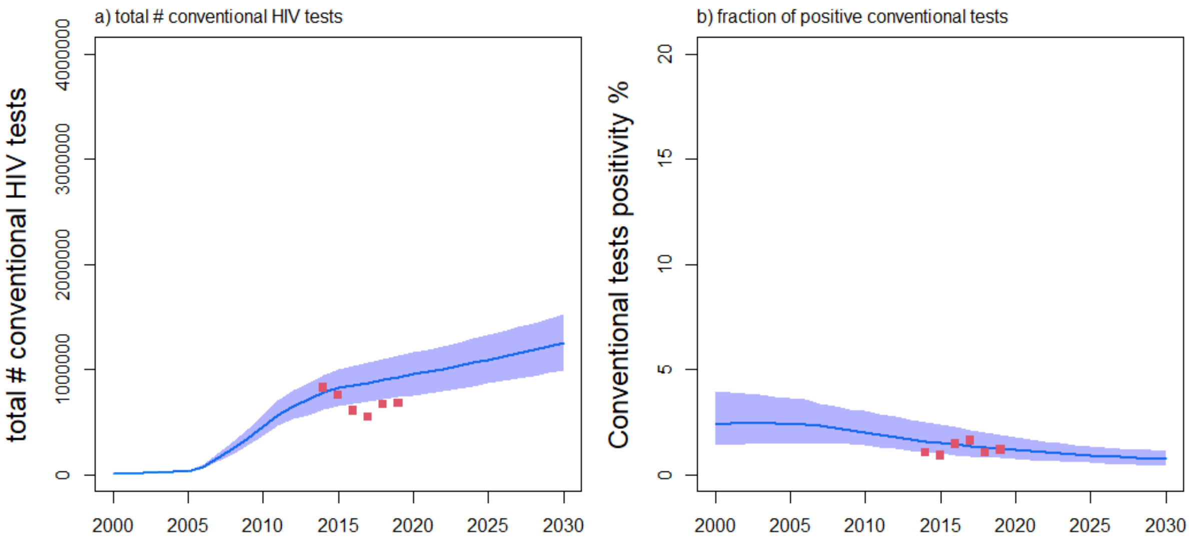

**Figure S6q:** Senegal model fitting to programmatic data on a) the total number of conventional tests among and b) proportion of these tests which were positive. Blue curves and shades represent median and 90% UI (5$^{th}$ and 95$^{th}$ percentiles across model fits), whereas red squares represent programmatic data communicated by UNAIDS.

### Results: HIVST distribution and impact

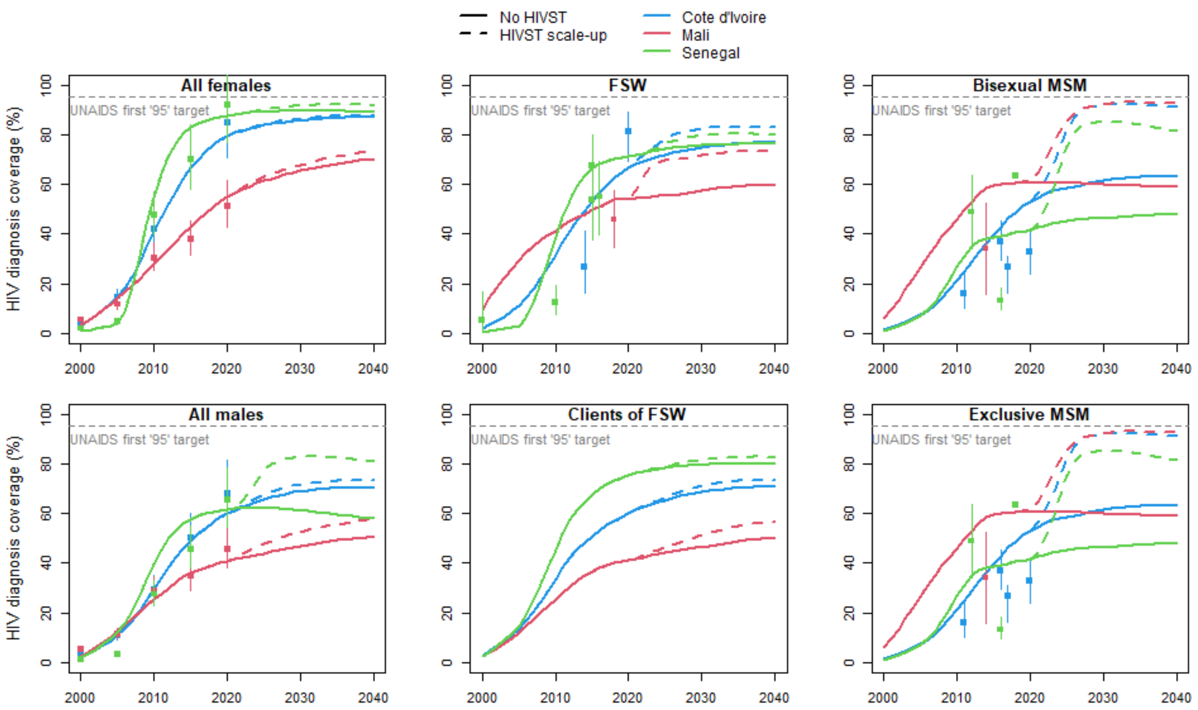

**Figure S7:** Empirical (dots and intervals) and modelled (lines) proportion of diagnosed PLHIV from different risk groups over 2000-2040 in Côte d'Ivoire (blue), Mali (red) and Senegal (green). Plain lines represent median estimates across model fits of a counterfactual scenario ("no HIVST"). Dashed lines represent median estimates under our scenario 3 representing ATLAS distribution of HIVST kits to FSW and MSM over 2019-2021, followed by a scale-up and plateau in HIVST distribution from 2025.

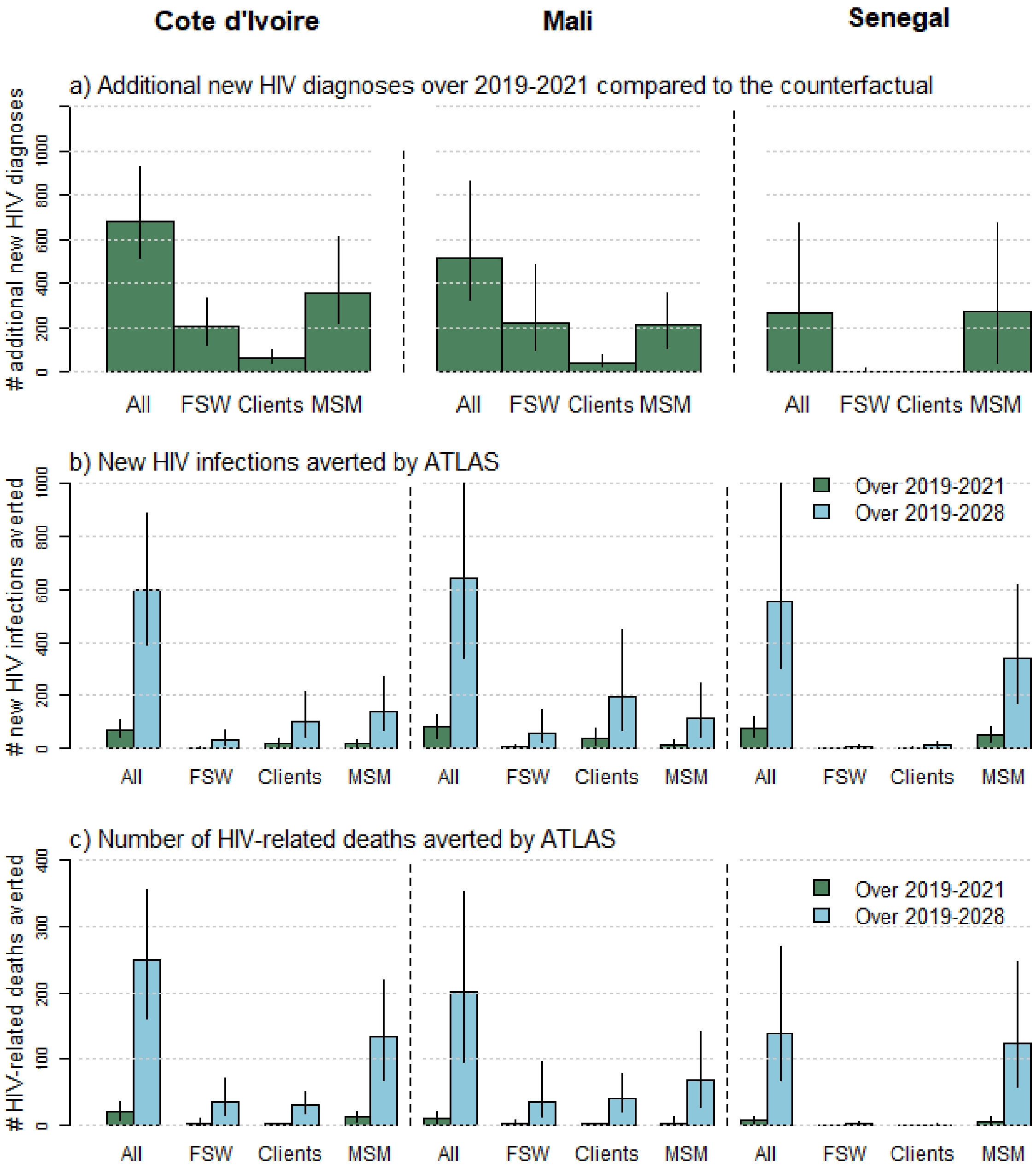

**Figure S8:** Estimated impact (in absolute numbers) of the ATLAS-only scenario compared to the counterfactual ("no-HIVST" scenario): a) additional new HIV diagnoses over the three-year intervention (2019-2021) and numbers of b) HIV infections (c) and HIV-related deaths averted by ATLAS over 2019-2021 (green bars) and 2019-2028 (light blue bars). Bars height represent median of model estimates, whereas error bars represent 90%UI of estimates (5th and 95th percentiles of estimates across model simulations).

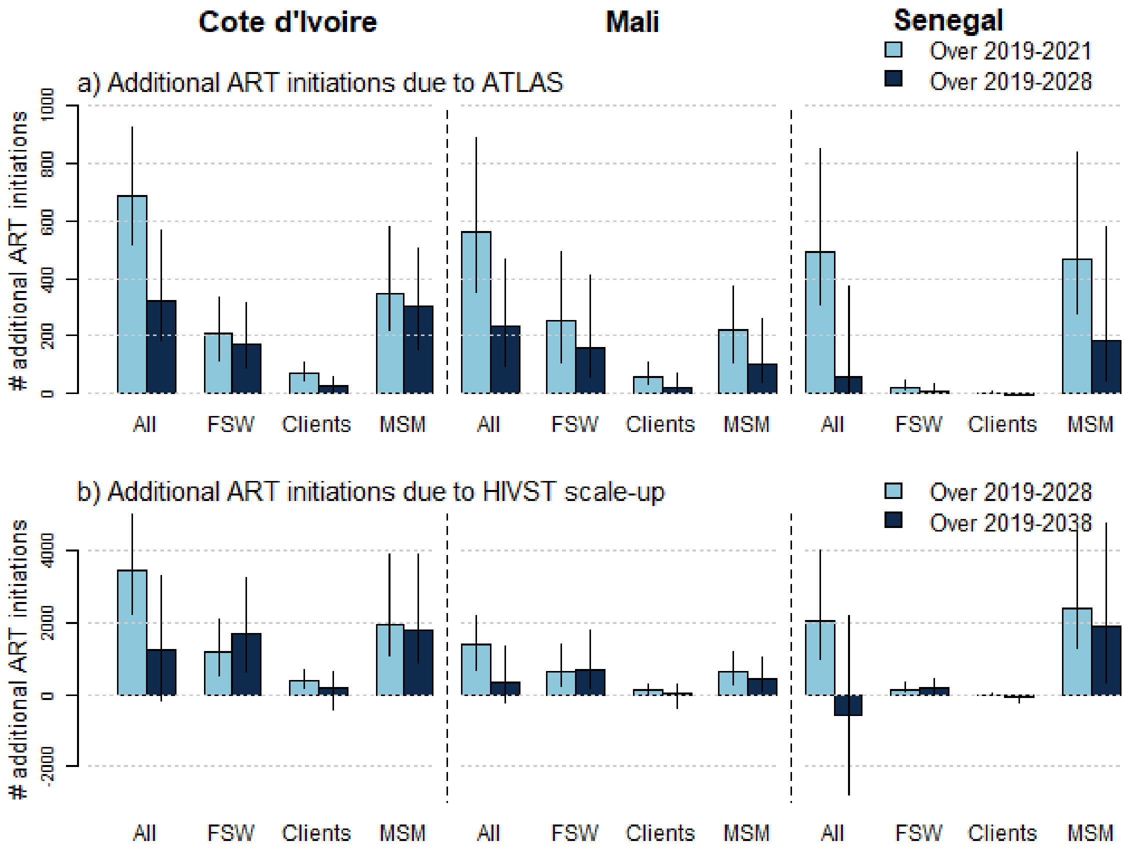

**Figure S9:** Estimated impact (in absolute numbers) of a) ATLAS and b) HIVST scale-up scenario compared to the counterfactual (no-HIVST scenario): additional new ART initiations over a) 2019-2021 (light blue bars) and 2019-2028 (dark blue bars), and b) over 2019-2028 (light blue bars) and 2019-2038 (dark blue bars). Bars height represent median of model, whereas error bars represent 90%UI of estimates ($5^{th}$ and $95^{th}$ percentiles of estimates across model simulations).

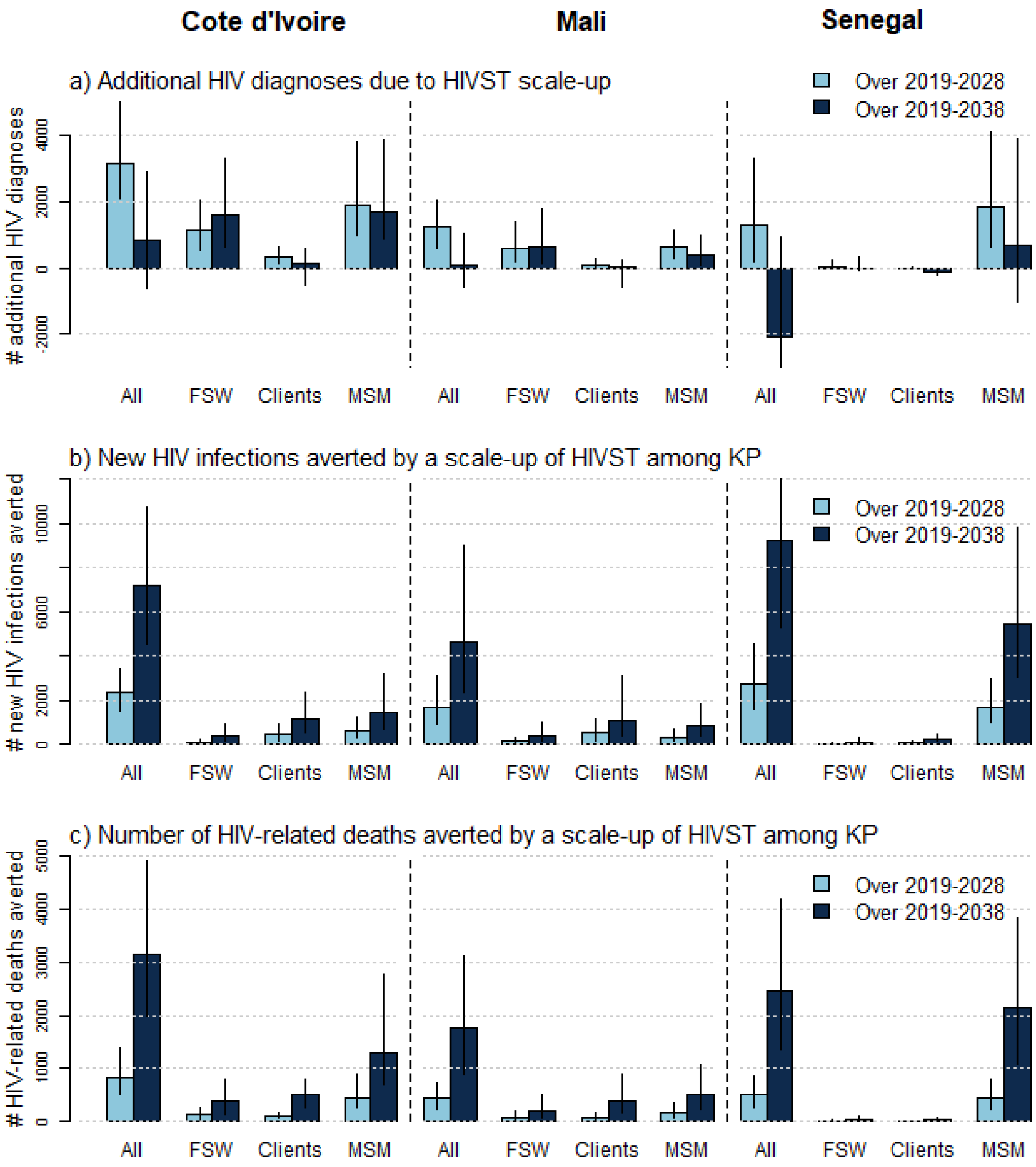

**Figure S10:** Estimated impact (in absolute numbers) of the HIVST scale-up scenario compared to the counterfactual (no-HIVST scenario): a) additional new HIV diagnoses over 2019-2021 (light blue bars) and 2019-2028 (dark blue bars) and numbers of b) HIV infections (c) and HIV-related deaths averted by ATLAS over 2019-2028 and 2019-2038. Bars height represent median of model estimates, whereas error bars represent 90% UI of estimates (5$^{th}$ and 95$^{th}$ percentiles of estimates across model simulations). "All" is the sum of all risk groups, including non-key populations. The negative numbers of new HIV diagnoses in panel a) are because, in the long term, the intervention decreases the number of HIV infections that can be diagnosed by decreasing the number of new HIV infections, while the intervention particularly increases new HIV diagnoses among key populations.

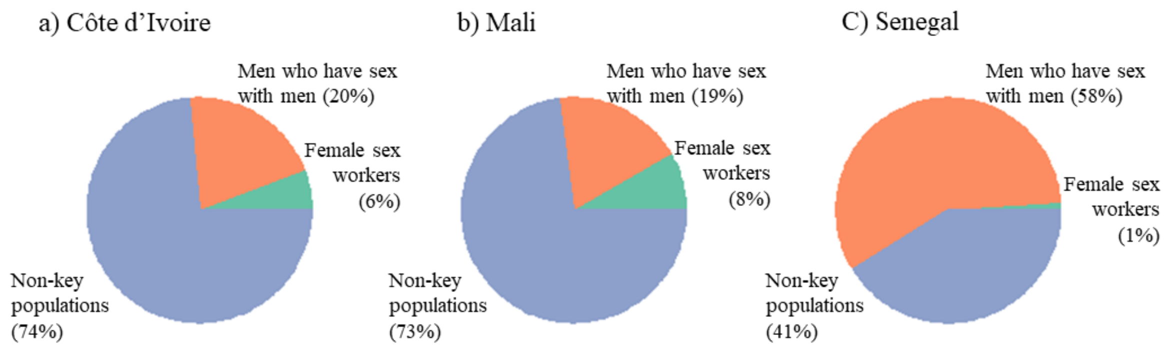

**Figure S11a:** Estimated distributions of infections averted by ATLAS scale-up by risk group over 2019-2038 in a) Cote d'Ivoire, b) Mali, and C) Senegal

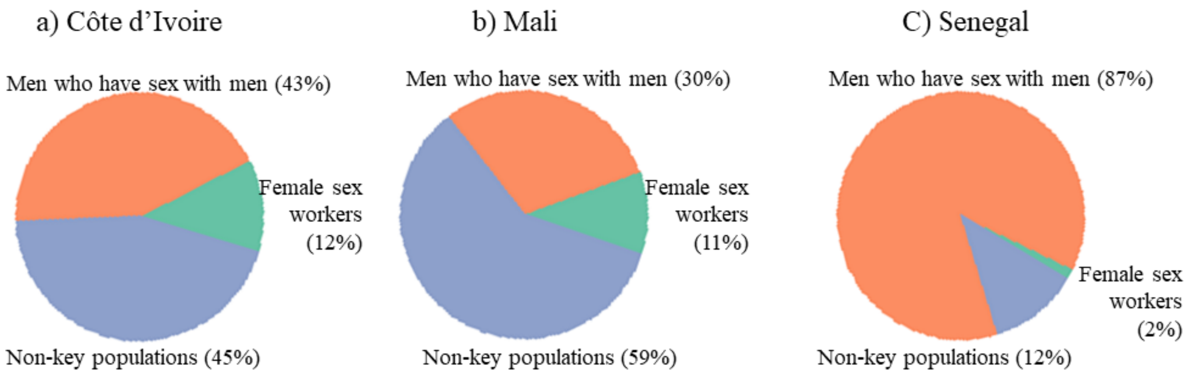

**Figure S11b:** Estimated distributions of HIV-related averted by ATLAS scale-up by risk group over 2019-2038 in a) Cote d'Ivoire, b) Mali, and C) Senegal

**Table S6a:** Estimated increase in HIV diagnosis coverage (expressed in percentage points) due to ATLAS and HIVST scale-up among KP in Côte d'Ivoire, Mali, and Senegal from 2019 compared to a counterfactual scenario with no HIVST. Median and 90%UI (5th and 95th percentiles) of estimates are shown.

| | | Côte d'Ivoire | | | Mali | | | Senegal | | |
|---|---|---|---|---|---|---|---|---|---|---|
| HIVST distribution scenario | Population | After 3 years | After 10 years | After 20 years | After 3 years | After 10 years | After 20 years | After 3 years | After 10 years | After 20 years |
| ATLAS-only (2019-2021) | Overall | 0.2pp (0.2-0.3) | 0.1pp (0.07-0.2) | 0.03pp (0.01-0.07) | 0.9 pp (0.5-1.5) | 0.7pp (0.4-1.4) | 0.4pp (0.2-0.9) | 1.1 pp (0.3-2.7) | 0.9pp (0.2-2.3) | 0.04pp (-0.1-0.6) |
| | FSW | 2.0pp (1.2-3.6) | 0.3pp (0.1-0.7) | 0.05pp (0.02-0.1) | 7.7 pp (2.9-14.6) | 1.4pp (0.5-3.9) | 0.6pp (0.2-1.8) | 0.0 pp (0.1-1.1) | 0.1pp (0.06-0.6) | 0.0pp (0.0-0.1) |
| | Clients | 0.3pp (0.2-0.4) | 0.2pp (0.1-0.4) | 0.04pp (-0.002-0.1) | 0.5 pp (0.3-1.1) | 1.0pp (0.5-1.8) | 0.4pp (0.1-1.0) | 0.1 pp (0.0-0.1) | 0.1pp (0.07-0.3) | 0.09pp (0.04-0.3) |
| | MSM | 4.6pp (3.1-8.3) | 3.2pp (1.7-5.7) | 1.1pp (0.4-2.6) | 9.3 pp (3.9-14.7) | 4.7pp (1.6- 9.6) | 1.5pp (0.4-4.9) | 5.7 pp (1.0-17.2) | 2.5pp (0.6-8.1) | 0.1pp (-0.1-1.4) |
| HIVST scale-up (2019-2038) | Overall | 0.2pp (0.2-0.3) | 1.3pp (0.8-1.9) | 1.2pp (0.7-2.0) | 0.9 pp (0.5-1.5) | 3.6pp (2.0-6.4) | 4.8pp (2.5- 9.7) | 1.1 pp (0.3-2.7) | 10.6pp (5.3-16.8) | 13.6pp (6.3-21.4) |
| | FSW | 2.0pp (1.2-3.6) | 7.6pp (4.2-11.7) | 5.8pp (3.3-9.2) | 7.7 pp (2.9-14.6) | 13.7pp (5.9-24.2) | 12.1pp (4.7-24.0) | 0.0 pp (0.1-1.1) | 4.1pp (0.8-14.7) | 3.1pp (1.1-13.2) |
| | Clients | 0.3pp (0.2-0.4) | 2.3pp (1.7-3.3) | 2.6pp (1.6-4.0) | 0.5 pp (0.3-1.1) | 4.1pp (2.4-6.6) | 5.7pp (3.2-10.2) | 0.1 pp (0.0-0.1) | 1.6pp (1.0-2.7) | 2.7pp (1.6-4.3) |
| | MSM | 4.6pp (3.1-8.3) | 30.3pp (20.0-45.4) | 27.1pp (16.0-46.0) | 9.3 pp (3.9-14.7) | 31.3pp (19.9-44.8) | 32.1pp (18.0-49.1) | 5.7 pp (1.0-17.2) | 40.5pp (23.3-59.6) | 36.2pp (19.7-57.2) |

| **Table S6b:** Estimated proportion of averted infections due to ATLAS and HIVST scale-up among KP in Côte d'Ivoire, Mali and Senegal from 2019 compared to a counterfactual scenario with no HIVST. Median and 90%UI (5th and 95th percentiles) of estimates are shown. | | | | | | | | | | |
|---|---|---|---|---|---|---|---|---|---|---|
| | | Côte d'Ivoire | | | Mali | | | Senegal | | |
| HIVST distribution scenario | Population | Over ATLAS (2019-2021) | Over 10 years (2019-2028) | Over 20 years (2019-2038) | Over ATLAS (2019-2021) | Over 10 years (2019-2028) | Over 20 years (2019-2038) | Over ATLAS (2019-2021) | Over 10 years (2019-2028) | Over 20 years (2019-2038) |
| ATLAS-only (2019-2021) | Overall | 0.1% (0.09-0.2) | 0.4% (0.3-0.6) | 0.4% (0.2-0.6) | 0.7% (0.4-1.2) | 2.1%( 1.1-3.7) | 2.2% (1.2-4.1) | 1.4% (0.8-2.3) | 3.3% (1.7-6.3) | 2.7% (1.2-6.1) |
| | FSW | 0.2% (0.1-0.3) | 0.6% (0.4-1.1) | 0.6% (0.3-1.1) | 0.4% (0.2-0.8) | 2.2%( 1.1-3.6) | 2.5% (1.2-4.4) | 0.2% (0.1-0.3) | 0.6% (0.3-1.1) | 0.7% (0.4-1.6) |
| | Clients | 0.4% (0.2-0.8) | 0.7% (0.4-1.6) | 0.5% (0.3-1.3) | 2.0% (0.8-3.4) | 3.9% (1.8-7.1) | 3.4% (1.6-6.8) | 0.6% (0.4-1.1) | 1.0% (0.6-2.0) | 1.0% (0.6-2.5) |
| | MSM | 1.5% (1.0-2.7) | 4.6% (3.1-7.5) | 4.3% (2.7-6.9) | 4.0% (1.8-6.5) | 9.7% (4.4-16.1) | 8.7% (3.7-16.3) | 3.1% (1.6-5.7) | 5.9% (2.6-11.5) | 3.7% (1.5-8.3) |
| HIVST scale-up (2019-2038) | Overall | 0.1% (0.09-0.2) | 1.6% (1.0-2.4) | 2.9% (1.7-4.6) | 0.7% (0.4-1.2) | 5.3% (3.0-8.9) | 9.3% (5.2-16.6) | 1.4% (0.8-2.3) | 16.2% (10.0-23.1) | 29.2% (17.8-40.1) |
| | FSW | 0.2% (0.1-0.3) | 2.3% (1.6-3.6) | 4.9% (3.1-7.4) | 0.4% (0.2-0.8) | 4.8% (2.7-7.7) | 9.3% (5.0-16.1) | 0.2% (0.1-0.3) | 2.7% (1.7-4.5) | 6.9% (4.2-10.4) |
| | Clients | 0.4% (0.2-0.8) | 3.4% (1.8-5.9) | 5.5% (2.9-9.1) | 2.0% (0.8-3.4) | 10.0% (5.1-17.6) | 14.0% (7.4-23.9) | 0.6% (0.4-1.1) | 5.9% (3.7- 9.9) | 11.6% (7.0-17.2) |
| | MSM | 1.5% (1.0-2.7) | 20.5% (15.3-27.6) | 31.4% (22.6-41.8) | 4.0% (1.8-6.5) | 27.3% (18.0-36.2) | 42.2% (28.6-55.4) | 3.1% (1.6-5.7) | 28.0% (21.3-37.5) | 41.9% (31.2-54.4) |

| **Table S6c:** Estimated proportion of averted HIV-related deaths due to ATLAS and HIVST scale-up among KP in Côte d'Ivoire, Mali and Senegal from 2019 compared to a counterfactual scenario with no HIVST. Median and 90%UI (5th and 95th percentiles) of estimates are shown. | | | | | | | | | | |
|---|---|---|---|---|---|---|---|---|---|---|
| | | Côte d'Ivoire | | | Mali | | | Senegal | | |
| HIVST distribution scenario | Population | Over ATLAS (2019-2021) | Over 10 years (2019-2028) | Over 20 years (2019-2038) | Over ATLAS (2019-2021) | Over 10 years (2019-2028) | Over 20 years (2019-2038) | Over ATLAS (2019-2021) | Over 10 years (2019-2028) | Over 20 years (2019-2038) |
| ATLAS-only (2019-2021) | Overall | 0.05% (0.02-0.09) | 0.2% (0.1-0.3) | 0.3% (0.2-0.4) | 0.09% (0.03-0.2) | 0.7% (0.4-1.3) | 1.1% (0.6-2.1) | 0.2% (0.06-0.5) | 1.5% (0.8-3.1) | 1.9% (1.0-4.3) |
| | FSW | 0.5% (0.1-1.0) | 1.4% (0.6-2.6) | 1.1% (0.5-1.8) | 0.6% (-0.02-2.2) | 3.9% (1.7-8.5) | 3.5% (1.7-7.4) | 0.2% (0.05-0.5) | 0.6% (0.3-1.3) | 0.6% (0.4-1.2) |
| | Clients | 0.05% (0.02-0.09) | 0.3% (0.2-0.5) | 0.4% (0.2-0.7) | 0.08% (0.04-0.2) | 0.7% (0.4-1.3) | 1.4% (0.7-2.5) | 0.01% (0.004-0.04) | 0.1% (0.08-0.3) | 0.3% (0.2-0.5) |
| | MSM | 0.9% (0.4-1.4) | 3.5% (2.2-5.7) | 3.7% (2.2-5.9) | 1.2% (0.2-3.4) | 6.6% (2.9-12.3) | 7.5% (3.3-12.8) | 1.0% (0.2-2.2) | 5.0% (2.1-10.9) | 5.0% (2.0-10.5) |
| HIVST scale-up (2019-2038) | Overall | 0.05% (0.02-0.09) | 0.7% (0.4-1.3) | 1.7% (1.1-2.6) | 0.09% (0.03-0.2) | 1.5% (0.8-2.7) | 3.8% (2.0-6.2) | 0.2% (0.06-0.5) | 5.5% (3.2-9.4) | 15.1% (9.1-24.8) |
| | FSW | 0.5% (0.1-1.0) | 4.7% (2.2-8.6) | 8.5% (4.6-12.9) | 0.6% (-0.02-2.2) | 8.8% (4.0-16.8) | 13.7% (7.3-23.9) | 0.2% (0.05-0.5) | 2.7% (1.5-5.9) | 6.3% (3.8-11.5) |
| | Clients | 0.05% (0.02-0.09) | 0.9% (0.6-1.3) | 2.7% (1.8-3.9) | 0.08% (0.04-0.2) | 1.4% (0.8-2.4) | 4.5% (2.7-7.5) | 0.01% (0.004-0.04) | 0.4% (0.2-0.8) | 2.2% (1.4-3.5) |
| | MSM | 0.9% (0.4-1.4) | 13.2% (7.7-18.0) | 24.2% (17.0-31.3) | 1.2% (0.2-3.4) | 15.9% (9.3-26.6) | 29.7% (20.0-43.3) | 1.0% (0.2-2.2) | 18.4% (12.4-27.4) | 37.8% (26.9-50.1) |

#### Sensitivity analysis

We conducted a sensitivity analysis to assess the influence of key assumptions of our main HIVST scale-up scenario on the secondary distribution of self-tests by risk groups (i.e., approximatively doubling the fraction of tests received by non-KPs, **Tables S4a,b**), age distribution of HIVST users (based on a phone survey conducted by ATLAS[144] instead of programmatic data, with little change in average age of population, **Figure S2**), more HIVST distributed over time (reflecting population growth, instead of a constant number of kits distributed annually) (**Table S5**), higher uses of distributed tests (100% instead of 80%), twice higher likelihood of receiving a HIVST kit for people having never tested for HIV and undiagnosed PLHIV (instead of the uniform likelihood of receiving a kit), higher rate of confirmation of reactive HIVST (80% instead of 50%), less substitutions of conventional test for HIVST (none instead of 20-40%), and higher accuracy of self-tests (i.e., 100%/100% sensitivity/specificity instead of 92/99%).

The estimated fraction of new HIV infections and deaths averted by scaling up HIVST among KP remained qualitatively similar for all alternative assumptions explored and countries (**Figures S12a,b**). The largest difference was observed when doubling the uptake of HIVST among undiagnosed PLHIV compared to HIV-uninfected populations (around 1·7-times more new HIV infections are averted over 10 years). Interestingly, assuming higher levels of secondary distribution of HIVST towards non-KP compared to the base-case (so that ~36% of self-tests kits are used by non-KP during scale-up, vs ~66% in the base-case) reduced the fraction of infections or deaths averted over 2019-2028 by ~1·3-fold compared to the base-case).

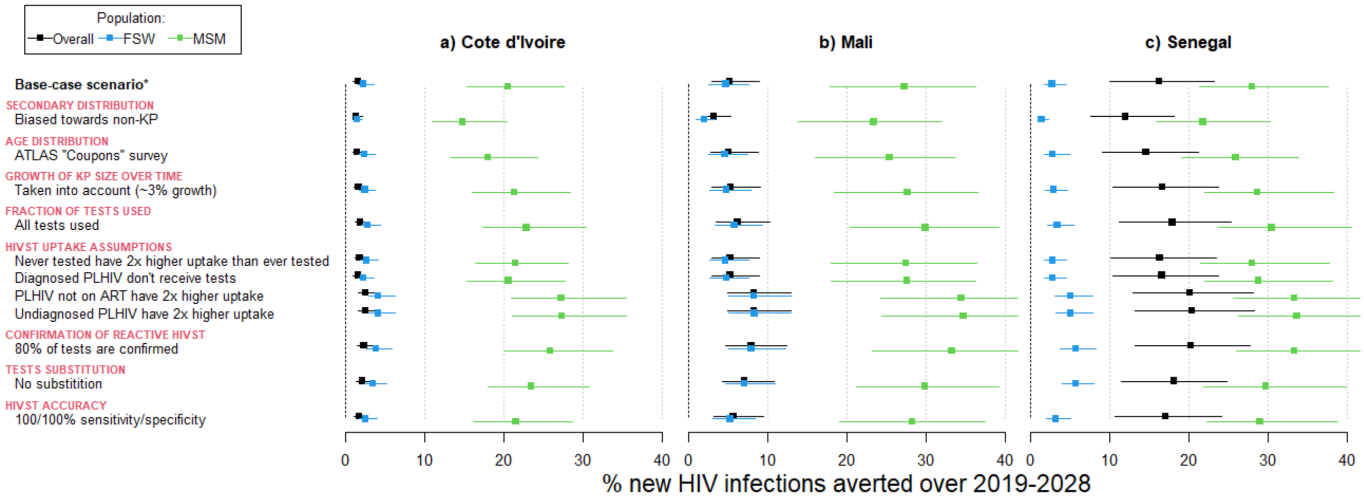

*assumes simplified secondary distribution of HIVST, age distribution from programmatic data, KP population growth not taken into account, 80% of HIVST are used, with uniform uptake across population groups, 50% of reactive self-tests are confirmed, 20-40% of self-tests replace conventional tests, and self tests have 92/99% sensitivity/specificity

**Figure S12a:** Estimated fraction of new HIV infections averted by a scale-up of HIVST among KP over 2019-2028 in the three ATLAS countries within our base-case scenario (top row) as well as sensitivity scenarios. The impact was calculated overall (black squares and intervals), as well as among FSW (blue squares and intervals) and MSM (green squares and intervals). Squares represent median estimates across model fits, whereas error bars represent 90% UI of estimates (5$^{th}$ and 95$^{th}$ percentiles of estimates across model simulations).

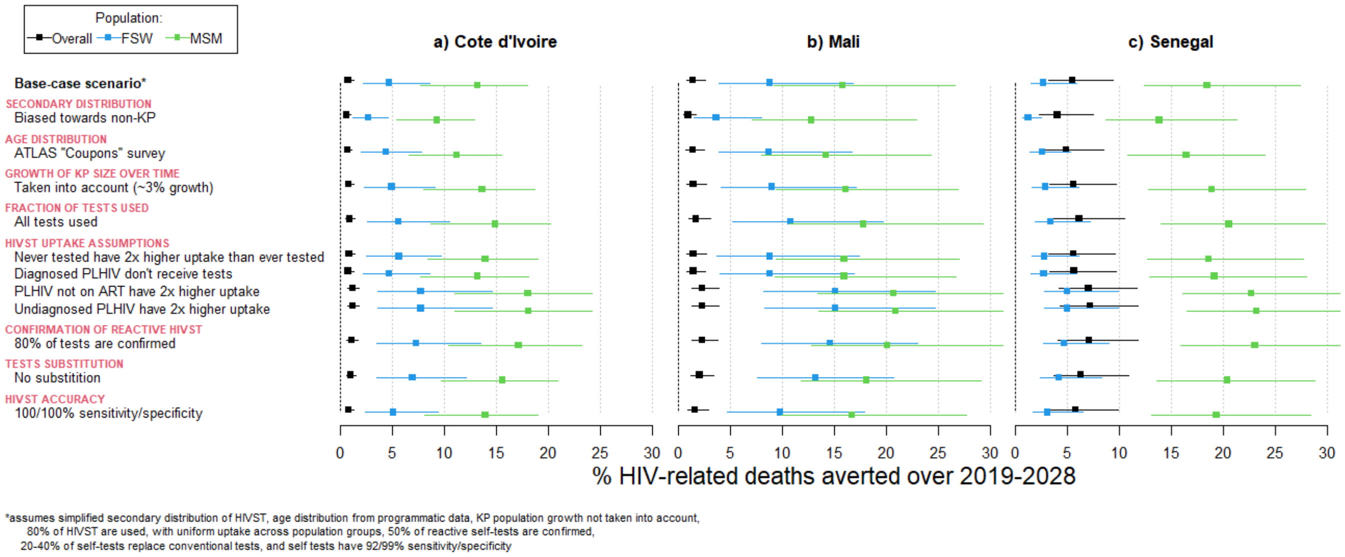

**Figure S12b:** Estimated fraction of HIV-related deaths averted by a scale-up of HIVST among KP over 2019-2028 in the three ATLAS countries within our base-case scenario (top row) as well as sensitivity scenarios. The impact was calculated overall (black squares and intervals), as well as among FSW (blue squares and intervals) and MSM (green squares and intervals). Squares represent median estimates across model fits, whereas error bars represent 90% UI of estimates ($5^{th}$ and $95^{th}$ percentiles of estimates across model simulations). Impacts are larger among MSM because of greater numbers of tests distributed to them on average (see Figure S3), and because new diagnosis decreases the risk of MSM to die from HIV and prevent more infections among their partners.
